# Host blood protein biomarkers to screen for Tuberculosis disease: a systematic review and meta-analysis

**DOI:** 10.1101/2024.05.24.24307893

**Authors:** Mary Gaeddert, Kerstin Glaser, Bih H. Chendi, Ayten Sultanli, Lisa Koeppel, Emily L. MacLean, Tobias Broger, Claudia M. Denkinger

## Abstract

**Introduction:** Non-sputum tests are needed to improve TB diagnosis and close the diagnostic gap. The World Health Organization target product profile (TPP) for point-of-care (POC) screening tests requires minimum sensitivity 90% and specificity 70%. Our objective was to identify host blood protein biomarkers meeting TPP criteria.

**Methods:** A systematic review was conducted and reported following PRISMA guidelines. Data extraction and quality assessment with QUADAS-2 were completed for included studies. Heterogeneity was assessed. For biomarkers reporting sensitivity and specificity in at least four studies, a random-effects meta-analysis was performed for biomarkers with similar cut-offs.

**Results:** We screened 4,651 citations and included 65 studies that enrolled 16,010 participants and evaluated 156 host proteins. Most (47/65) studies enrolled adult pulmonary TB (PTB), with 15 studies in adult extra-pulmonary TB and 5 in children. Small early-stage discovery studies with case-control design were common (24/65) and had high risk of bias. For adult PTB, CRP, IP-10, NCAM-1, and SAA met TPP criteria in high-quality studies. There was a high degree of heterogeneity in biomarker cut-offs and study design. CRP at 10mg/L cut-off was meta-analyzed from 10 studies; pooled sensitivity 86% (95% CI: 80-95) and pooled specificity 67% (95% CI: 54-79). In people living with HIV (6 studies) CRP pooled sensitivity was 93% (95% CI: 90-95) and pooled specificity 59% (95% CI: 40-78).

**Discussion:** We identified promising biomarkers that performed well in high-quality studies. Data overall are limited and highly heterogenous. Further standardized validation across subgroups in prospective studies is needed before translating into POC assays.

## Introduction

Tuberculosis (TB) was the second leading cause of death by a single infectious disease worldwide in 2022 after COVID-19 (1). Even though the disease is curable, individuals must first be diagnosed before starting on treatment, and the limitations of current TB diagnostics is one factor contributing to the estimated 3 million TB cases that were missed due to underdiagnosis or underreporting in 2022 (1). With 87% of the global TB cases concentrated in only 30 high burden countries (1), many of which have limited resources, there is a need for new diagnostics that have wider reach.

Current diagnostics for TB disease are mainly sputum-based, and there are well-recognized challenges with obtaining adequate sputum samples for testing, particularly in people living with HIV (PLHIV) and children (2). Sputum-based methods are also unsuitable for the detection of extra-pulmonary TB disease (EPTB) (3). A molecular WHO-recommended rapid diagnostic (mWRD) was used as the initial test for 47% of the newly diagnosed TB cases in 2022, and the goal is to reach 100% coverage by 2027 (1). In order to close this gap and improve TB diagnostics globally, there is a need for non-sputum based rapid tests that can be used in a point of care (POC) setting where they would reach more patients earlier in the disease course.

In 2014, the World Health Organization (WHO) issued target product profiles (TPPs) to guide development of new TB diagnostics which included recommendations for a screening test (4). The goal is to develop a high-sensitivity test that can be used to rule out adults and children with presumptive TB at lower levels of the healthcare system. A positive result on the screening test would require a second test with higher specificity as confirmation. The minimum accuracy recommended by the TPP was sensitivity of 90% and specificity of 70%. The TPP also recommends operational characteristics to enable a better reach in resource-limited settings. This includes non-sputum samples that are easy to collect such as blood from a finger-stick, testing platforms that can be used at POC, and a low cost per test.

Host blood biomarkers could possibly meet these accuracy targets as suggested by results from a recent study on the Xpert MTB Host Response assay using a 3-gene signature (sensitivity 90%; specificity of 63%) (5). However, tests based on mRNA targets or other genomic signatures require complex equipment and are unlikely to meet stated cost targets (6, 7). Blood protein biomarkers and biomarker signatures are more likely to translate into affordable POC tests, such as a lateral flow assay. C-reactive protein (CRP) has been evaluated in many studies, and while it does not meet performance targets in meta-analyses, it can be measured using POC testing platforms (8, 9). The question remains whether other host blood protein biomarkers, or a combination of markers, could reach the TPP accuracy targets. Well-performing markers would also need to be measurable by POC platforms, ideally using blood from a finger-prick sample.

This systematic review focused on blood host proteins with the goal of identifying markers that meet the TPP performance criteria and could be translated to POC tests. The primary objective was to review the diagnostic accuracy of host proteins in adult pulmonary TB, and the secondary objectives were to review host proteins in adult extrapulmonary and childhood TB.

## Methods

### Search strategy

Studies were identified through a systematic search of the databases EMBASE, PubMed, Cochrane Controlled Trials Register, Scopus, the Web of Science, clinicaltrials.gov, and African Journals Online. The search was conducted for all studies from 1 January 2010 to 5 October 2023 with no language restrictions (Table S1). We also identified papers by reviewing the citations of included papers and relevant reviews. The systematic review protocol and search strategy are registered on PROSPERO (registration number: CRD42022298906) and followed PRISMA reporting guidelines (10). We included randomized clinical trials, cohort studies, case-control studies, and cross-sectional studies. Studies with less than 20 TB cases positive by either MRS or CRS were excluded.

### Index Test

Studies using index tests able to identify proteins in blood were included. The major testing platforms are enzyme-linked immunosorbent assay (ELISA), immunoassays in general, multiplexing platforms such as Luminex and Meso Scale Discovery (MSD), and mass spectrometry. Studies that used blood samples stimulated with TB antigens were excluded as they would likely not meet operational characteristics necessary for use at POC. While antibodies are part of the proteome, we excluded antibody tests as they have shown unreliable diagnostic accuracy and WHO strongly recommended against their use in 2011 (11).

### Reference standard

To be considered for inclusion, adult PTB studies must have used a microbiological reference standard (MRS) based on culture or a mWRD on any sample (12). TB cases were those who tested positive on at least one culture or mWRD and control groups were negative on all tests. Bacteriological confirmation is difficult for extra-pulmonary and childhood TB due to their paucibacillary nature and diagnosis often relies on a combination of symptoms, imaging, and microbiologic testing (3). Therefore, we included studies that used a composite reference standard (CRS) for the childhood and EPTB groups and extracted details of the CRS definition.

### Study selection and data extraction

Results of the literature search were exported to EndNote and duplicates were removed. For title and abstract screening, a sample of 5% were screened by three reviewers (MG, KG, AS), discussed to reach concordance, and then each reviewer independently assessed the remaining results. Two reviewers (MG and KG) independently reviewed the full text and conducted data extraction; discrepancies were resolved by discussion between the two reviewers, or by decision of a third reviewer (CMD). Data was extracted from eligible studies using GoogleForms. Study quality was assessed only for adult pulmonary TB studies using a modified version of the Quality Assessment of Diagnostic Accuracy Studies (QUADAS-2) tool (13). QUADAS signalling questions were modified to fit the review question, and the description of questions and scoring is in Table S2. Covidence was used for full text review and QUADAS extraction.

When studies reported results from both discovery and validation cohorts, only data from the validation cohort was extracted. For studies that reported biomarker results at multiple cut-off points, we extracted the results with higher sensitivity in order to reach the TPP goal of 90% sensitivity. We reported the results of each biomarker separately from studies that reported results of multiple biomarkers.

The most clinically relevant comparison group for a screening test is other patients who present with symptoms of presumptive TB and are later diagnosed with other respiratory diseases (ORD), as they likely have an inflammatory process causing symptoms and will be the target population for routine use. A hierarchy of control groups was developed (Supplementary Methods, (14)), and if studies used multiple control groups, results from the group with the highest clinical relevance were extracted. Biomarker signatures were also extracted. Biomarker names were harmonized and abbreviations are listed in Table S3. All results were reported separately by HIV status and country of enrolment where information was available.

### Statistical Analysis

When studies did not report 95% confidence intervals (CI’s) for sensitivity and specificity, we calculated the CI’s using the Wilson Score Interval. The Deeks test for funnel-plot asymmetry was performed to investigate publication bias for diagnostic test accuracy meta-analyses using the ‘midas, pubbias’ command in Stata; a *p*-value < 0.10 for the slope coefficient indicates significant asymmetry (15, 16). If at least four studies reported sensitivity and specificity for adult pulmonary TB, heterogeneity was assessed by visually examining forest plots and hierarchical summary receiver operating characteristic (HSROC) plots and underlying causes of heterogeneity in study design were investigated. A random-effects meta-analysis was done using the ‘meta’ package in Stata for biomarkers with similar cut-offs. Stata 17 (STATA Corporation, College Station, TX, USA) was used for all analyses.

## Results

### Summary of studies

The literature search resulted in 4,651 articles after de-duplication. After title and abstract screening, 369 papers were eligible for full-text review. Of these, 304 were excluded, leaving 65 studies in the systematic review (Figure 1); 45 provided data for adult PTB, 13 for EPTB, 2 for both PTB and EPTB, and 5 for childhood TB. The 65 studies enrolled a total of 16,010 participants from 17 different countries across all continents except Australia. Most studies enrolled participants from the regions of Southeast Asia (26/65, 40%) and Sub-Saharan Africa (24/65, 37%), with South Africa (12/65, 18%), India (12/65, 18%), and China (11/65, 17%) being the most frequent countries.

**Figure 1.**
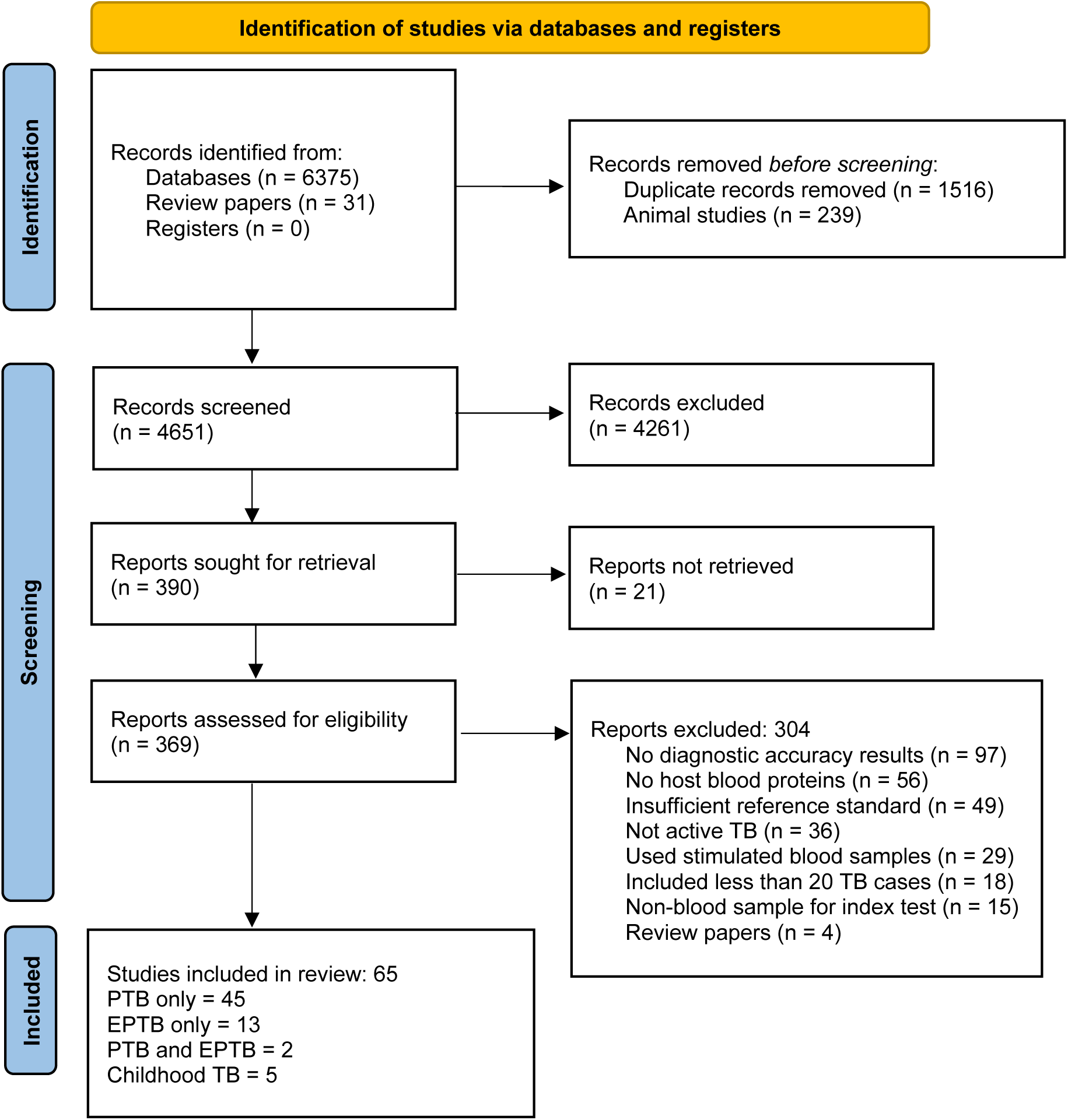
PRISMA flow diagram of study selection.

The majority of studies used serum (41/65, 63%) and plasma (19/65, 29%) for biomarker testing, and whole blood was used in 4 studies (6%). All studies that reported sample condition used frozen blood (44/65, 69%) except 4 studies (6%) that used fresh blood.

Immunoassays in general (37/65, 54%), and ELISA (27/65, 39%) specifically, were the most common index test, followed by Luminex (18/65, 26%) and Turbidimetry (6/65, 9%). Mass spectrometry, MSD, nephelometry, SomaScan, the Simoa assay, and the peroxidase method were used in one study each.

### Methodological quality of studies

Many papers reviewed were early-stage discovery studies that enrolled small numbers of known TB cases and controls. Of the 47 papers that enrolled adult patients with PTB, 24 (51%) used a case-control design, resulting in a higher risk of bias in the QUADAS patient selection domain (Figure 2). The use of a healthy comparator group without TB symptoms resulted in a high concern of applicability for patient selection. Studies using healthy controls did not apply the same reference standard testing to the non-TB comparator group, increasing the risk of bias in the ‘flow and timing’ domain. Studies often did not report whether the index test was interpreted in blinded manner from the reference standard results, and cut-offs were often chosen based on results of each study and not pre-specified or validated in other cohorts. In the domain concerning applicability of index test for POC use, only four studies (17–20) used a POC assay for CRP, so all other studies had a high concern for applicability. The inclusion criteria requiring an MRS resulted in an overall low risk of bias and low applicability concerns with respect to the reference standard. The detailed list of results by study for adult PTB are in Table S4.

**Figure 2.**
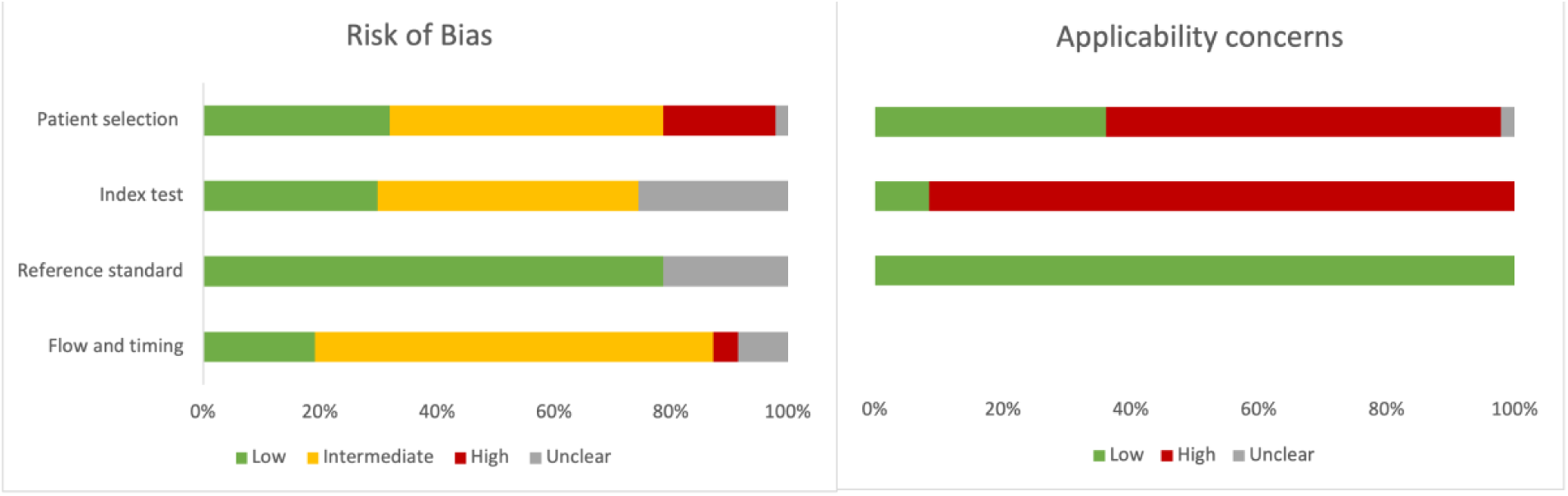
Summary of QUADAS-2 assessment for risk of bias and applicability concerns.

The funnel plot of included adult PTB studies shows a high degree of symmetry and does not indicate substantial publication bias (p-value=0.62) (Figure S1).

### Summary of biomarkers

A total of 156 host proteins were evaluated across all included studies. The 47 adult PTB studies evaluated the diagnostic accuracy of 102 individual host blood proteins and 18 different signatures by calculating the sensitivity and specificity at a chosen cut-off value (Tables S5-S8). The majority of biomarkers (73/102, 72%) have results from only one study. The biomarker with the most evidence was CRP, reported by 19 studies. Some studies reported CRP performance separately by HIV status and cut-off, so there are 30 unique results included.

In total, 19 individual biomarkers tested by 14 separate studies met the TPP criteria of sensitivity > 90% and specificity > 70% in adult PTB. Table 1 shows the performance of each marker and the summary of study quality. Eighteen markers met TPP in HIV-negative patients: CALCOCO2 (21), CD14 (22), CRP (23), Ferritin (24), GBP1 (21), HO-1 (25, 26), IFIT3 (21), IFITM3 (21), IP-10 (18), MMP-1 (19), OPN (27), PD-L1 (21), SAA (25), SAMD9L (21), SELL (22), SNX10 (21), TIMP-2 (19), and TIMP-4 (28). All of these studies had a high overall risk of bias due to the use of LTBI or healthy control groups in the absence of patients with ORD. Furthermore, all studies choose the cut-off value which provided the highest accuracy post-hoc and did not use a pre-defined cut-off. Although these biomarkers met TPP in at least one study, there is a wide range of performance and results from other studies on the same biomarker had lower accuracy (Figure 3a).

**Figure 3.**
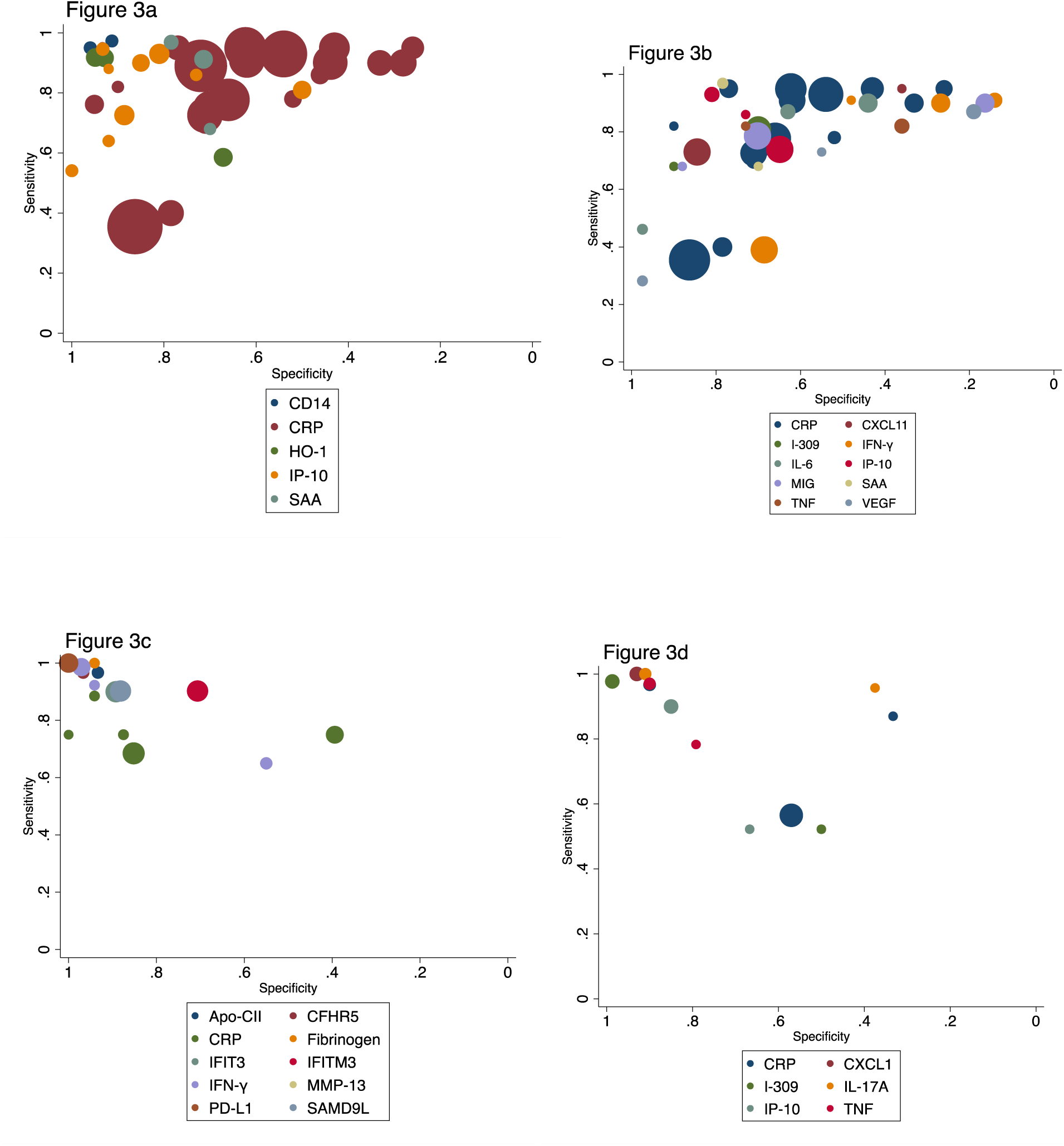
Summary plots. 3a. Complete results of adult PTB biomarkers that met the TPP criteria in two or more studies 3b. Complete results of adult PTB biomarkers tested in two or more studies with limited bias 3c. Complete results of adult Extrapulmonary TB biomarkers that met the TPP criteria in one or more studies 3d. Complete results of childhood TB biomarkers that met the TPP criteria in one or more studies symbol colors represent different biomarkers and the size of the markers is proportional to sample size.

**Table 1.**
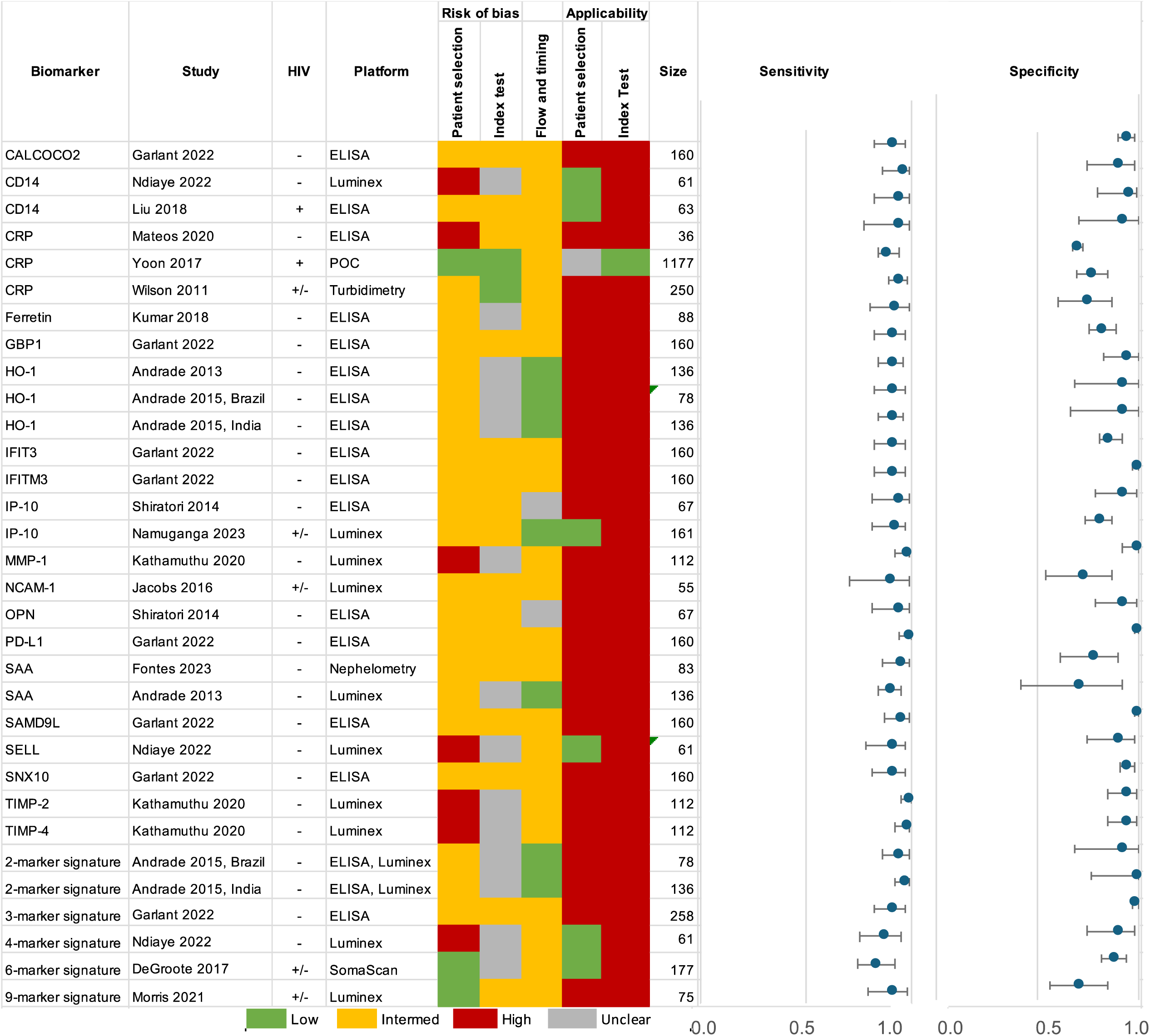
Summary of adult PTB biomarkers and signatures that meet the TPP criteria. Legend: 2-marker signatures: HO-1, MMP-1; 3-marker signature: CALCOCO2, IFITM3, SAMD9L; 4-marker signature: CLEC3B, ECM1, IP-10, SELL; 6-marker signature: SYWC, kallistatin, C9, gelsolin, testican-2, aldolase C; 9-marker signature: Fibrinogen, α-2-M, CRP, MMP-9, TTR, CFH, IFN-γ, IP-10, TNF. The Reference Standard results for Risk of Bias and Applicability were low for all studies.

In studies that reported results for PLHIV, only CD14 (29) met the TPP criteria for performance. This study used healthy controls and the cut-off value was chosen to reach the highest diagnostic accuracy for their cohort. In studies with a mix of HIV-positive and -negative adults, three individual biomarkers, CRP (30), IP-10 (31), and NCAM (32) met the TPP criteria in at least two of the studies. As these studies all used patients with symptoms of presumed TB as the comparator, the risk of bias was low. Nevertheless, heterogeneity was substantial and complete results from studies that used a clinically relevant population with a low risk of bias shows that many biomarkers did not perform as well (Figure 3b).

In addition, five biomarker signatures met the TPP criteria. A combination of HO-1 and MMP-1 using participants with LTBI as the comparator group was reported separately by sites in Brazil and India (26). A three-marker signature of CALCOCO2, IFITM3, and SAMD9L tested lysed whole blood with ELISA, comparing TB cases in India to asymptomatic contacts from India and the United Kingdom (21). The four-marker signature of CLEC3B, ECM1, IP-10, and SELL used Luminex to test plasma samples from TB cases and LTBI controls in Madagascar (22). The 6-marker signature by DeGroote (SYWC, kallistatin, C9, gelsolin, testican-2, and aldolase C) was developed by testing serum samples from patients with symptoms of presumptive TB with SomaScan (33). The signature’s performance was reported separately by HIV status and had an AUC>0.9 for all groups; although sensitivity is slightly below 90%, specificity is above 80%. A study of a nine-marker signature consisting of fibrinogen, α-2-M, CRP, MMP-9, TTR, CFH, IFN-γ, IP-10, and TNF-α enrolled patients with symptoms of presumptive TB from South Africa and Malawi, testing serum samples on Luminex (34).

Adult PTB studies which reported only the area under the curve (AUC) are summarized in Table S9. Of the 45 individual markers, 12 had an AUC>0.9, which is likely to meet TPP criteria if a cut-off had been defined to calculate sensitivity and specificity: C1q (35), I-309 (36), IFN-γ (37), IL-2 (37), IL-5 (37), IL-6 (37), IL-10 (37), IL-17A (37), IL-1α (37), IP-10 (37), MIG (37), and TNF (37). All of these markers were in HIV-negative patients, and 8/12 came from one study (37) that tested both drug-susceptible and drug-resistant TB cases compared to LTBI or healthy controls. Two signatures reached an AUC of 0.9: a 2-marker combination of SYWC and I-309 in PLHIV and a 3-marker combination of I-309, SYWC, and kallistatin in a population with mixed HIV status across three countries (38).

The CRS criteria for the EPTB and childhood TB studies are listed in Table S10. Two EPTB studies (21, 36) and two childhood TB studies (39, 40) reported that all TB cases were positive on either NAAT or culture. The other studies used a combination of AFB smear or culture, and three studies (28, 41, 42) did not provide details on the reference standard.

Ten individual biomarkers met the TPP criteria for adult EPTB: Fibrinogen (43) and IFN-γ (43) in a population with mixed HIV status, and Apo-CII (36), CFHR5 (36), IFIT3 (21), IFITM3 (21), IFN-γ (44), MMP-13 (28), PD-L1 (21), and SAMD9L (21) in HIV-negative patients (Figure 3c, full results in Table S11). Two signatures met the TPP criteria for EPTB: a five-marker signature consisting of CRP, NCAM, Ferritin, IL-8 and GDF-15 in population with mixed HIV-status (43) and a 2-marker signature of CFHR2 and CFHR3 in HIV-negative participants (36). There were no studies in the review that enrolled only PLHIV with EPTB.

A study by Garlant et al. tested a range of markers in both PTB and EPTB patients; IFIT3, IFITM3, PD-L1, and SAMD9L, met the TPP criteria in both groups (21). Importantly, the EPTB group was defined using an MRS. Also, IFN-γ performed well in two EPTB studies with HIV-negative participants that used a CRS (43, 44) and one study by Sampath, et al. with both drug-resistant and drug-susceptible HIV-negative PTB cases (37). While the Sampath study did not report sensitivity and specificity, the AUC for IFN-γ was 0.95 in drug-susceptible and 0.94 in drug-resistant cases.

Five childhood TB studies evaluated a total of 70 host blood proteins (Figure 3d, full results in Table S12). Four studies enrolled HIV-negative children and one study enrolled children with mixed HIV status. Two biomarkers met TPP criteria in the study using a MRS: IL-17A (40) and TNF (40). This study compared TB to ORD and enrolled a separate validation cohort to reduce the risk of bias. The remaining biomarkers meeting TPP were compared to a CRS: CRP in a study that enrolled children with both PTB and EPTB (45), and CXCL-1, I-309, and IP-10 in a study with PTB (46). Two signatures met TPP for childhood TB, a two-marker combination of I 309 and CXCL-1 (46) in children diagnosed with a CRS and a three-marker signature of TNF, IL-2, and IL-17A (40) using a MRS. All markers that met TPP in children also performed well in adult studies, except CXCL1. CRP and IP-10 met TPP criteria in both groups, and I-309, IL-17A, and TNF performed well in adult studies that reported only AUC.

### Meta-analysis

Heterogeneity was assessed for biomarkers that reported sensitivity and specificity in at least four studies for adult pulmonary TB (CRP, HO-1, IFN-γ, IL-6, IP-10, MIG, TNF, and VEGF). Due to the high level of heterogeneity for clear reasons like study-design, population included, index test, sample type, and cut-off value, but also for additional reasons (as visualized in forest plots and HSROC curves) a meta-analysis was only possible for CRP.

The results of CRP at a cut-off of 10mg/L were meta-analyzed from 10 studies (Figure 4a). The pooled sensitivity was 86% (95% CI: 80-95) and the pooled specificity with 67% (95% CI: 54-79) (Figure 4a). Visually assessing the forest plots and HSROC curves indicates the heterogeneity in CRP results was moderate for the sensitivity but high for specificity. The results of CRP in only PLHIV at a cut-off of 10mg/L were meta-analyzed from 6 studies (Figure 4b). The pooled sensitivity was 93% (95% CI: 90-95) and the pooled specificity was 59% (95% CI: 40-78). One study by Ciccacci, et al. from Mozambique that tested frozen plasma samples on ELISA was an outlier, reporting lower sensitivity but higher specificity than the others (47). This study enrolled PLHIV being screened for TB before starting antiretroviral therapy (ART).

**Figure 4.**
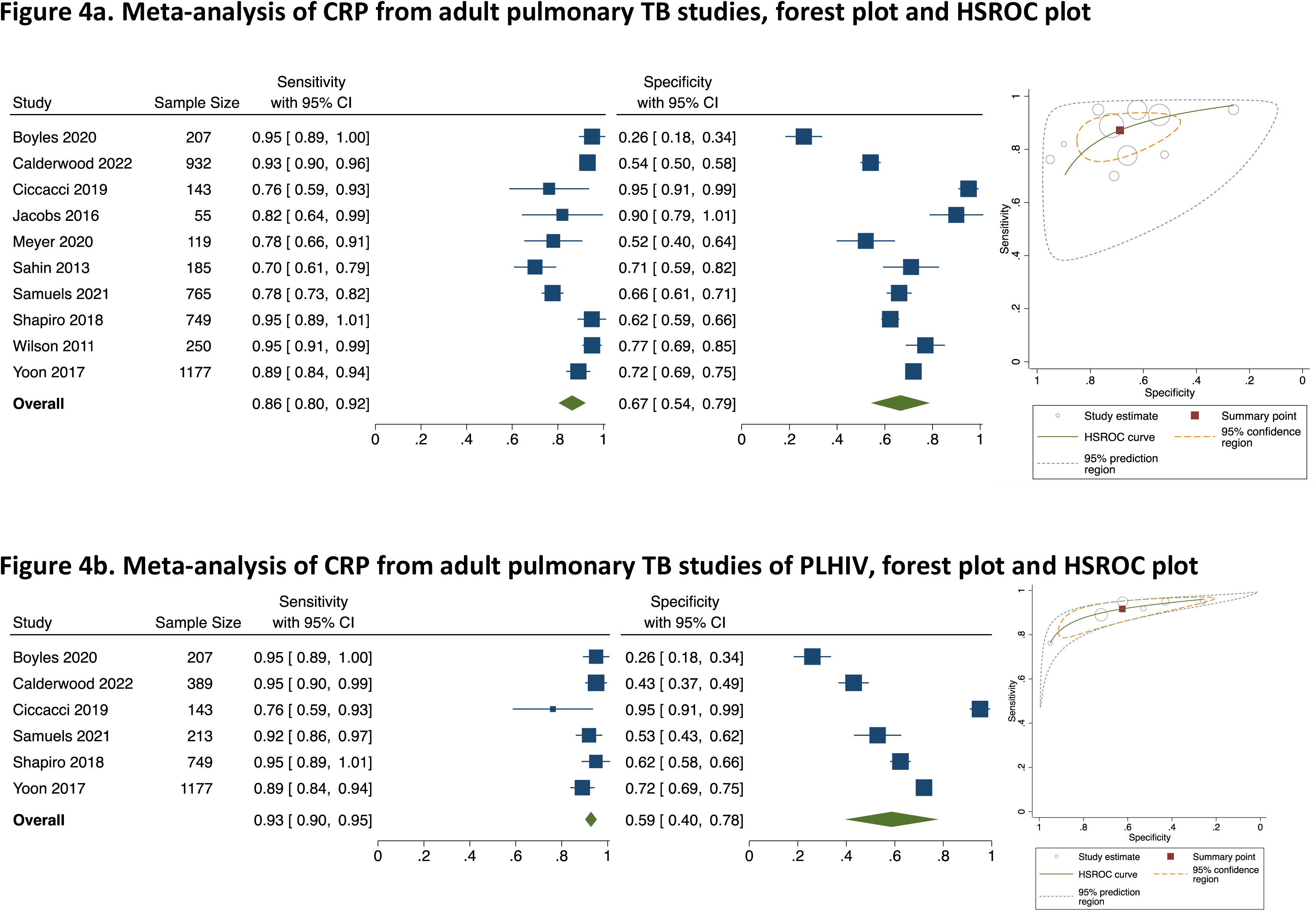
Results of CRP meta-analysis at 10mg/L cut-off.

## Discussion

This is the first systematic review to comprehensively look at host blood protein biomarkers for TB disease that could translate into POC assays. A number of individual biomarkers and biomarker signatures met the TPP performance criteria for a screening test and are promising. However, most markers were evaluated by only one study each in small study populations that were likely selected in a biased manner, and require further validation. For biomarkers that were evaluated in multiple studies, the heterogeneity in study designs and biomarker cut-offs make it difficult to draw broad conclusions.

When considering the risk of bias, the most promising individual host proteins for adult PTB are CRP (30), IP-10 (31), NCAM-1 (32), and SAA (48). These markers not only met TPP criteria but were evaluated in clinically relevant populations of patients with presumptive TB symptoms. Many other studies were early-stage discovery studies that used participants with LTBI or healthy controls, which likely overestimated biomarker performance (49).

Also, many studies excluded PLHIV, limiting the applicability of findings to this group. CD14 was the best performing marker out of the limited number tested in PLHIV (29), and also performed well in HIV-negative participants (22). In studies evaluating participants with ORD that enrolled a population of mixed HIV status, CRP (32) and IP-10 (31) had high accuracy.

CRP was the most-studied marker and our results support the findings of previous reviews that have shown it not to perform well enough as a single marker (8, 9, 50). While WHO recommends a CRP cut-off of 5 mg/L for PLHIV (51) our review did not have a sufficient number of results at this cut-off to perform a meta-analysis.

CRP was also the only biomarker with enough previous research to establish pre-defined cut-offs. Many other biomarkers have not been evaluated before, and studies were done for the purpose of discovery and the cut-off thus defined post-hoc to optimize performance. As the body of evidence for biomarkers is growing, it will be important to develop consensus cut-offs and conduct validation studies with fixed, pre-defined cut-offs. This will enable results to be compared across studies and perform meta-analyses in the future.

As the performance of individual markers often do not meet TPP requirements in clinically relevant populations, combinations of markers will likely be necessary. One of the best performing combinations in patients with symptoms of presumptive TB was the 6-marker signature by De Groote, et al. (33). Building off of this study, a recent analysis using machine-learning methods identified a 3-marker signature of I-309, SYWC, and kallistatin with an AUC of 0.90 (38). The other signatures that met TPP in studies with a low risk of bias for patient selection was a 4-marker combination with an AUC of 0.93 (22) and a 9-marker combination with an AUC of 0.84 (34). These results may indicate a plateau in performance around AUC 0.90, wherein adding additional biomarkers to a signature may not increase the diagnostic accuracy any further.

Ideally, the same markers would perform well across multiple populations, such as PTB and EPTB and between adults and children. However, our review did not identify many overlapping markers between patient groups. Possible reasons for this could be differences in the studies themselves (e.g. different reference standard) (52) but also in the host response to TB disease, particularly in children (53).

The best sample for blood-based POC tests would be fresh capillary (finger-prick) blood because it is easy to collect and does not require processing. However, most studies used frozen plasma or serum samples. While this is the most feasible way to do discovery and early validation studies, there are differences when a capillary sample is used (54, 55) and further validation studies on the relevant clinical sample will be needed.

The most commonly used platforms for protein measurement in this review were ELISA and Luminex, which differ in their limits of detection and quantification. While data generated on ELISA can be conceivably translated into a lateral-flow assay, the translation from a multiplexing platform such as Luminex is more difficult as it has superior sensitivity and a broader dynamic range than ELISA (56). Studies that used a POC CRP assay reported lower sensitivity on average than studies using other methods such as ELISA and Luminex, although heterogeneity in study designs makes it difficult to compare results between platforms directly. Of the biomarkers that met TPP in adult PTB, studies used a POC test (18), turbidimetry (30), and nephelometry (48) indicating that less sensitive assays can still achieve high diagnostic accuracy.

Multiplex lateral-flow assays (LFA) are a possible solution to achieve the TPP objective for a screening test; this would enable testing a combination of biomarkers on low-cost POC platform. Multiplex LFAs are not yet common and typically restricted to three markers with similar concentration ranges and LFAs have limited ability for quantification (57). However, advancements have been made in developing LFAs for cardiac biomarkers, including an up-converting phosphor technology-based lateral flow (UPT-LF) assay for diagnosis of acute heart failure as a POC test (58, 59). There is a study currently evaluating a 3-host protein marker lateral flow assay as a screening test for TB (60).

### Strengths

Our review conducted a comprehensive literature search and included studies conducted in 17 countries. Study quality was assessed using the standard QUADAS-2 tool, and the inclusion criteria requiring an MRS for adult PTB reduced the risk of bias for misclassifying the TB group.

### Limitations

Most biomarkers were investigated in one study each, limiting the meta-analysis to CRP. For studies that reported results at multiple cut-off values, the a-priori decision to extract the results with higher sensitivity resulted in a bias towards including the results with higher sensitivity and lower specificity. Despite our comprehensive literature search, there is a possibility that some papers could have been missed.

### Recommendations for future research

As there are few individual host proteins that meet the TPP requirements in high quality studies, further work should be done validating these markers and new combinations of markers in populations that reflect the intended use-case. Validation studies should be conducted using fresh blood samples on POC assays, and standardized, ideally pre-defined cut-off values should be used whenever possible. There is especially a need for more biomarker studies in children and PLHIV, both with PTB and EPTB disease.

## Conclusion

The large number of host blood protein biomarkers studied indicates a strong interest in this area of research, and some biomarkers have promising performance under controlled research conditions. However, further advancements are needed in testing on POC platforms as well as prospective validation studies using clinically relevant populations and sample types.

## Supplemental Materials

**Table S1:** Search strategy.

|  |  |  |
| --- | --- | --- |
| #1 | P: TB | "Mycobacterium tuberculosis"[MeSH] OR "Tuberculosis"[MeSH] OR Tuberculo*[tiab] OR TB[tiab] |
| #2 | I: protein, cytokine | "Cytokines/blood"[MeSH] OR "Blood Proteins"[MeSH] OR "Biomarkers/blood"[MeSH] OR proteom*[tiab] OR cytokin*[tiab] OR CRP[tiab] OR "C-reactive protein"[tiab] OR "IP-10"[tiab] |
| #3 | I: blood | blood[tw] OR plasma[tw] OR serum[tw] |
| #4 | O: diagnosis | "diagnosis"[MeSH] OR "Sensitivity and Specificity"[MeSH] OR diagnos*[tiab] OR detect*[tiab] OR discriminat*[tiab] |
| #5 | Date | "2010/01/01"[PDat "2023/10/05"[PDat] |
| #6 |  | (#1) AND (#2) AND (#3) AND (#4) AND (#5) |

**Table S2:**
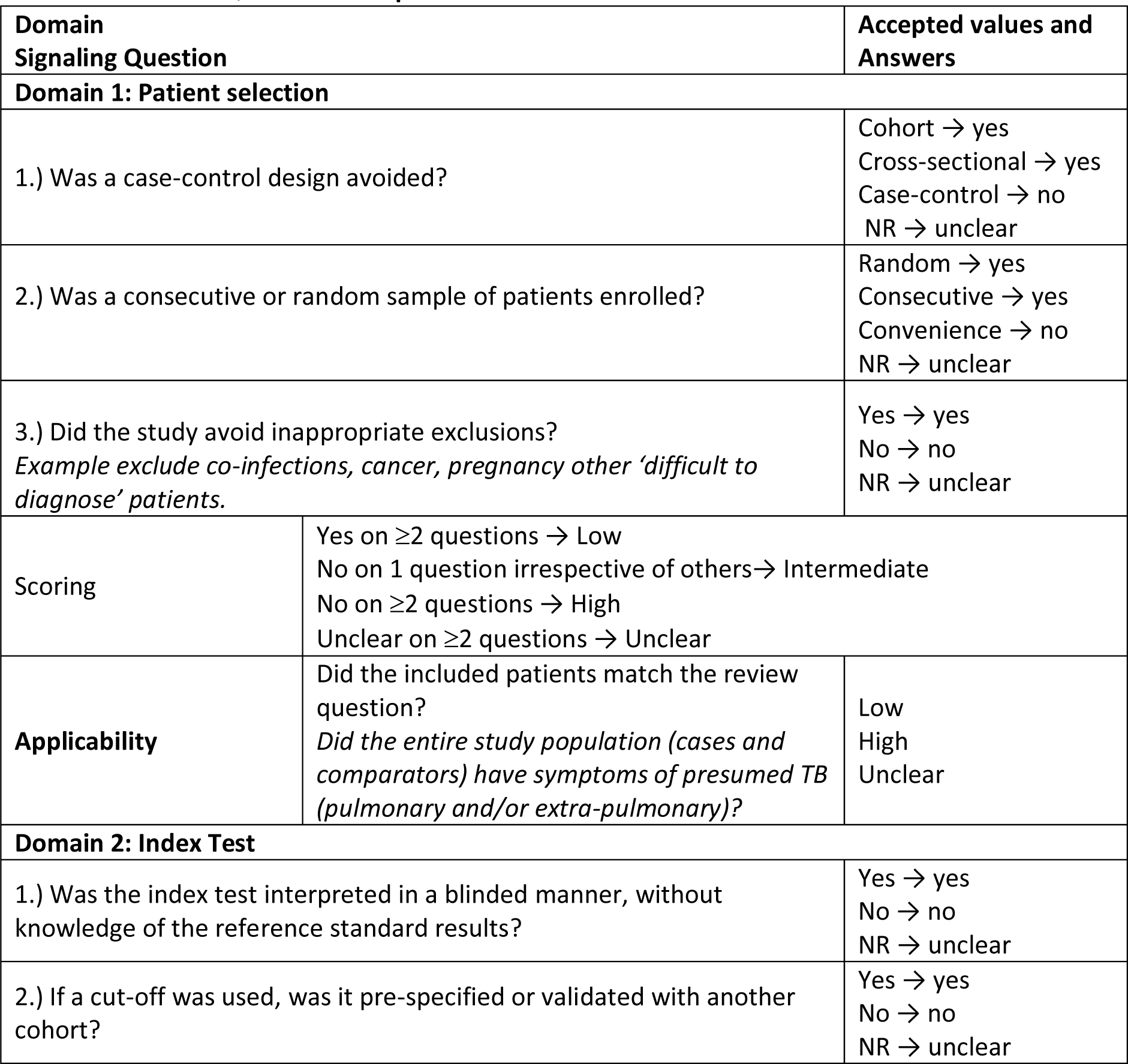
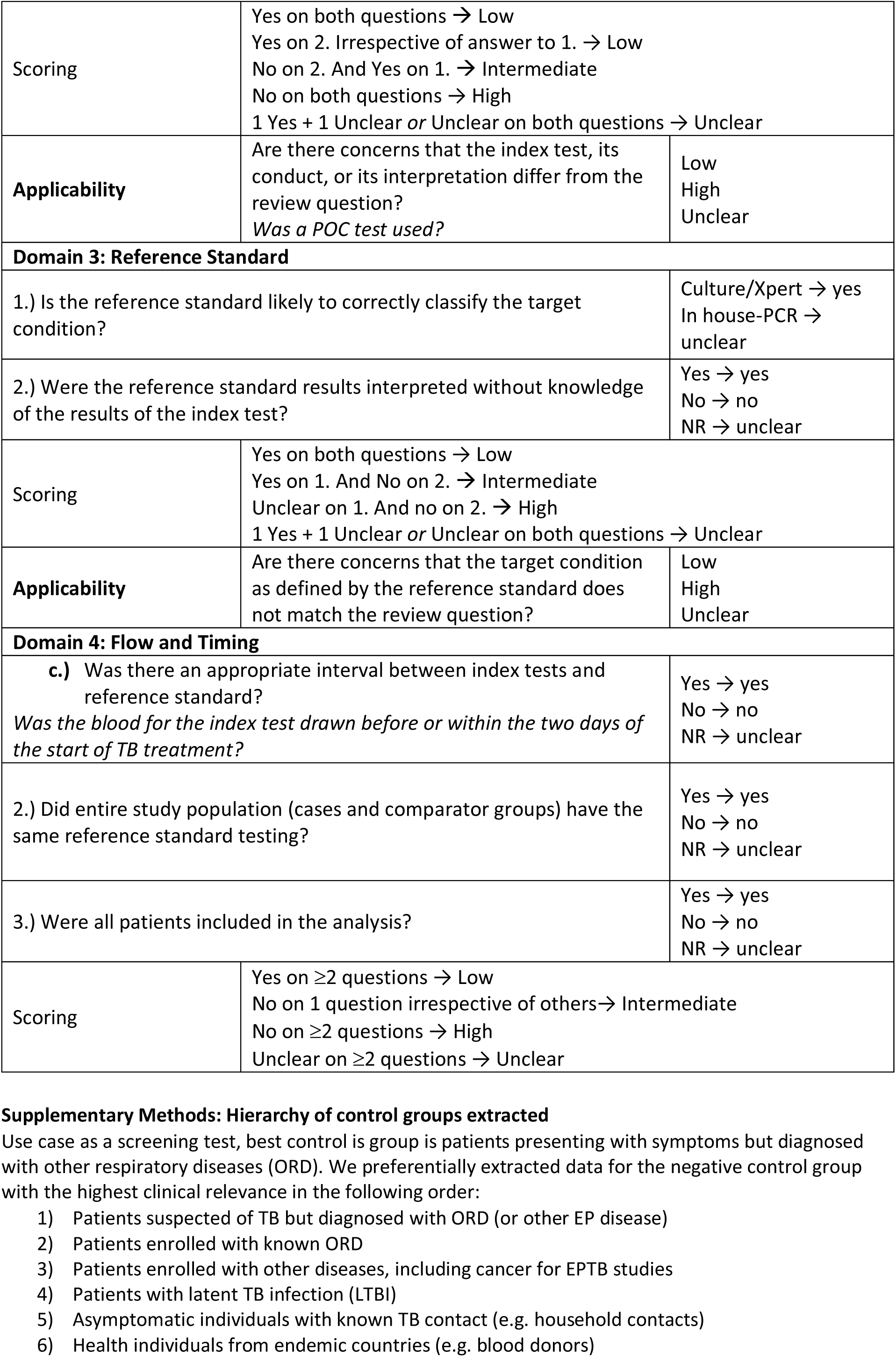
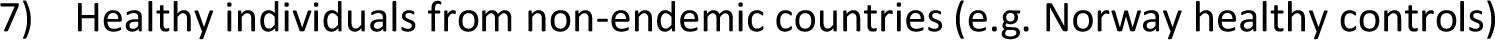
Modified QUADAS-2 template.

**Table S3:**
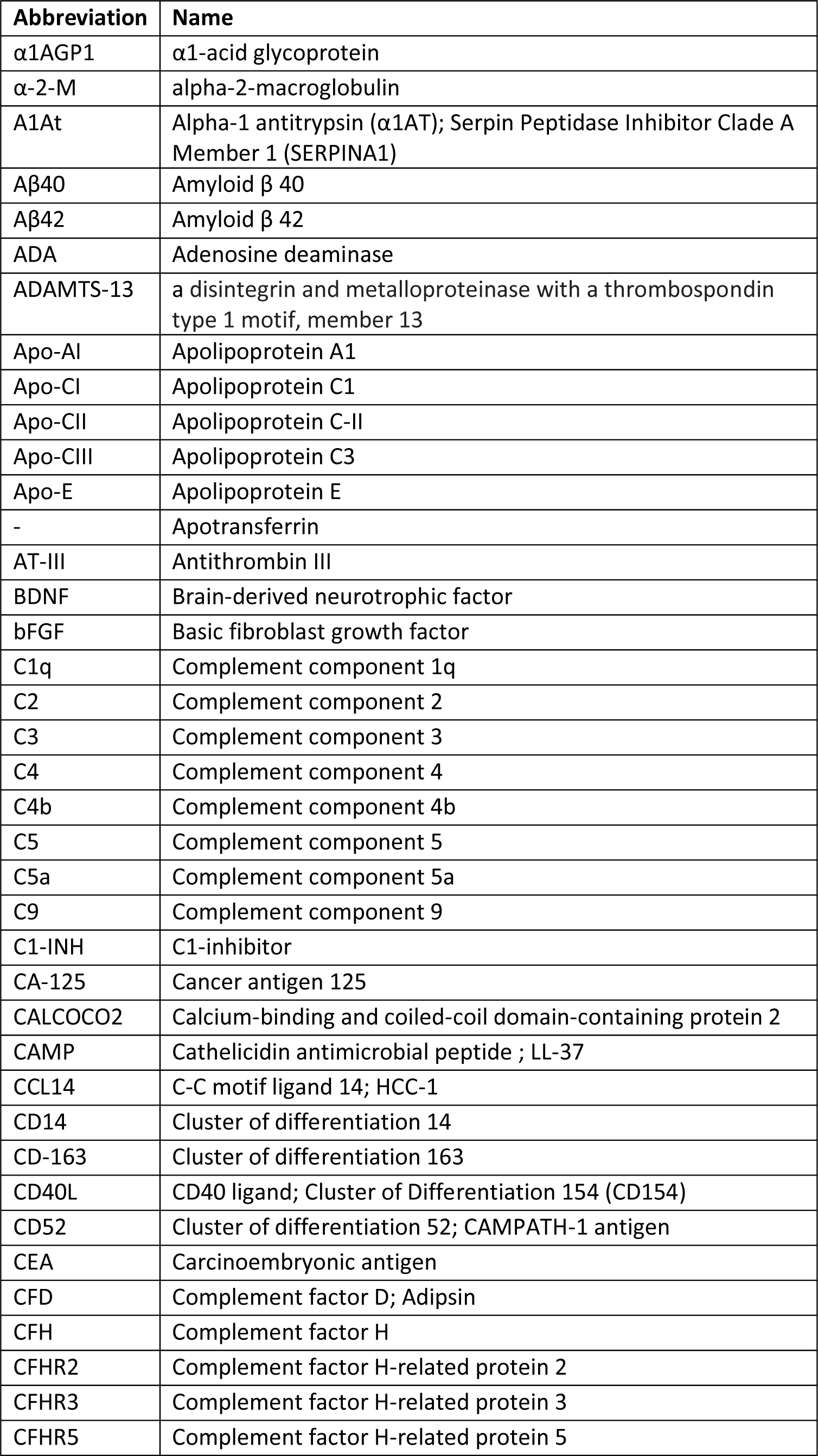
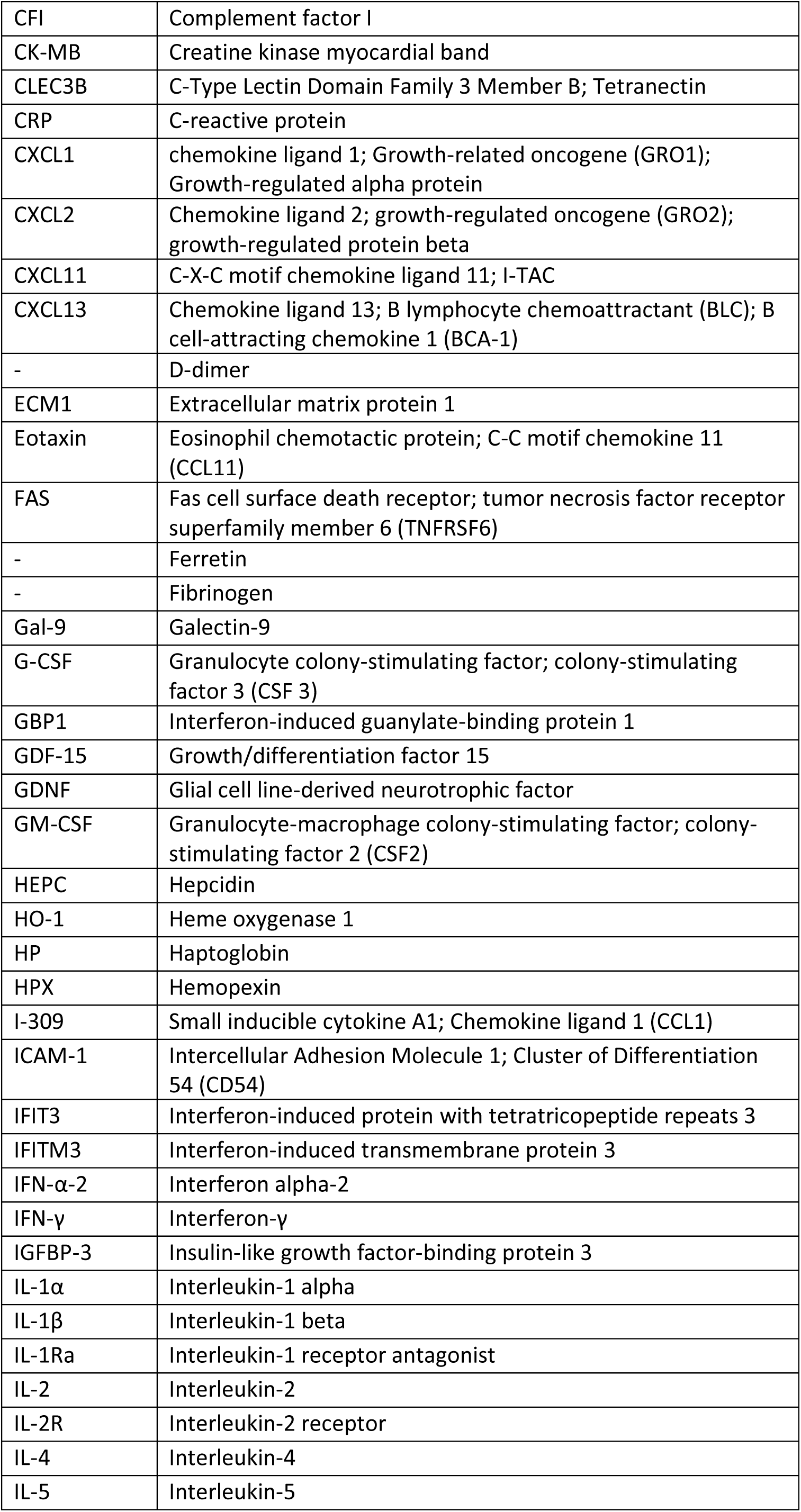
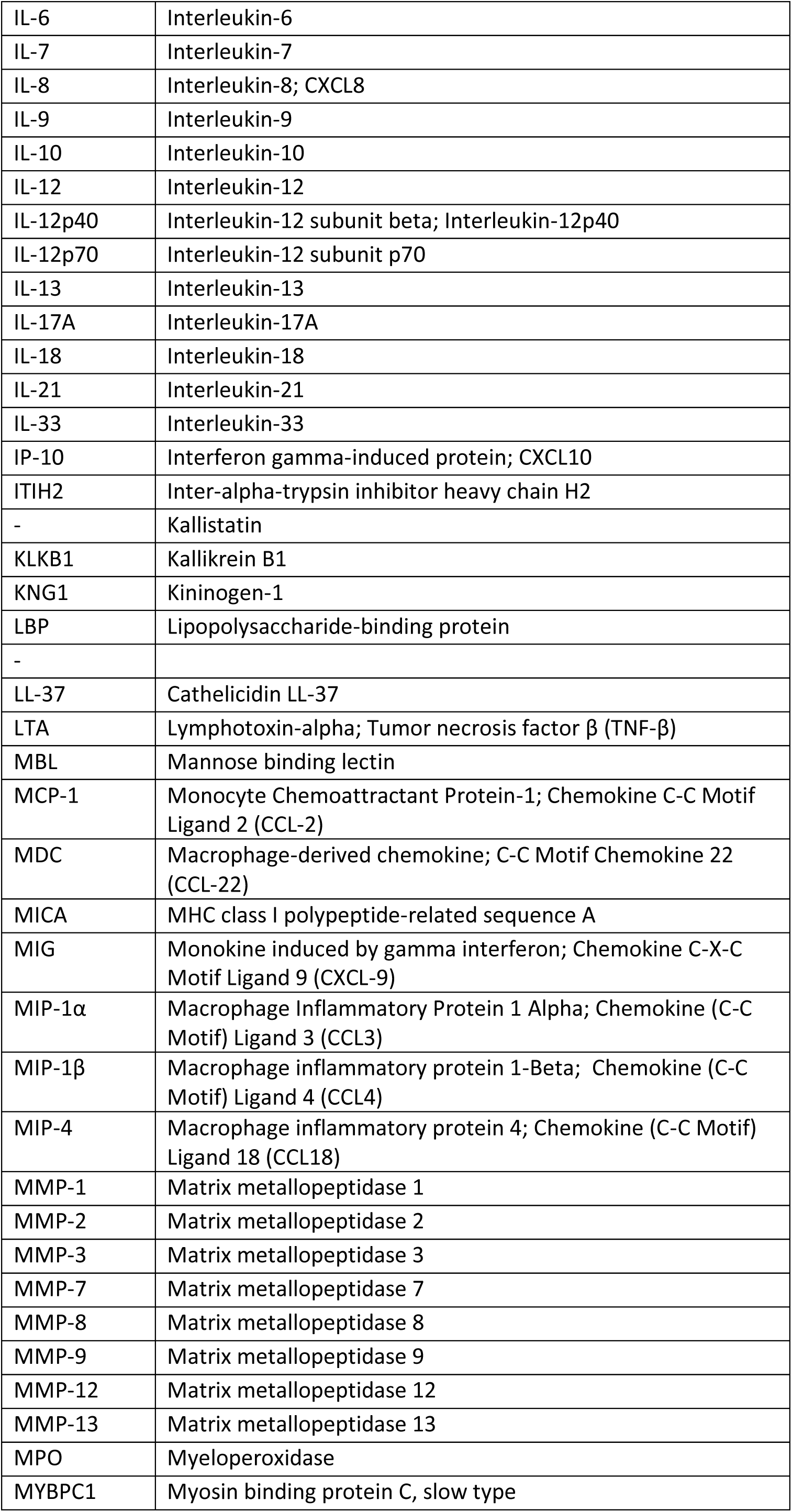
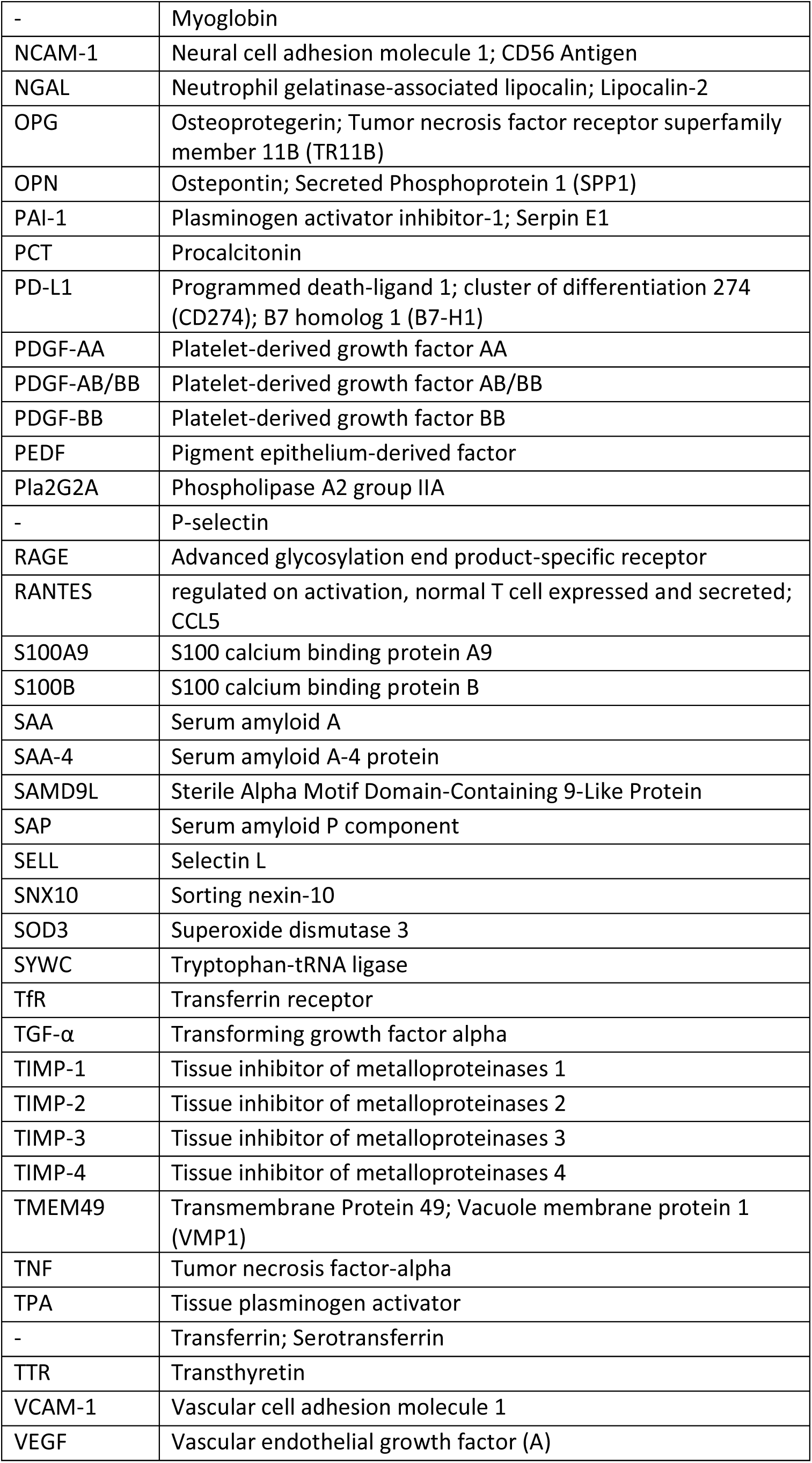
Biomarker abbreviations.

**Table S4:**
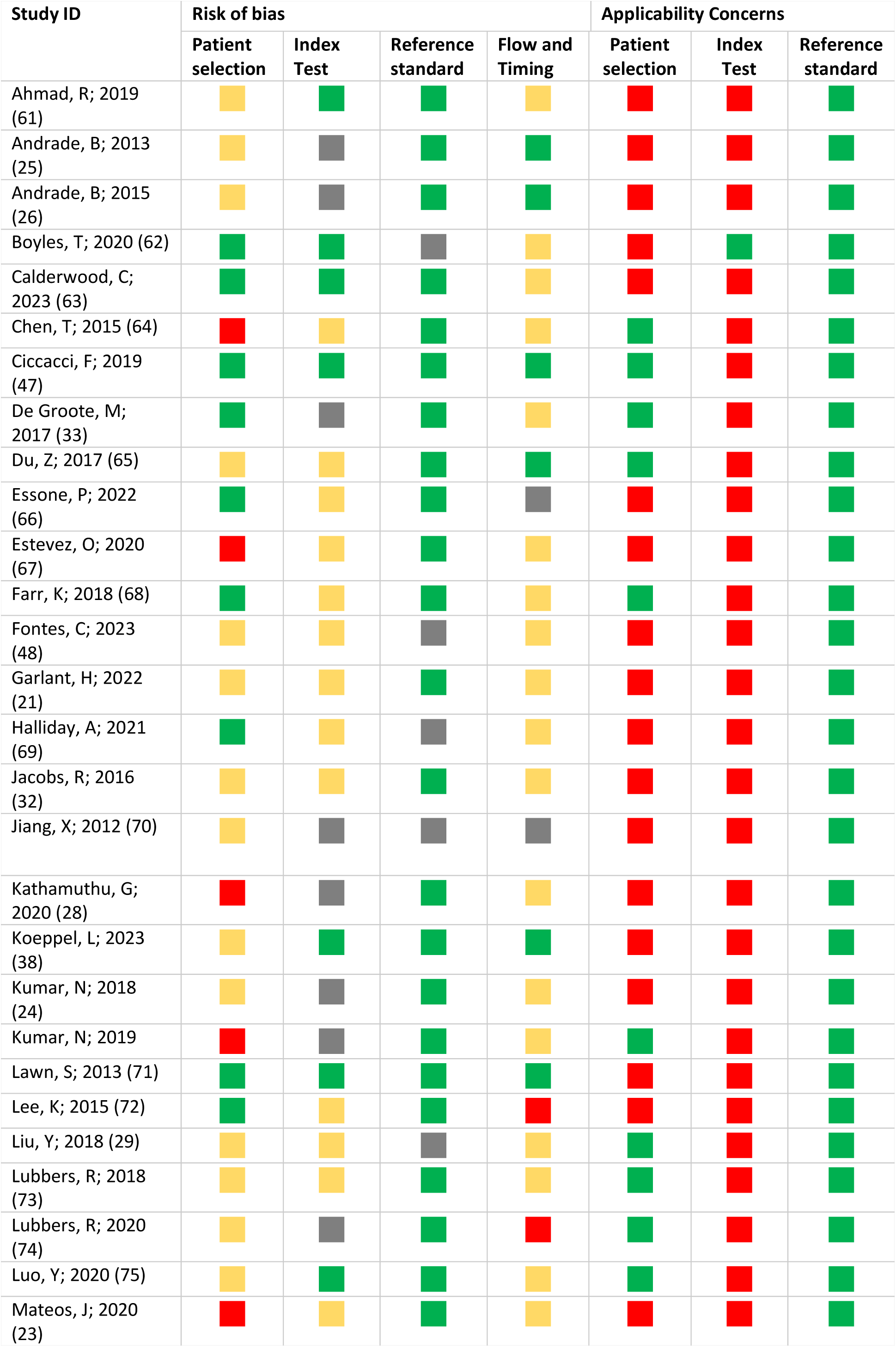
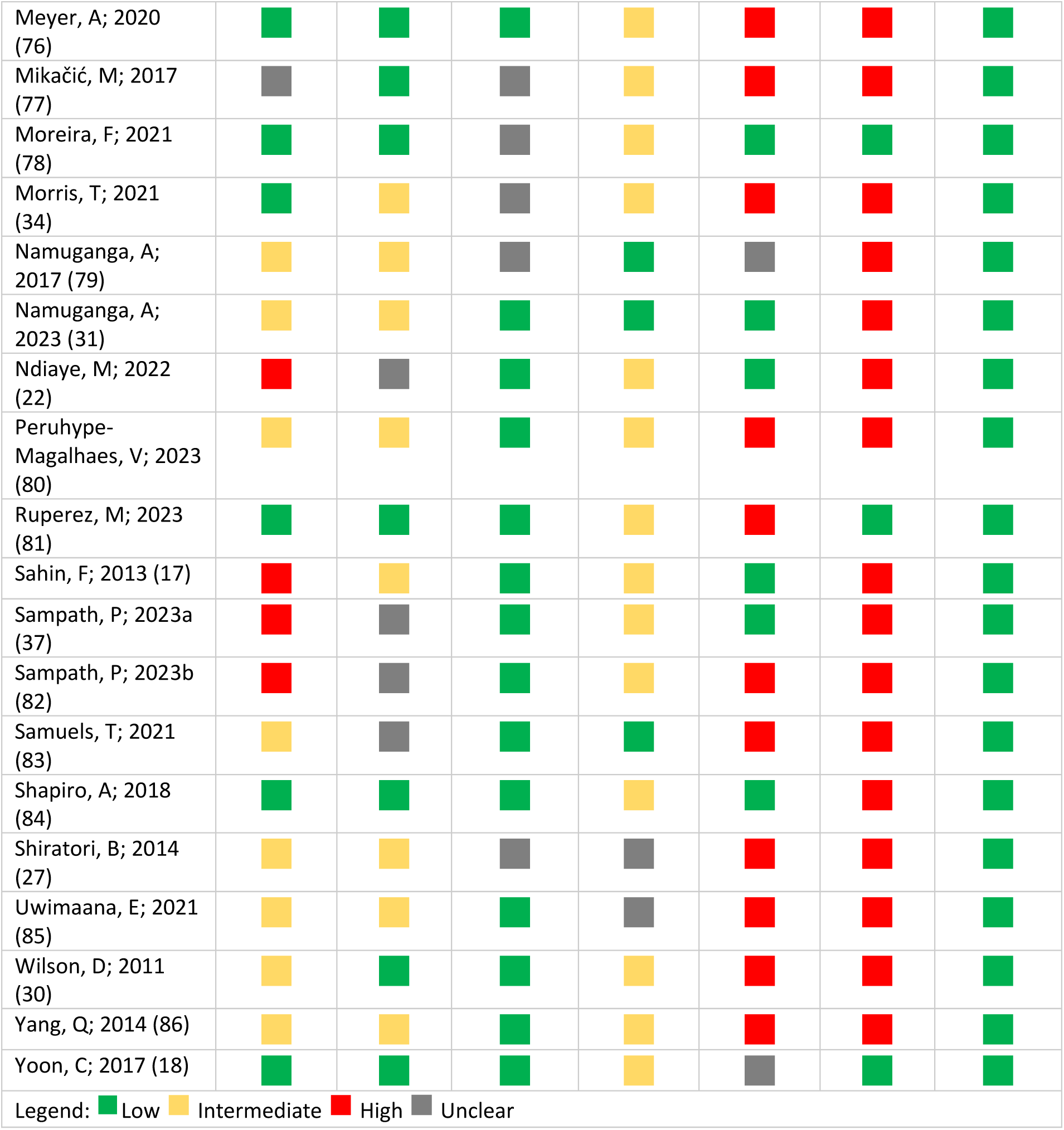
QUADAS-2 detailed results by paper, adult pulmonary TB.

**Figure S1.**
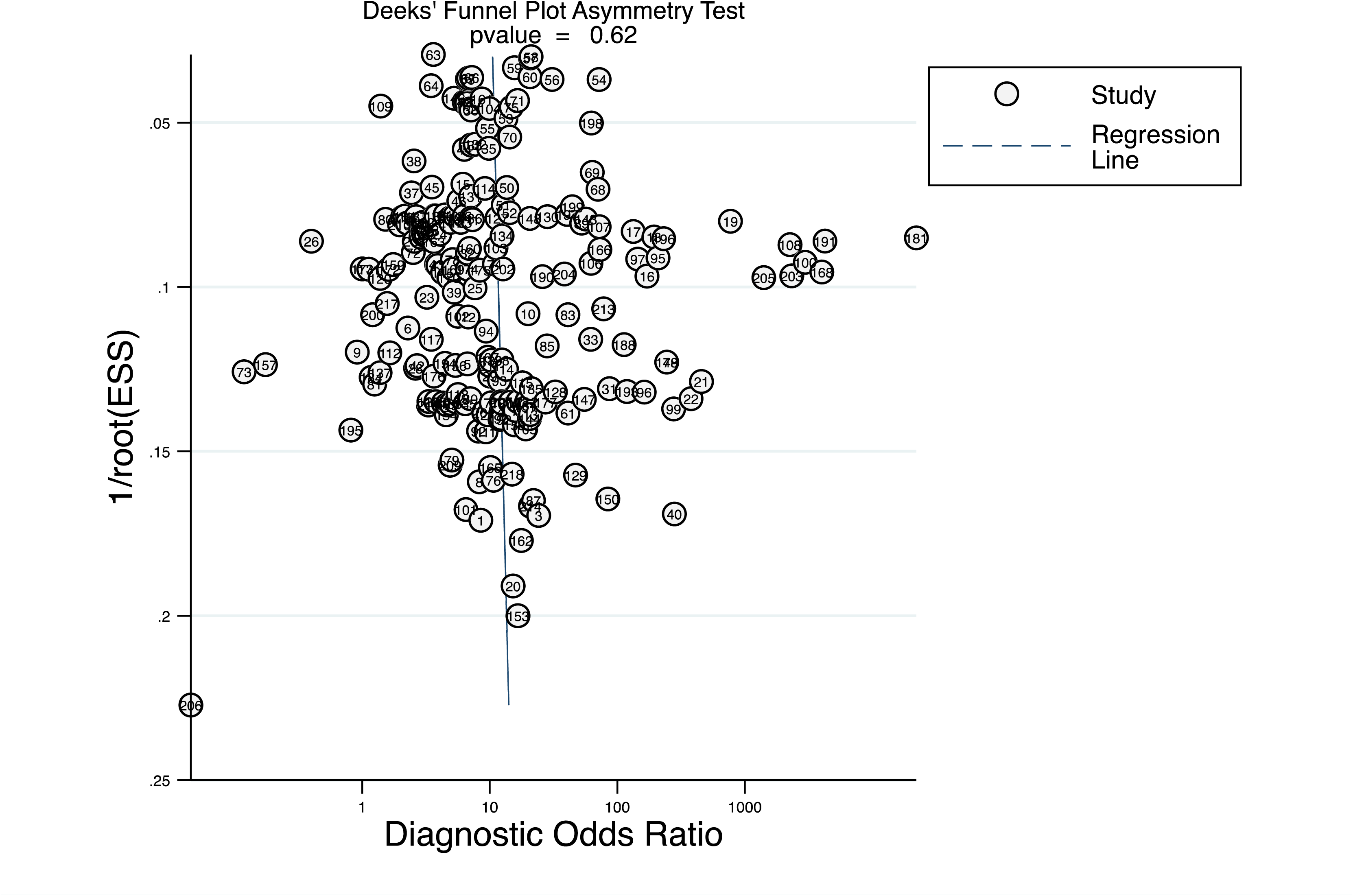
Funnel plot for publication bias, all adult pulmonary TB studies.

**Table S5.**
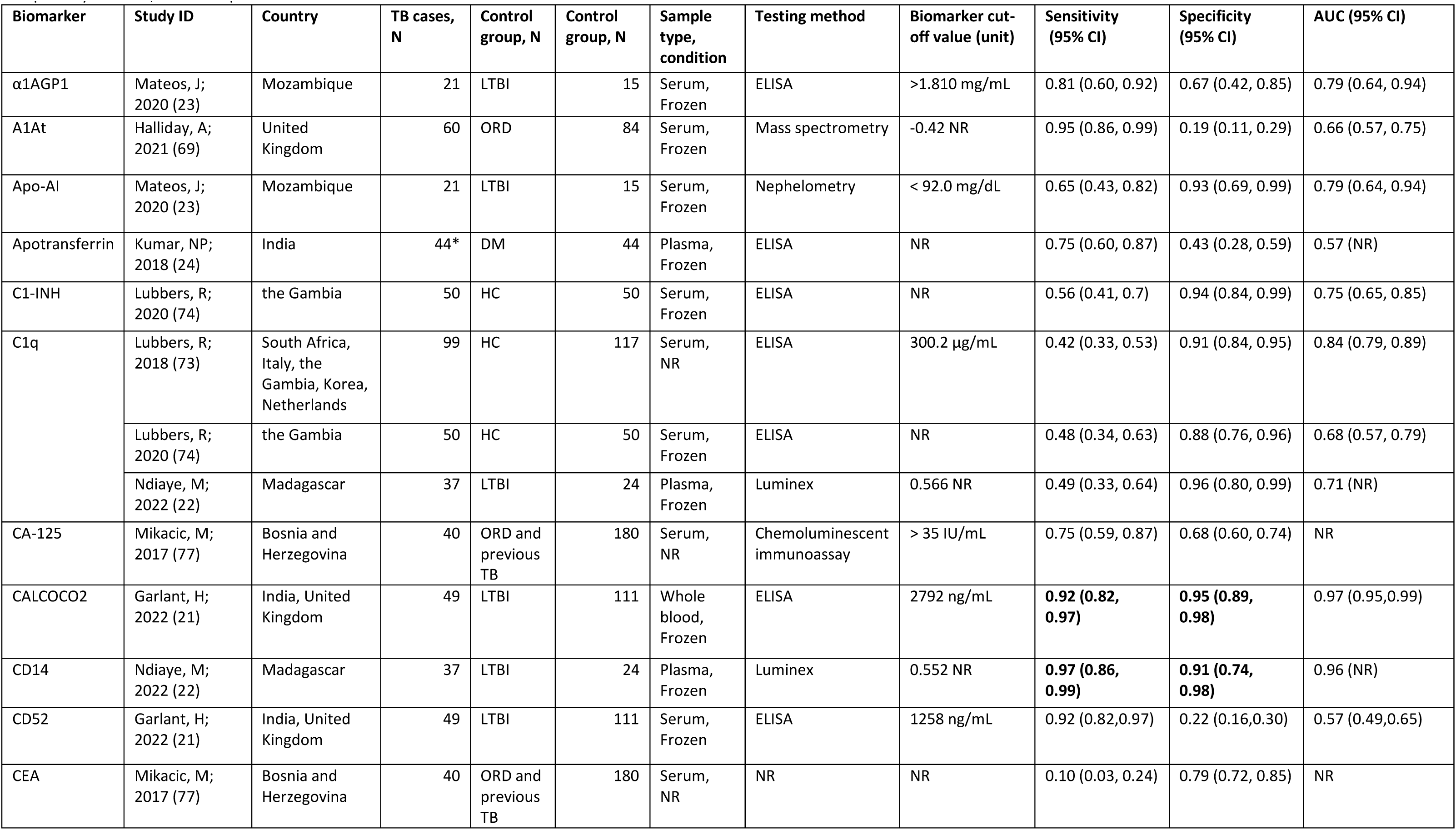
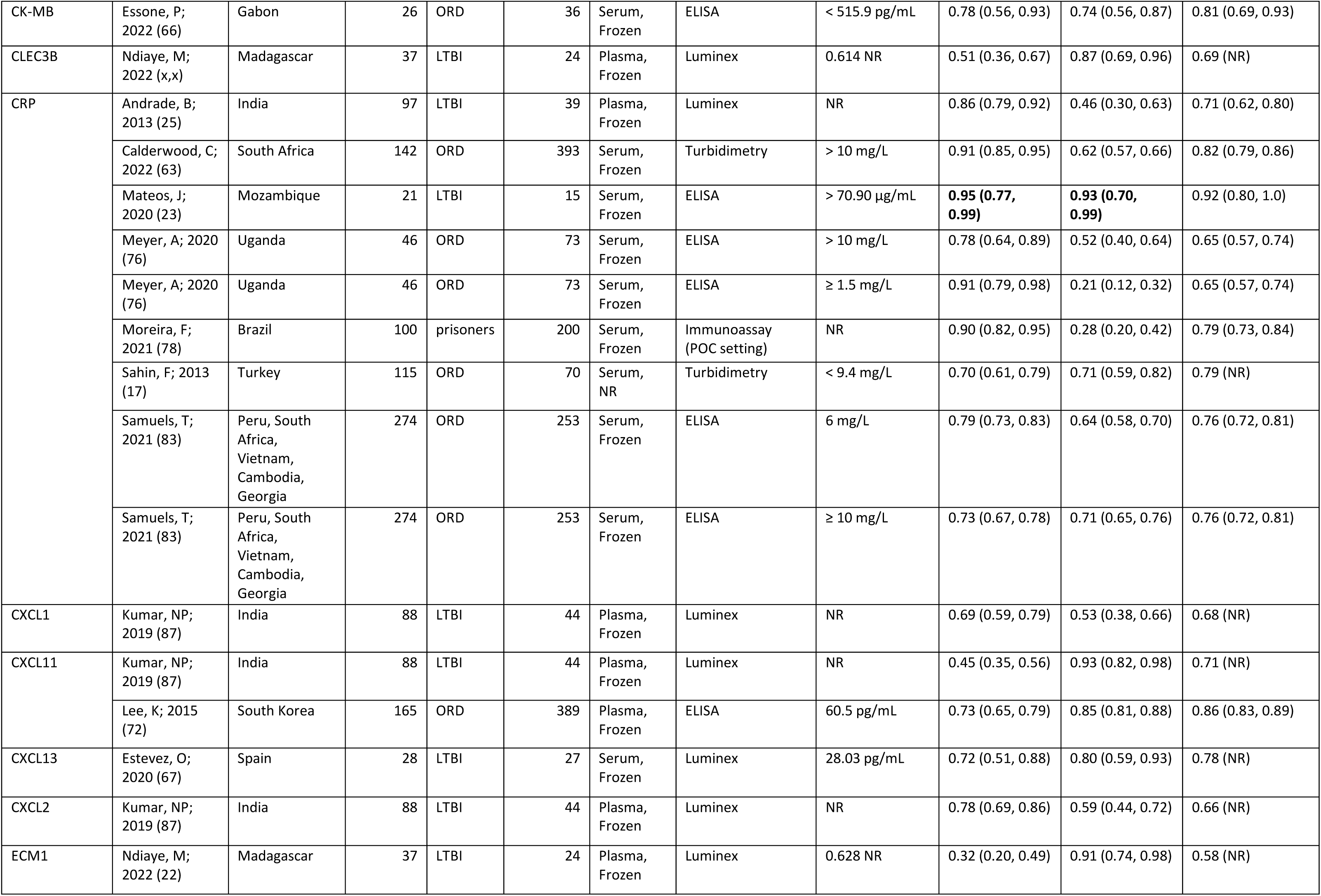
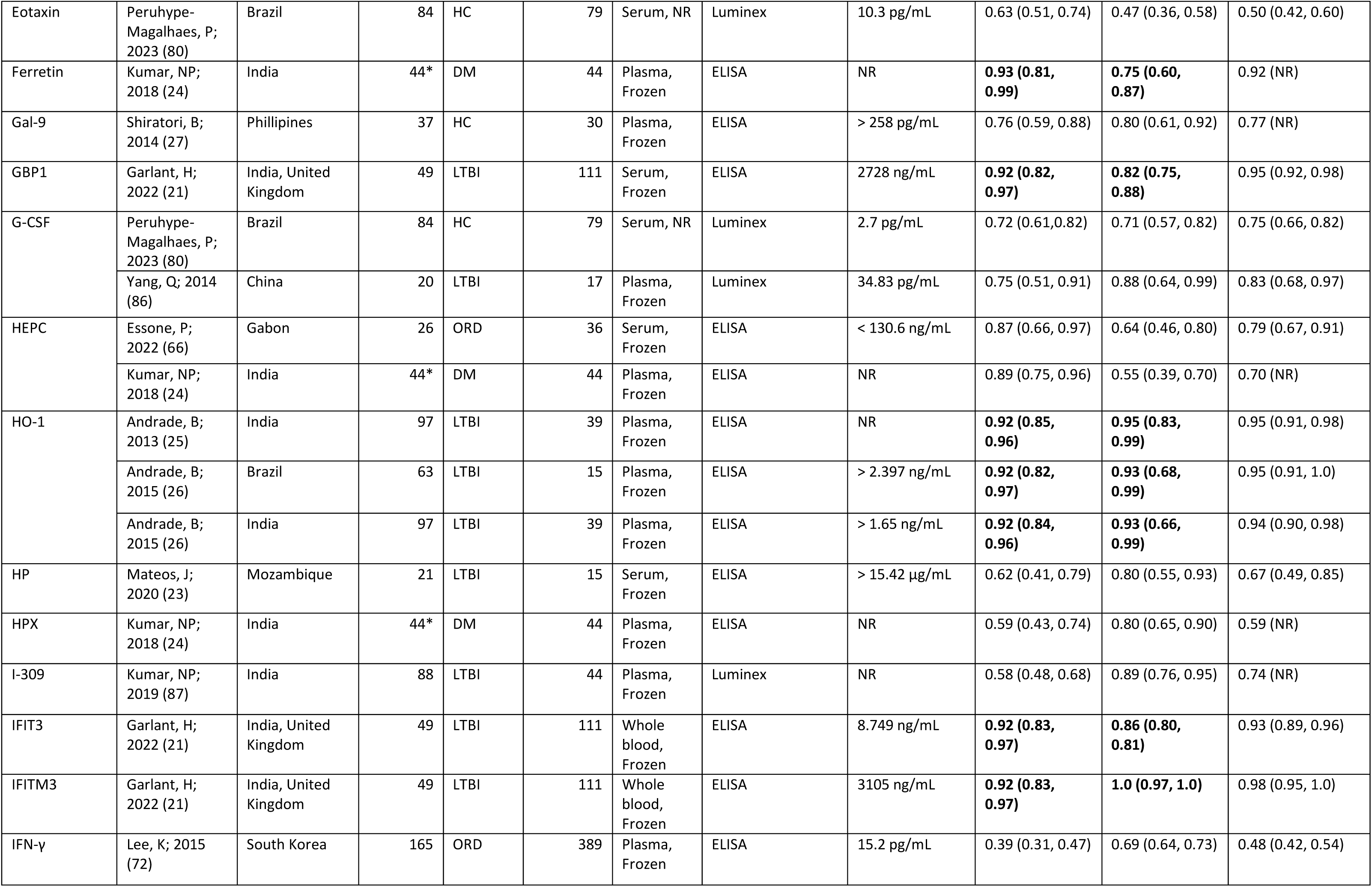
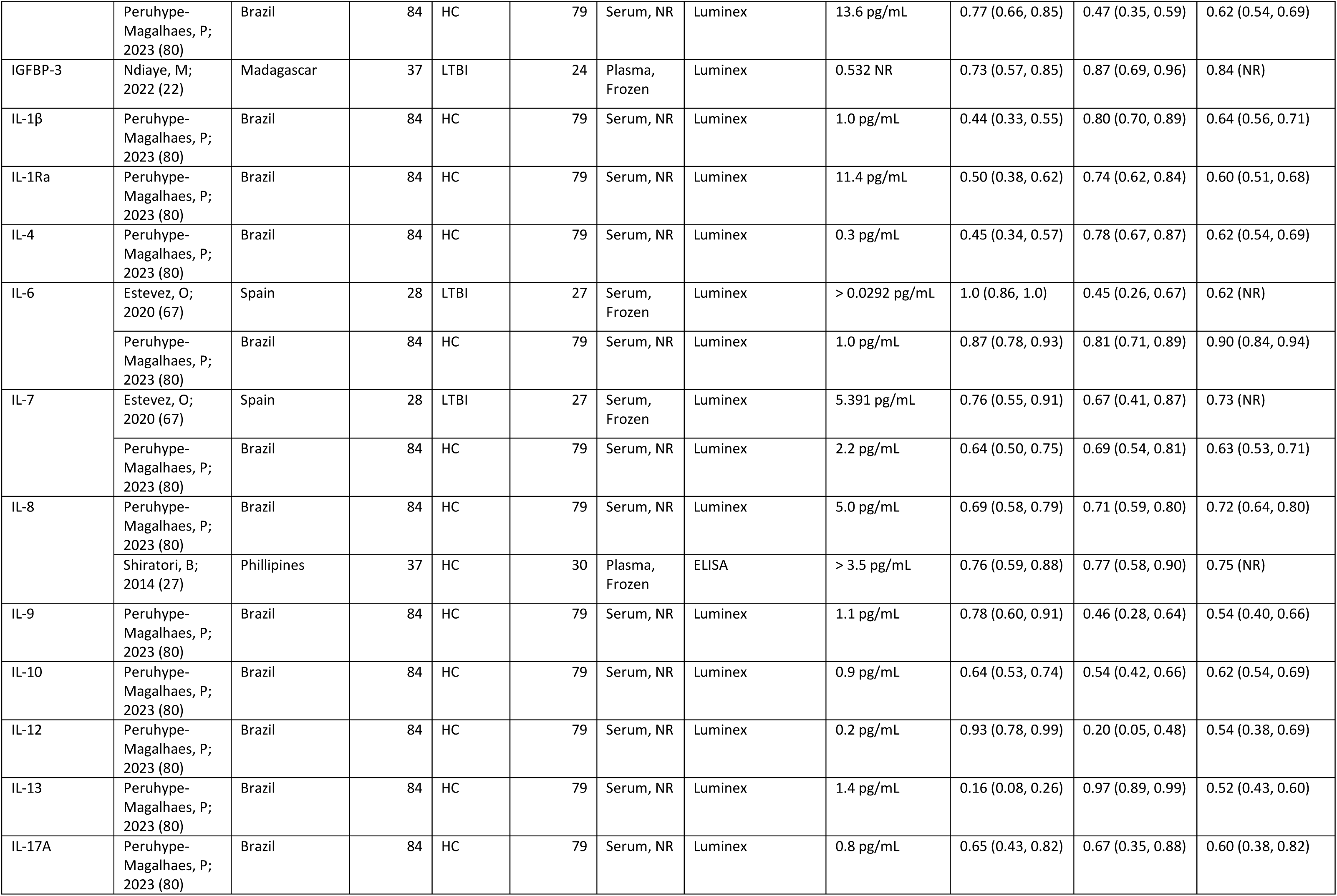
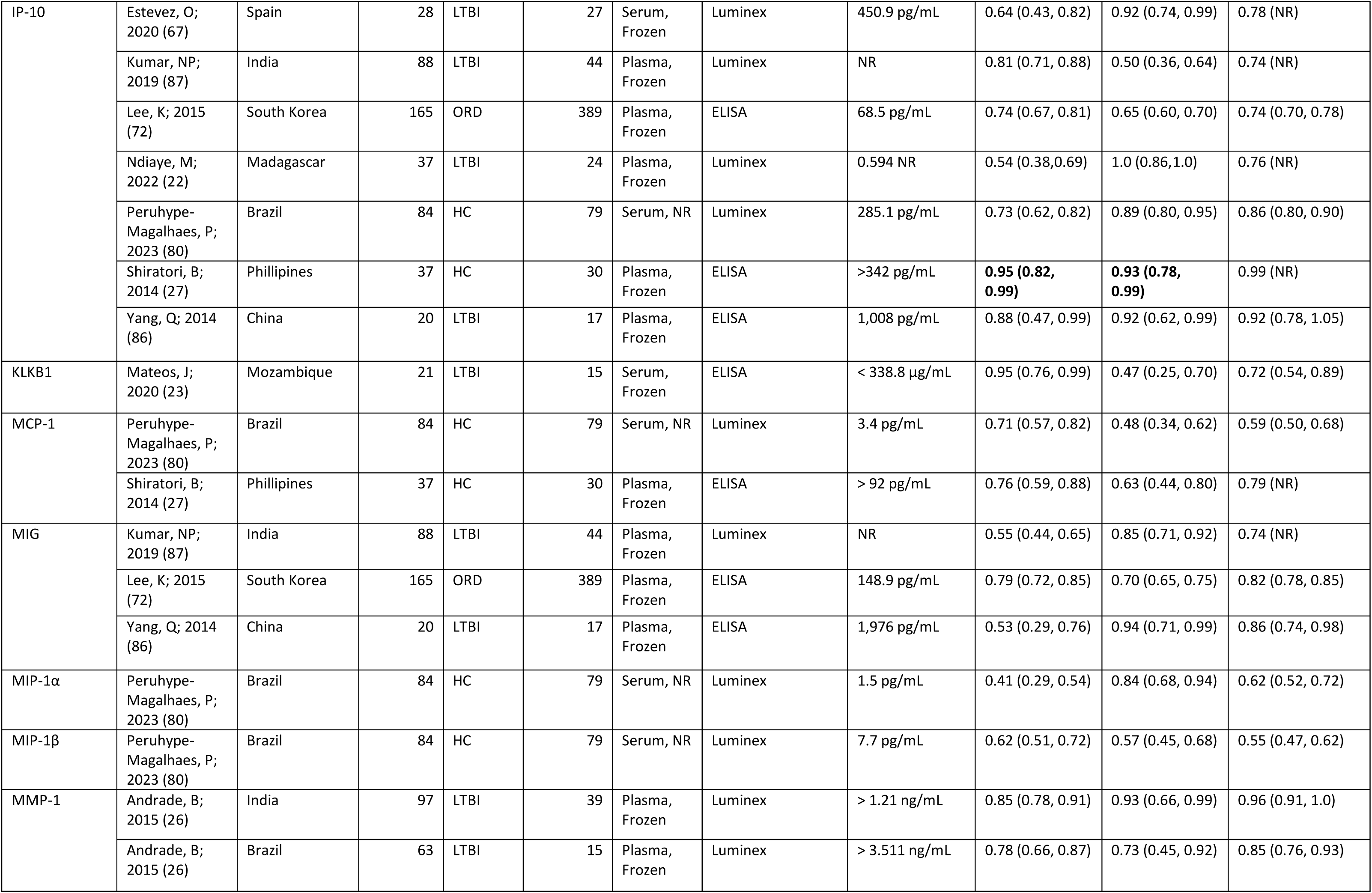
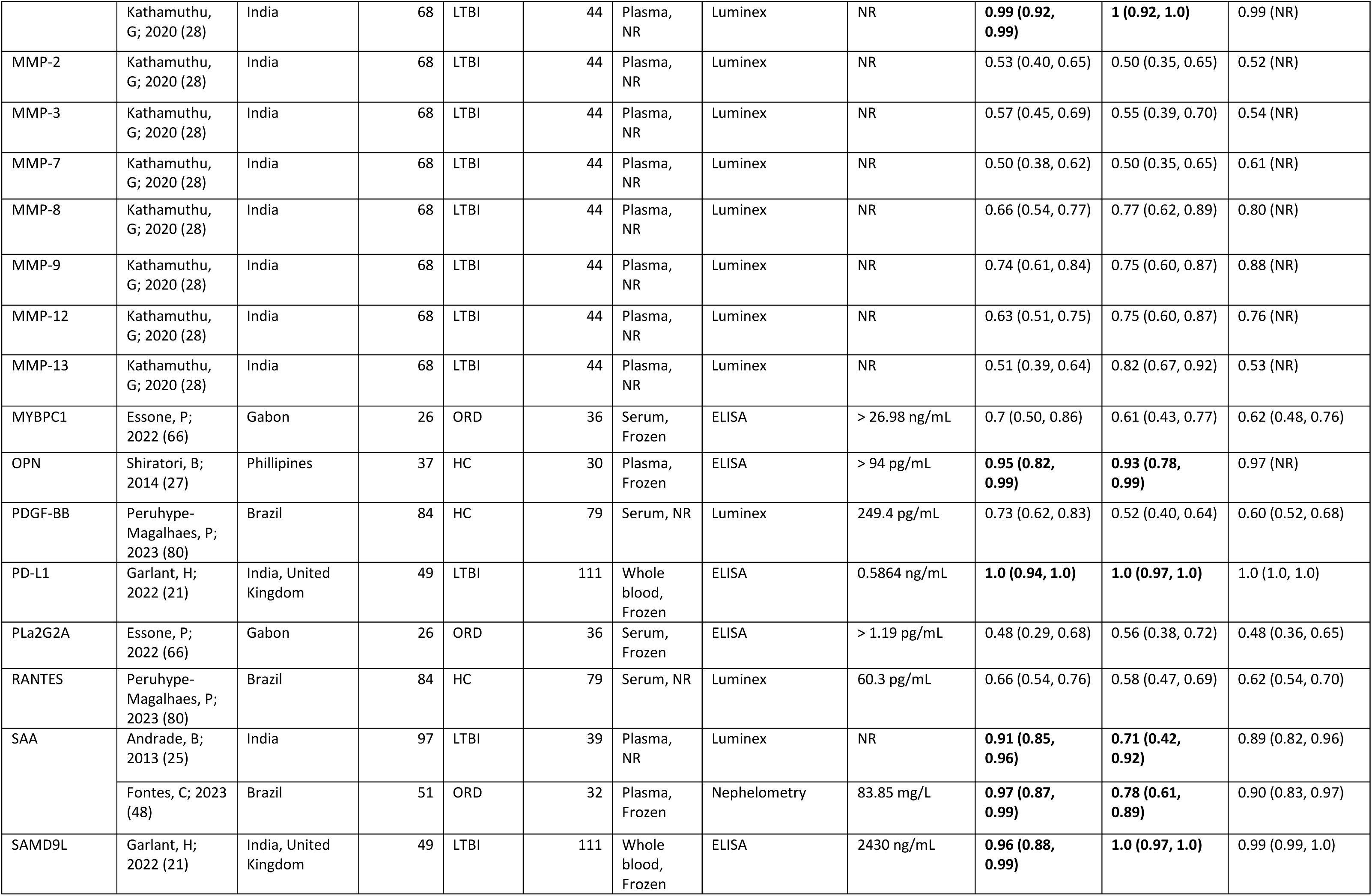
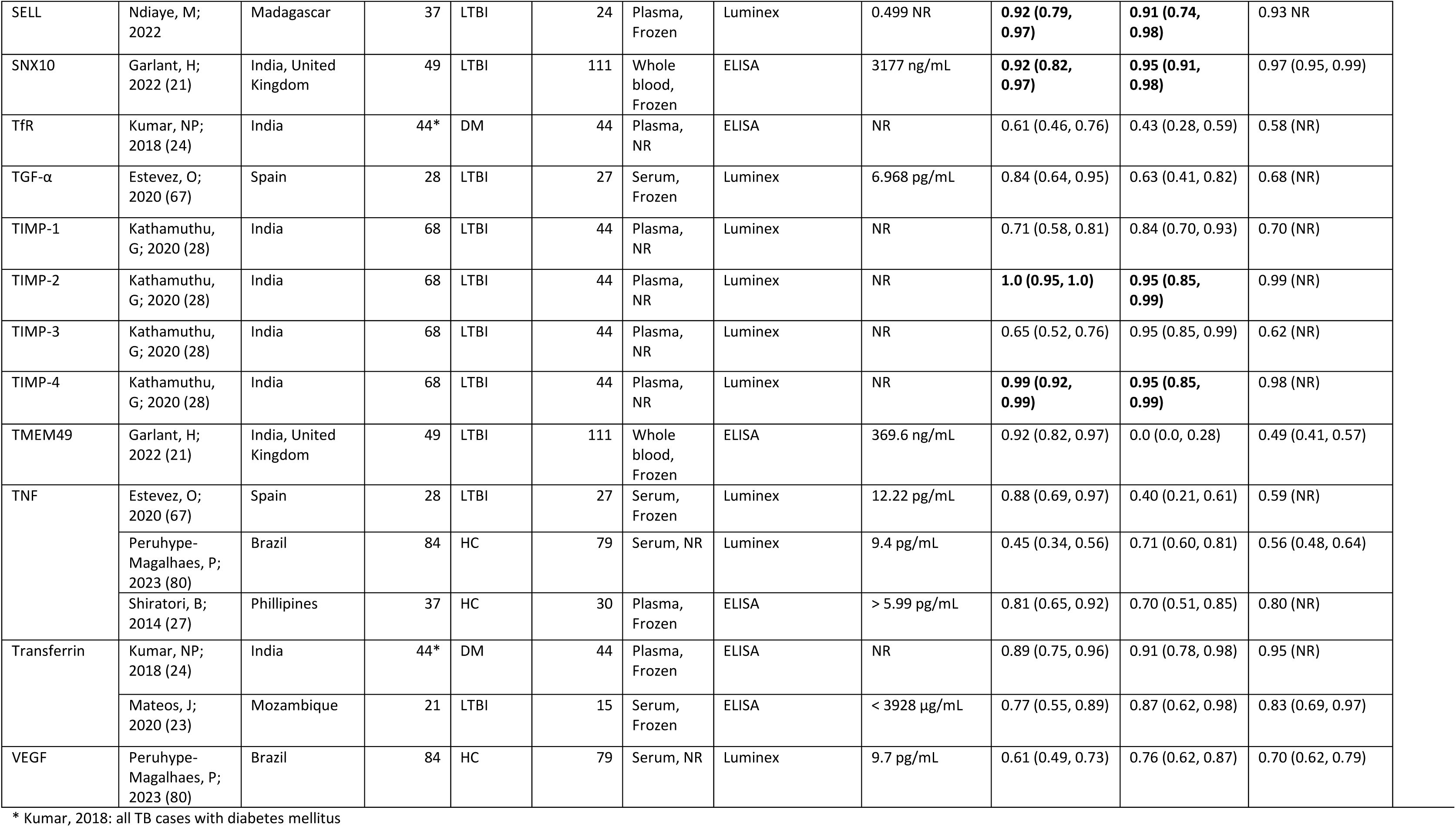
Biomarkers in adults, pulmonary TB, HIV-negative. Biomarkers meeting TPP criteria are indicated in bold AUC=area under the curve, DM=diabetes mellitus, ELISA=enzyme-linked immunosorbent assay, HC= healthy controls, LTBI=latent TB infection, MSD=Meso Scale Discovery, N=sample size, ORD=other respiratory diseases, NR=not reported

**Table S6.** Biomarkers in adults, pulmonary TB, HIV-positive. Biomarkers meeting TPP criteria are indicated in bold AUC=area under the curve, DM=diabetes mellitus, ELISA=enzyme-linked immunosorbent assay, HC= healthy controls, LTBI=latent TB infection, MSD=Meso Scale Discovery, N=sample size, ORD=other respiratory diseases, NR=not reported

| Biomarker | Study ID | Country | TB cases, N | Control group | Control group, N | Sample type, condition | Testing method | Biomarker cut-off value (unit) | Sensitivity (95% CI) | Specificity (95% CI) | AUC (95% CI) |
| --- | --- | --- | --- | --- | --- | --- | --- | --- | --- | --- | --- |
| CD14 | Liu, Y; 2018 (29) | South Africa | 39 | HC | 24 | Serum, Frozen | ELISA | 1540 ng/mL | **0.95 (0.83, 0.99)** | **0.96 (0.79, 0.99)** | NR |
| CRP | Boyles, T; 2020 (62) | South Africa | 75 | ORD | 132 | NR, Fresh | Turbidimetry (POC setting) | ≥10 mg/L | 0.95 (0.87, 0.98) | 0.26 (0.19, 0.34) | NR |
|  | Calderwood, C; 2022 (63) | South Africa | 113 | ORD | 276 | Serum, Frozen | Turbidimetry | ≥10 mg/L | 0.95 (0.89, 0.98) | 0.43 (0.37, 0.49) | 0.77 (0.73, 0.82) |
|  | Ciccacci, F; 2019 (47) | Mozambique | 21 | pre-ART screening | 122 | Plasma, Frozen | ELISA | > 10 mg/L | 0.76 (0.55, 0.89) | 0.95 (0.90, 0.98) | NR |
|  | Farr, K; 2018 (68) | Uganda | 155 | ORD | 107 | Plasma, Frozen | Turbidimetry | NR | 0.90 (0.85, 0.95) | 0.33 (0.30, 0.36) | NR |
|  | Lawn, S; 2013 (71) | South Africa | 81 | pre-ART screening | 415 | Serum, Frozen | ELISA | ≥ 5 mg/l | 0.90 (0.81, 0.95) | 0.44 (0.39, 0.49) | 0.81 (NR) |
|  | Ruperez, M; 2023 (81) | South Africa, Zambia | 25 | ORD | 251 | Whole blood, Fresh | Immunoassay (POC setting) | ≥10 mg/L | 0.40 (0.21, 0.61) | 0.79 (0.73, 0.83) | 0.59 (0.49, 0.69) |
|  | Ruperez, M; 2023 (81) | South Africa, Zambia | 25 | ORD | 251 | Whole blood, Fresh | Immunoassay (POC setting) | ≥5 mg/L | 0.60 (0.39, 0.79) | 0.63 (0.57, 0.69) | 0.61 (0.51, 0.72) |
|  | Samuels, T; 2021 (83) | Peru, South Africa, Vietnam, Cambodia, Georgia | 111 | ORD | 102 | Serum, Frozen | ELISA | ≥ 10 mg/L | 0.92 (0.85, 0.96) | 0.53 (0.43, 0.62) | 0.83 (0.78, 0.89) |
|  | Shapiro, A; 2018 (84) | South Africa | 42 | pre-ART screening | 383 | Serum, Frozen | Turbidimetry | > 5 mg/L | 0.91 (0.77, 0.97) | 0.59 (0.53, 0.64) | 0.80 (0.72, 0.88) |
|  | Shapiro, A; 2018 (84) | South Africa | 42 | pre-ART screening | 383 | Serum, Frozen | Turbidimetry | > 10 mg/L | 0.79 (0.63, 0.90) | 0.72 (0.68, 0.77) | 0.80 (0.72, 0.88) |
|  | Yoon, C; 2017 (18) | Uganda | 163 | pre-ART screening | 1014 | Whole blood, Fresh | Immunoassay (POC setting) | ≥ 8 mg/L | **0.90 (0.85, 0.94)** | **0.70 (0.67, 0.72)** | 0.81 (0.78, 0.83) |
|  | Yoon, C; 2017 (18) | Uganda | 163 | pre-ART screening | 1014 | Whole blood, Fresh | Immunoassay (POC setting) | ≥ 10 mg/L | 0.89 (0.83, 0.93) | 0.72 (0.69, 0.75) | 0.81 (0.78, 0.83) |
| CXCL1 | Farr, K; 2018 (68) | Uganda | 155 | ORD | 107 | Plasma, Frozen | Luminex | NR | 0.90 (0.85, 0.95) | 0.01 (0.00, 0.03) | NR |
| HO-1 | Uwimaana, E; 2021 (85) | Uganda | 70 | LTBI | 70 | Plasma, Frozen | ELISA | > 8.95 ng/mL | 0.59 (0.47, 0.69) | 0.67 (0.56, 0.77) | 0.57 (0.47, 0.66) |
| IFN-γ | Farr, K; 2018 (68) | Uganda | 155 | ORD | 107 | Plasma, Frozen | Luminex | NR | 0.90 (0.85, 0.95) | 0.27 (0.22, 0.32) | NR |
| IL-6 | Farr, K; 2018 (68) | Uganda | 155 | ORD | 107 | Plasma, Frozen | Luminex | NR | 0.90 (0.85, 0.95) | 0.44 (0.37, 0.50) | NR |
| IL-18 | Farr, K; 2018 (68) | Uganda | 155 | ORD | 107 | Plasma, Frozen | Luminex | NR | 0.90 (0.85, 0.95) | 0.13 (0.08, 0.19) | NR |
| MDC | Farr, K; 2018 (68) | Uganda | 155 | ORD | 107 | Plasma, Frozen | Luminex | NR | 0.90 (0.85, 0.95) | 0.02 (0.00, 0.04) | NR |
| MIG | Farr, K; 2018 (68) | Uganda | 155 | ORD | 107 | Plasma, Frozen | Luminex | NR | 0.90 (0.85, 0.95) | 0.16 (0.11, 0.21) | NR |

**Table S7.**
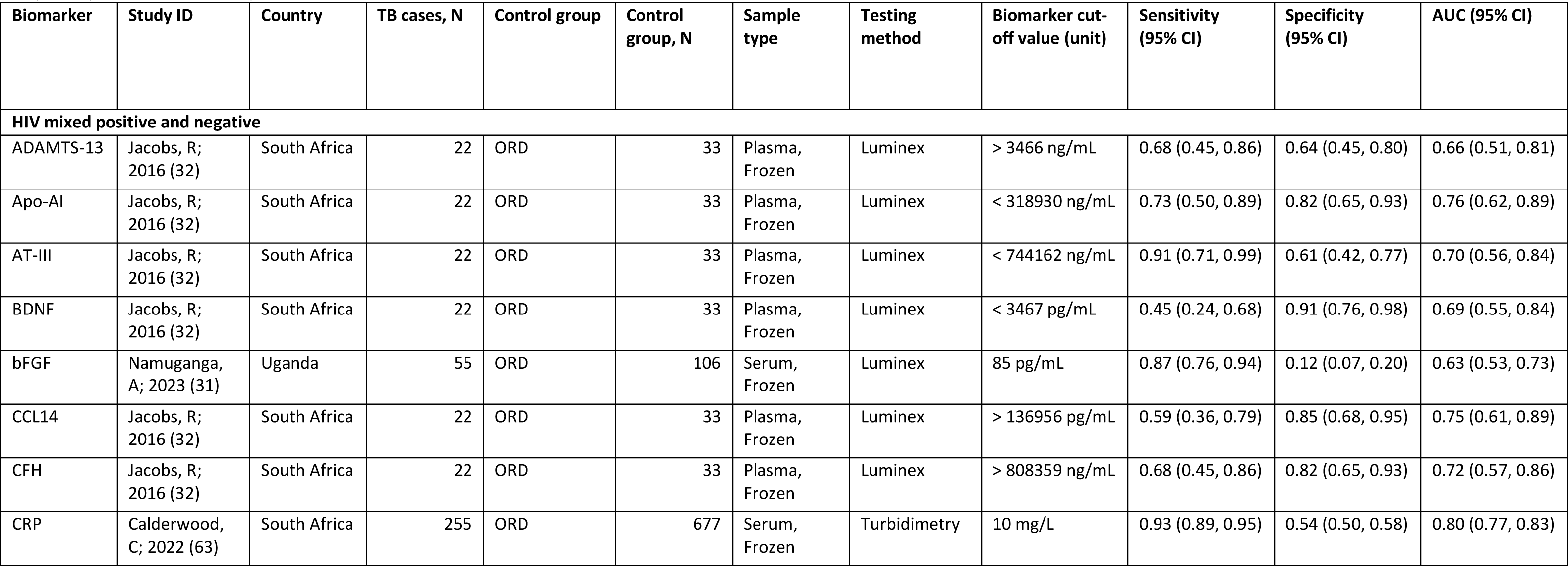
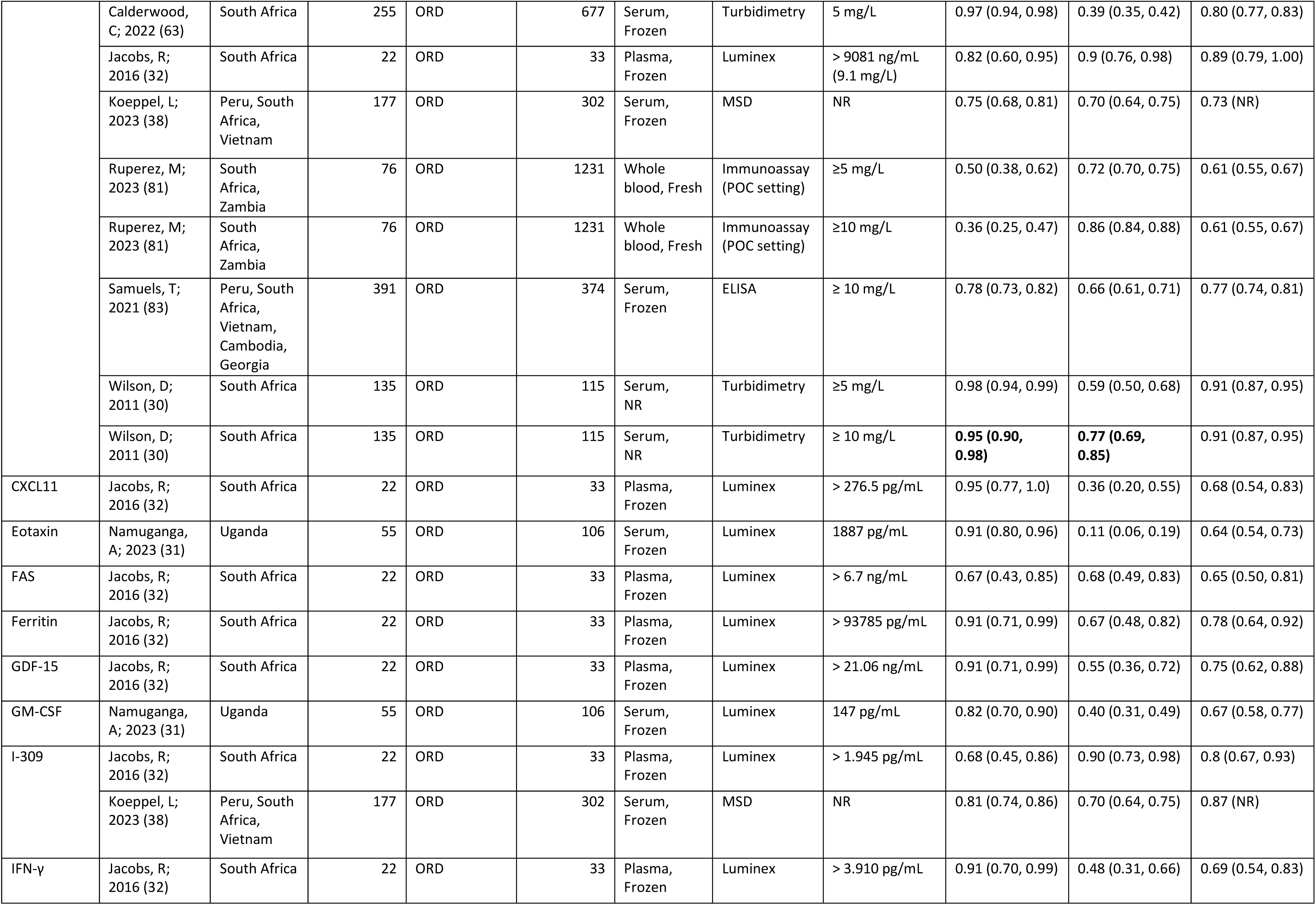
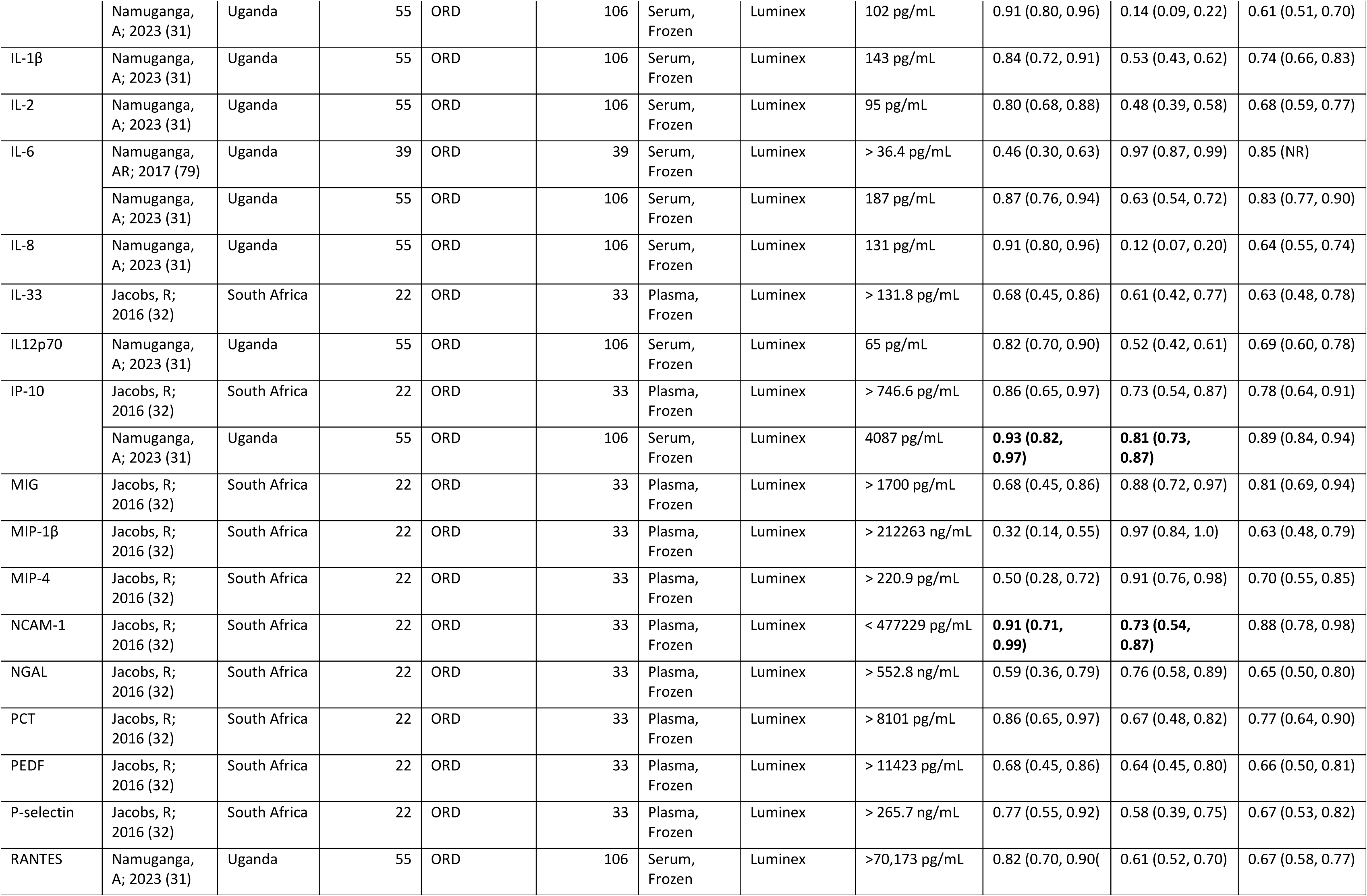
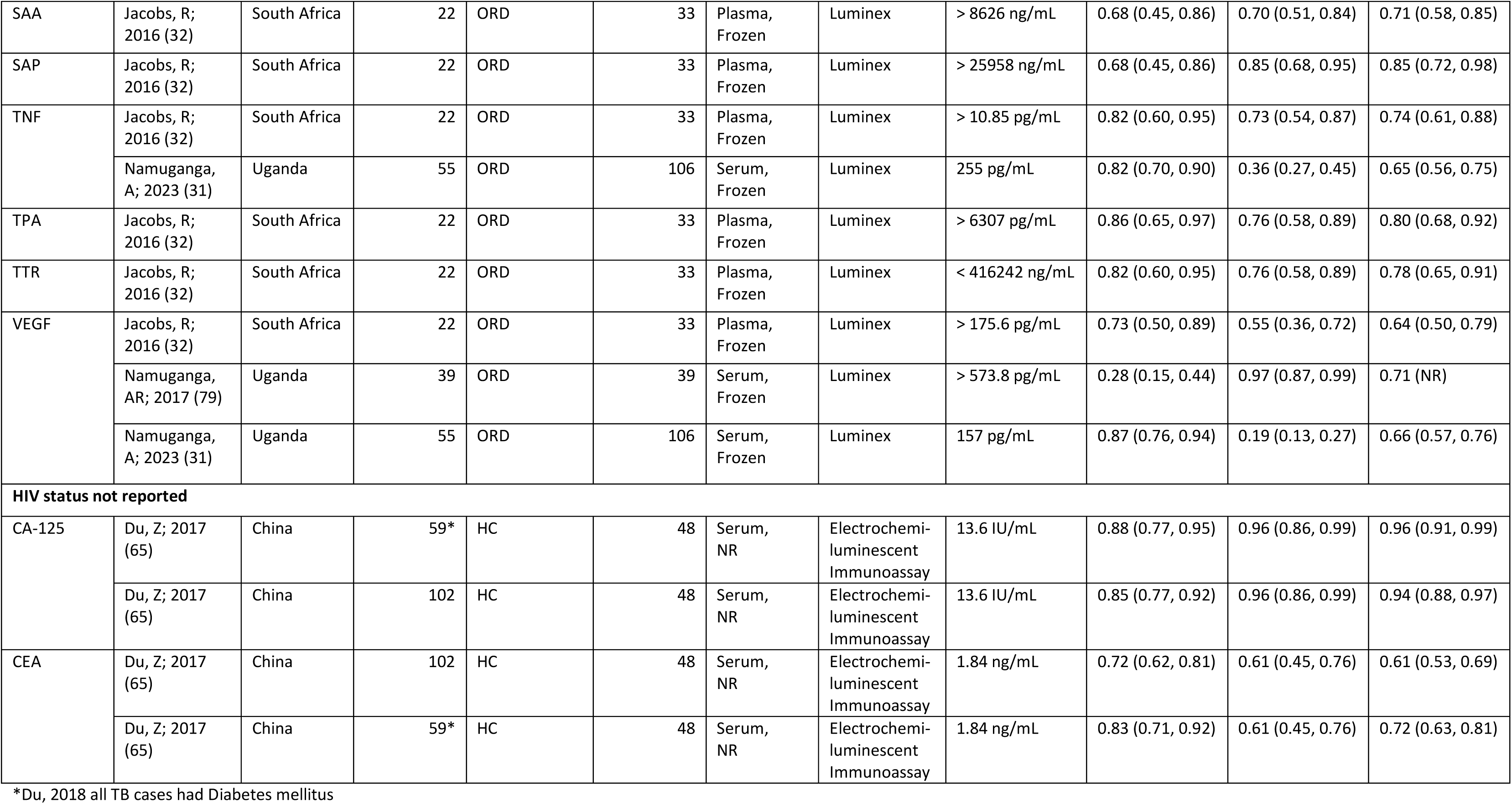
Biomarkers in adults, pulmonary TB, HIV-positive and negative, HIV unknown. Biomarkers meeting TPP criteria are indicated in bold AUC=area under the curve, DM=diabetes mellitus, ELISA=enzyme-linked immunosorbent assay, HC= healthy controls, LTBI=latent TB infection, MSD=Meso Scale Discovery, N=sample size, ORD=other respiratory diseases, NR=not reported

| Biomarker | Study ID | Country | TB cases, N | Control group | Control group, N | Sample type | Testing method | Biomarker cut-off value (unit) | Sensitivity (95% CI) | Specificity (95% CI) | AUC (95% CI) |
| --- | --- | --- | --- | --- | --- | --- | --- | --- | --- | --- | --- |
| **HIV mixed positive and negative** |  |  |  |  |  |  |  |  |  |  |  |
| ADAMTS-13 | Jacobs, R; 2016 (32) | South Africa | 22 | ORD | 33 | Plasma, Frozen | Luminex | > 3466 ng/mL | 0.68 (0.45, 0.86) | 0.64 (0.45, 0.80) | 0.66 (0.51, 0.81) |
| Apo-AI | Jacobs, R; 2016 (32) | South Africa | 22 | ORD | 33 | Plasma, Frozen | Luminex | < 318930 ng/mL | 0.73 (0.50, 0.89) | 0.82 (0.65, 0.93) | 0.76 (0.62, 0.89) |
| AT-III | Jacobs, R; 2016 (32) | South Africa | 22 | ORD | 33 | Plasma, Frozen | Luminex | < 744162 ng/mL | 0.91 (0.71, 0.99) | 0.61 (0.42, 0.77) | 0.70 (0.56, 0.84) |
| BDNF | Jacobs, R; 2016 (32) | South Africa | 22 | ORD | 33 | Plasma, Frozen | Luminex | < 3467 pg/mL | 0.45 (0.24, 0.68) | 0.91 (0.76, 0.98) | 0.69 (0.55, 0.84) |
| bFGF | Namuganga, A; 2023 (31) | Uganda | 55 | ORD | 106 | Serum, Frozen | Luminex | 85 pg/mL | 0.87 (0.76, 0.94) | 0.12 (0.07, 0.20) | 0.63 (0.53, 0.73) |
| CCL14 | Jacobs, R; 2016 (32) | South Africa | 22 | ORD | 33 | Plasma, Frozen | Luminex | > 136956 pg/mL | 0.59 (0.36, 0.79) | 0.85 (0.68, 0.95) | 0.75 (0.61, 0.89) |
| CFH | Jacobs, R; 2016 (32) | South Africa | 22 | ORD | 33 | Plasma, Frozen | Luminex | > 808359 ng/mL | 0.68 (0.45, 0.86) | 0.82 (0.65, 0.93) | 0.72 (0.57, 0.86) |
| CRP | Calderwood, C; 2022 (63) | South Africa | 255 | ORD | 677 | Serum, Frozen | Turbidimetry | 10 mg/L | 0.93 (0.89, 0.95) | 0.54 (0.50, 0.58) | 0.80 (0.77, 0.83) |
|  | Calderwood, C; 2022 (63) | South Africa | 255 | ORD | 677 | Serum, Frozen | Turbidimetry | 5 mg/L | 0.97 (0.94, 0.98) | 0.39 (0.35, 0.42) | 0.80 (0.77, 0.83) |
|  | Jacobs, R; 2016 (32) | South Africa | 22 | ORD | 33 | Plasma, Frozen | Luminex | > 9081 ng/mL (9.1 mg/L) | 0.82 (0.60, 0.95) | 0.9 (0.76, 0.98) | 0.89 (0.79, 1.00) |
|  | Koeppel, L; 2023 (38) | Peru, South Africa, Vietnam | 177 | ORD | 302 | Serum, Frozen | MSD | NR | 0.75 (0.68, 0.81) | 0.70 (0.64, 0.75) | 0.73 (NR) |
|  | Ruperez, M; 2023 (81) | South Africa, Zambia | 76 | ORD | 1231 | Whole blood, Fresh | Immunoassay (POC setting) | ≥5 mg/L | 0.50 (0.38, 0.62) | 0.72 (0.70, 0.75) | 0.61 (0.55, 0.67) |
|  | Ruperez, M; 2023 (81) | South Africa, Zambia | 76 | ORD | 1231 | Whole blood, Fresh | Immunoassay (POC setting) | ≥10 mg/L | 0.36 (0.25, 0.47) | 0.86 (0.84, 0.88) | 0.61 (0.55, 0.67) |
|  | Samuels, T; 2021 (83) | Peru, South Africa, Vietnam, Cambodia, Georgia | 391 | ORD | 374 | Serum, Frozen | ELISA | ≥ 10 mg/L | 0.78 (0.73, 0.82) | 0.66 (0.61, 0.71) | 0.77 (0.74, 0.81) |
|  | Wilson, D; 2011 (30) | South Africa | 135 | ORD | 115 | Serum, NR | Turbidimetry | ≥5 mg/L | 0.98 (0.94, 0.99) | 0.59 (0.50, 0.68) | 0.91 (0.87, 0.95) |
|  | Wilson, D; 2011 (30) | South Africa | 135 | ORD | 115 | Serum, NR | Turbidimetry | ≥ 10 mg/L | **0.95 (0.90, 0.98)** | **0.77 (0.69, 0.85)** | 0.91 (0.87, 0.95) |
| CXCL11 | Jacobs, R; 2016 (32) | South Africa | 22 | ORD | 33 | Plasma, Frozen | Luminex | > 276.5 pg/mL | 0.95 (0.77, 1.0) | 0.36 (0.20, 0.55) | 0.68 (0.54, 0.83) |
| Eotaxin | Namuganga, A; 2023 (31) | Uganda | 55 | ORD | 106 | Serum, Frozen | Luminex | 1887 pg/mL | 0.91 (0.80, 0.96) | 0.11 (0.06, 0.19) | 0.64 (0.54, 0.73) |
| FAS | Jacobs, R; 2016 (32) | South Africa | 22 | ORD | 33 | Plasma, Frozen | Luminex | > 6.7 ng/mL | 0.67 (0.43, 0.85) | 0.68 (0.49, 0.83) | 0.65 (0.50, 0.81) |
| Ferritin | Jacobs, R; 2016 (32) | South Africa | 22 | ORD | 33 | Plasma, Frozen | Luminex | > 93785 pg/mL | 0.91 (0.71, 0.99) | 0.67 (0.48, 0.82) | 0.78 (0.64, 0.92) |
| GDF-15 | Jacobs, R; 2016 (32) | South Africa | 22 | ORD | 33 | Plasma, Frozen | Luminex | > 21.06 ng/mL | 0.91 (0.71, 0.99) | 0.55 (0.36, 0.72) | 0.75 (0.62, 0.88) |
| GM-CSF | Namuganga, A; 2023 (31) | Uganda | 55 | ORD | 106 | Serum, Frozen | Luminex | 147 pg/mL | 0.82 (0.70, 0.90) | 0.40 (0.31, 0.49) | 0.67 (0.58, 0.77) |
| I-309 | Jacobs, R; 2016 (32) | South Africa | 22 | ORD | 33 | Plasma, Frozen | Luminex | > 1.945 pg/mL | 0.68 (0.45, 0.86) | 0.90 (0.73, 0.98) | 0.8 (0.67, 0.93) |
|  | Koeppel, L; 2023 (38) | Peru, South Africa, Vietnam | 177 | ORD | 302 | Serum, Frozen | MSD | NR | 0.81 (0.74, 0.86) | 0.70 (0.64, 0.75) | 0.87 (NR) |
| IFN-γ | Jacobs, R; 2016 (32) | South Africa | 22 | ORD | 33 | Plasma, Frozen | Luminex | > 3.910 pg/mL | 0.91 (0.70, 0.99) | 0.48 (0.31, 0.66) | 0.69 (0.54, 0.83) |
|  | Namuganga, A; 2023 (31) | Uganda | 55 | ORD | 106 | Serum, Frozen | Luminex | 102 pg/mL | 0.91 (0.80, 0.96) | 0.14 (0.09, 0.22) | 0.61 (0.51, 0.70) |
| IL-1β | Namuganga, A; 2023 (31) | Uganda | 55 | ORD | 106 | Serum, Frozen | Luminex | 143 pg/mL | 0.84 (0.72, 0.91) | 0.53 (0.43, 0.62) | 0.74 (0.66, 0.83) |
| IL-2 | Namuganga, A; 2023 (31) | Uganda | 55 | ORD | 106 | Serum, Frozen | Luminex | 95 pg/mL | 0.80 (0.68, 0.88) | 0.48 (0.39, 0.58) | 0.68 (0.59, 0.77) |
| IL-6 | Namuganga, AR; 2017 (79) | Uganda | 39 | ORD | 39 | Serum, Frozen | Luminex | > 36.4 pg/mL | 0.46 (0.30, 0.63) | 0.97 (0.87, 0.99) | 0.85 (NR) |
|  | Namuganga, A; 2023 (31) | Uganda | 55 | ORD | 106 | Serum, Frozen | Luminex | 187 pg/mL | 0.87 (0.76, 0.94) | 0.63 (0.54, 0.72) | 0.83 (0.77, 0.90) |
| IL-8 | Namuganga, A; 2023 (31) | Uganda | 55 | ORD | 106 | Serum, Frozen | Luminex | 131 pg/mL | 0.91 (0.80, 0.96) | 0.12 (0.07, 0.20) | 0.64 (0.55, 0.74) |
| IL-33 | Jacobs, R; 2016 (32) | South Africa | 22 | ORD | 33 | Plasma, Frozen | Luminex | > 131.8 pg/mL | 0.68 (0.45, 0.86) | 0.61 (0.42, 0.77) | 0.63 (0.48, 0.78) |
| IL12p70 | Namuganga, A; 2023 (31) | Uganda | 55 | ORD | 106 | Serum, Frozen | Luminex | 65 pg/mL | 0.82 (0.70, 0.90) | 0.52 (0.42, 0.61) | 0.69 (0.60, 0.78) |
| IP-10 | Jacobs, R; 2016 (32) | South Africa | 22 | ORD | 33 | Plasma, Frozen | Luminex | > 746.6 pg/mL | 0.86 (0.65, 0.97) | 0.73 (0.54, 0.87) | 0.78 (0.64, 0.91) |
|  | Namuganga, A; 2023 (31) | Uganda | 55 | ORD | 106 | Serum, Frozen | Luminex | 4087 pg/mL | **0.93 (0.82, 0.97)** | **0.81 (0.73, 0.87)** | 0.89 (0.84, 0.94) |
| MIG | Jacobs, R; 2016 (32) | South Africa | 22 | ORD | 33 | Plasma, Frozen | Luminex | > 1700 pg/mL | 0.68 (0.45, 0.86) | 0.88 (0.72, 0.97) | 0.81 (0.69, 0.94) |
| MIP-1β | Jacobs, R; 2016 (32) | South Africa | 22 | ORD | 33 | Plasma, Frozen | Luminex | > 212263 ng/mL | 0.32 (0.14, 0.55) | 0.97 (0.84, 1.0) | 0.63 (0.48, 0.79) |
| MIP-4 | Jacobs, R; 2016 (32) | South Africa | 22 | ORD | 33 | Plasma, Frozen | Luminex | > 220.9 pg/mL | 0.50 (0.28, 0.72) | 0.91 (0.76, 0.98) | 0.70 (0.55, 0.85) |
| NCAM-1 | Jacobs, R; 2016 (32) | South Africa | 22 | ORD | 33 | Plasma, Frozen | Luminex | < 477229 pg/mL | **0.91 (0.71, 0.99)** | **0.73 (0.54, 0.87)** | 0.88 (0.78, 0.98) |
| NGAL | Jacobs, R; 2016 (32) | South Africa | 22 | ORD | 33 | Plasma, Frozen | Luminex | > 552.8 ng/mL | 0.59 (0.36, 0.79) | 0.76 (0.58, 0.89) | 0.65 (0.50, 0.80) |
| PCT | Jacobs, R; 2016 (32) | South Africa | 22 | ORD | 33 | Plasma, Frozen | Luminex | > 8101 pg/mL | 0.86 (0.65, 0.97) | 0.67 (0.48, 0.82) | 0.77 (0.64, 0.90) |
| PEDF | Jacobs, R; 2016 (32) | South Africa | 22 | ORD | 33 | Plasma, Frozen | Luminex | > 11423 pg/mL | 0.68 (0.45, 0.86) | 0.64 (0.45, 0.80) | 0.66 (0.50, 0.81) |
| P-selectin | Jacobs, R; 2016 (32) | South Africa | 22 | ORD | 33 | Plasma, Frozen | Luminex | > 265.7 ng/mL | 0.77 (0.55, 0.92) | 0.58 (0.39, 0.75) | 0.67 (0.53, 0.82) |
| RANTES | Namuganga, A; 2023 (31) | Uganda | 55 | ORD | 106 | Serum, Frozen | Luminex | >70,173 pg/mL | 0.82 (0.70, 0.90) | 0.61 (0.52, 0.70) | 0.67 (0.58, 0.77) |
| SAA | Jacobs, R;<br>2016 (32) | South Africa | 22 | ORD | 33 | Plasma,<br>Frozen | Luminex | > 8626 ng/mL | 0.68 (0.45, 0.86) | 0.70 (0.51, 0.84) | 0.71 (0.58, 0.85) |
| SAP | Jacobs, R;<br>2016 (32) | South Africa | 22 | ORD | 33 | Plasma,<br>Frozen | Luminex | > 25958 ng/mL | 0.68 (0.45, 0.86) | 0.85 (0.68, 0.95) | 0.85 (0.72, 0.98) |
| TNF | Jacobs, R;<br>2016 (32) | South Africa | 22 | ORD | 33 | Plasma,<br>Frozen | Luminex | > 10.85 pg/mL | 0.82 (0.60, 0.95) | 0.73 (0.54, 0.87) | 0.74 (0.61, 0.88) |
|  | Namuganga,<br>A; 2023 (31) | Uganda | 55 | ORD | 106 | Serum,<br>Frozen | Luminex | 255 pg/mL | 0.82 (0.70, 0.90) | 0.36 (0.27, 0.45) | 0.65 (0.56, 0.75) |
| TPA | Jacobs, R;<br>2016 (32) | South Africa | 22 | ORD | 33 | Plasma,<br>Frozen | Luminex | > 6307 pg/mL | 0.86 (0.65, 0.97) | 0.76 (0.58, 0.89) | 0.80 (0.68, 0.92) |
| TTR | Jacobs, R;<br>2016 (32) | South Africa | 22 | ORD | 33 | Plasma,<br>Frozen | Luminex | < 416242 ng/mL | 0.82 (0.60, 0.95) | 0.76 (0.58, 0.89) | 0.78 (0.65, 0.91) |
| VEGF | Jacobs, R;<br>2016 (32) | South Africa | 22 | ORD | 33 | Plasma,<br>Frozen | Luminex | > 175.6 pg/mL | 0.73 (0.50, 0.89) | 0.55 (0.36, 0.72) | 0.64 (0.50, 0.79) |
|  | Namuganga,<br>AR; 2017 (79) | Uganda | 39 | ORD | 39 | Serum,<br>Frozen | Luminex | > 573.8 pg/mL | 0.28 (0.15, 0.44) | 0.97 (0.87, 0.99) | 0.71 (NR) |
|  | Namuganga,<br>A; 2023 (31) | Uganda | 55 | ORD | 106 | Serum,<br>Frozen | Luminex | 157 pg/mL | 0.87 (0.76, 0.94) | 0.19 (0.13, 0.27) | 0.66 (0.57, 0.76) |
| **HIV status not reported** |  |  |  |  |  |  |  |  |  |  |  |
| CA-125 | Du, Z; 2017<br>(65) | China | 59* | HC | 48 | Serum,<br>NR | Electrochemi-<br>luminescent<br>Immunoassay | 13.6 IU/mL | 0.88 (0.77, 0.95) | 0.96 (0.86, 0.99) | 0.96 (0.91, 0.99) |
|  | Du, Z; 2017<br>(65) | China | 102 | HC | 48 | Serum,<br>NR | Electrochemi-<br>luminescent<br>Immunoassay | 13.6 IU/mL | 0.85 (0.77, 0.92) | 0.96 (0.86, 0.99) | 0.94 (0.88, 0.97) |
| CEA | Du, Z; 2017<br>(65) | China | 102 | HC | 48 | Serum,<br>NR | Electrochemi-<br>luminescent<br>Immunoassay | 1.84 ng/mL | 0.72 (0.62, 0.81) | 0.61 (0.45, 0.76) | 0.61 (0.53, 0.69) |
|  | Du, Z; 2017<br>(65) | China | 59* | HC | 48 | Serum,<br>NR | Electrochemi-<br>luminescent<br>Immunoassay | 1.84 ng/mL | 0.83 (0.71, 0.92) | 0.61 (0.45, 0.76) | 0.72 (0.63, 0.81) |

\*Du, 2018 all TB cases had Diabetes mellitus

**Table S8.**
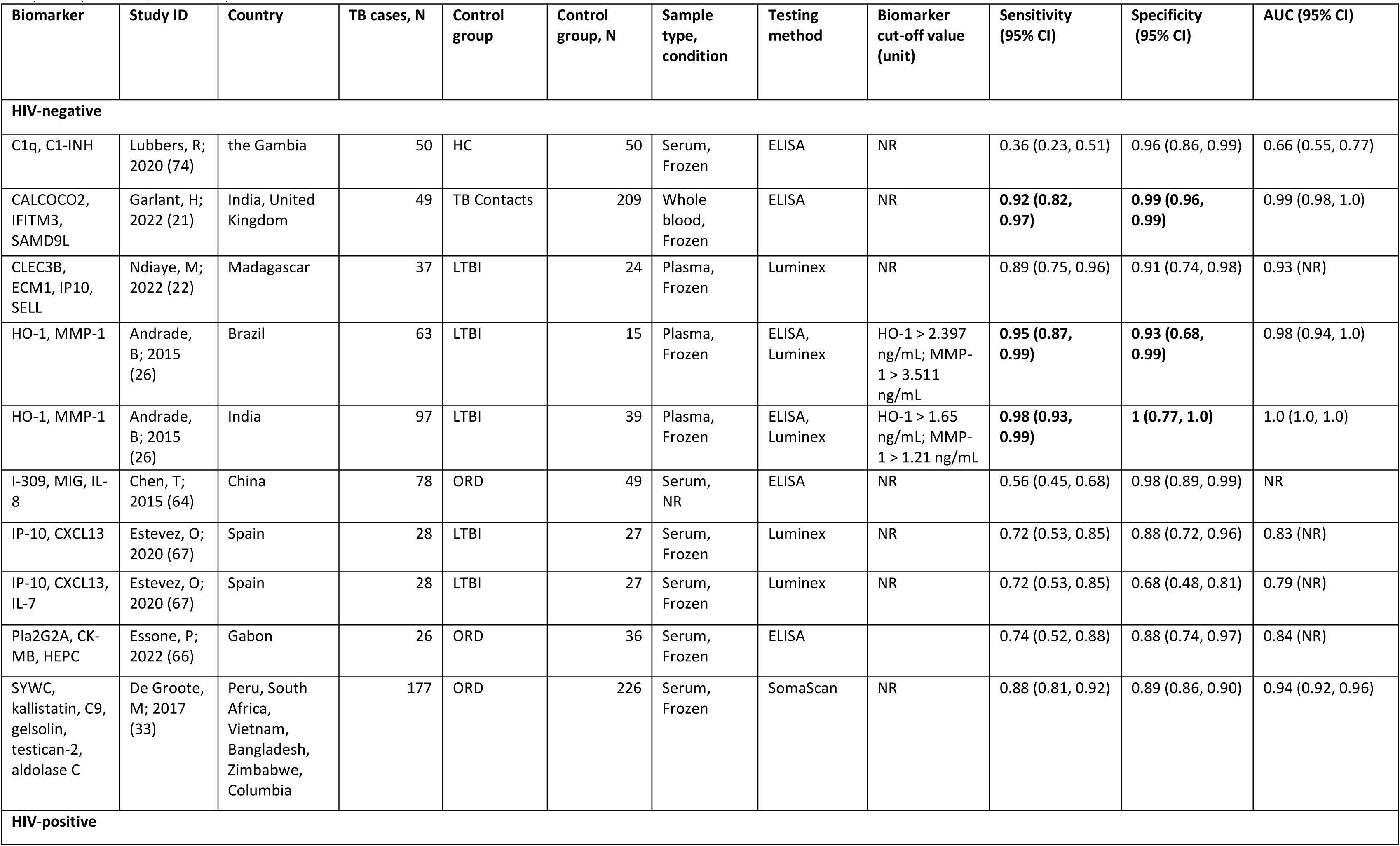
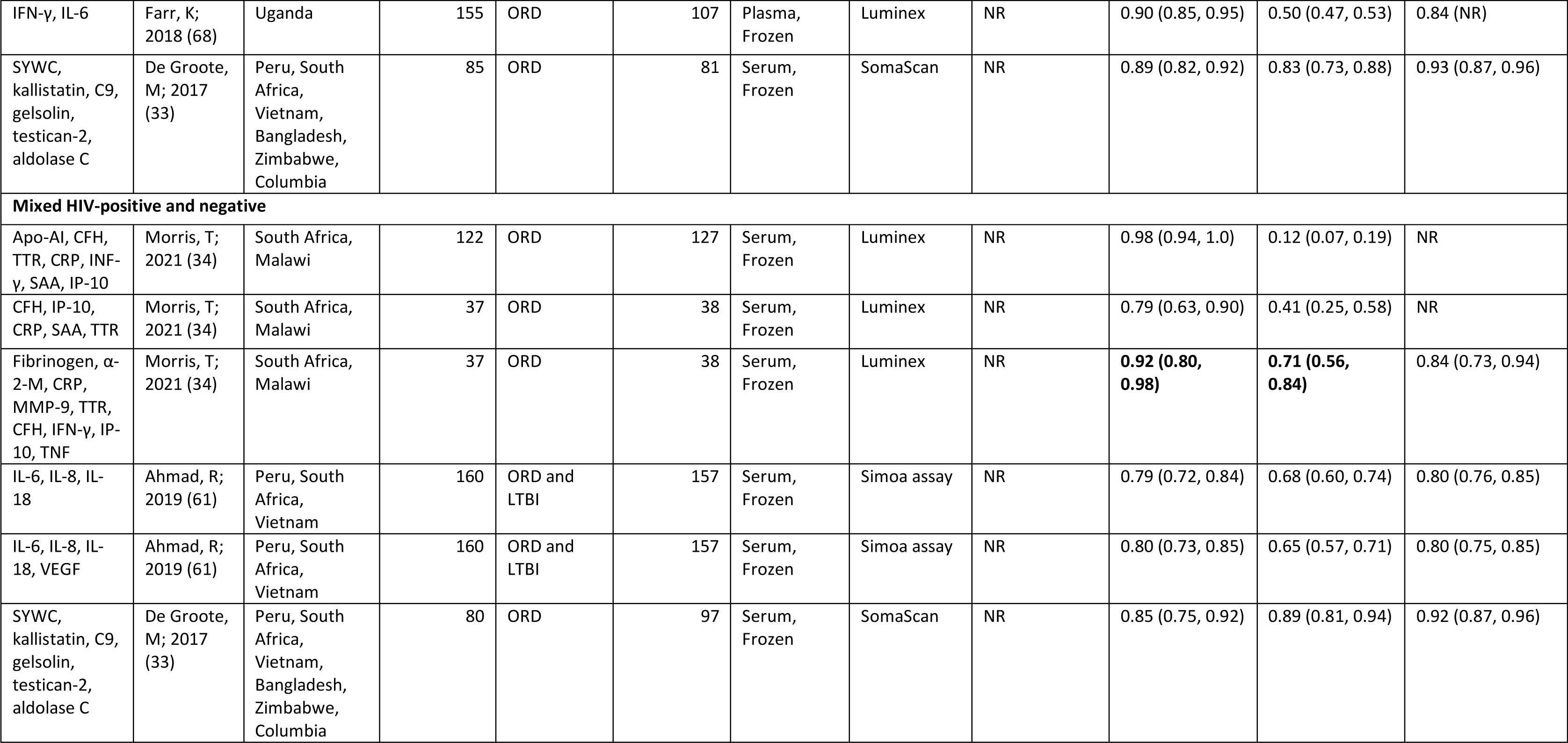
Biomarker signatures for adult pulmonary TB, by HIV status. Biomarkers meeting TPP criteria are indicated in bold AUC=area under the curve, DM=diabetes mellitus, ELISA=enzyme-linked immunosorbent assay, HC= healthy controls, LTBI=latent TB infection, MSD=Meso Scale Discovery, N=sample size, ORD=other respiratory diseases, NR=not reported

| Biomarker | Study ID | Country | TB cases, N | Control group | Control group, N | Sample type, condition | Testing method | Biomarker cut-off value (unit) | Sensitivity (95% CI) | Specificity (95% CI) | AUC (95% CI) |
| --- | --- | --- | --- | --- | --- | --- | --- | --- | --- | --- | --- |
| HIV-negative |  |  |  |  |  |  |  |  |  |  |  |
| C1q, C1-INH | Lubbers, R; 2020 (74) | the Gambia | 50 | HC | 50 | Serum, Frozen | ELISA | NR | 0.36 (0.23, 0.51) | 0.96 (0.86, 0.99) | 0.66 (0.55, 0.77) |
| CALCOCO2, IFITM3, SAMD9L | Garlant, H; 2022 (21) | India, United Kingdom | 49 | TB Contacts | 209 | Whole blood, Frozen | ELISA | NR | **0.92 (0.82, 0.97)** | **0.99 (0.96, 0.99)** | 0.99 (0.98, 1.0) |
| CLEC3B, ECM1, IP10, SELL | Ndiaye, M; 2022 (22) | Madagascar | 37 | LTBI | 24 | Plasma, Frozen | Luminex | NR | 0.89 (0.75, 0.96) | 0.91 (0.74, 0.98) | 0.93 (NR) |
| HO-1, MMP-1 | Andrade, B; 2015 (26) | Brazil | 63 | LTBI | 15 | Plasma, Frozen | ELISA, Luminex | HO-1 > 2.397 ng/mL; MMP-1 > 3.511 ng/mL | **0.95 (0.87, 0.99)** | **0.93 (0.68, 0.99)** | 0.98 (0.94, 1.0) |
| HO-1, MMP-1 | Andrade, B; 2015 (26) | India | 97 | LTBI | 39 | Plasma, Frozen | ELISA, Luminex | HO-1 > 1.65 ng/mL; MMP-1 > 1.21 ng/mL | **0.98 (0.93, 0.99)** | **1 (0.77, 1.0)** | 1.0 (1.0, 1.0) |
| I-309, MIG, IL-8 | Chen, T; 2015 (64) | China | 78 | ORD | 49 | Serum, NR | ELISA | NR | 0.56 (0.45, 0.68) | 0.98 (0.89, 0.99) | NR |
| IP-10, CXCL13 | Estevez, O; 2020 (67) | Spain | 28 | LTBI | 27 | Serum, Frozen | Luminex | NR | 0.72 (0.53, 0.85) | 0.88 (0.72, 0.96) | 0.83 (NR) |
| IP-10, CXCL13, IL-7 | Estevez, O; 2020 (67) | Spain | 28 | LTBI | 27 | Serum, Frozen | Luminex | NR | 0.72 (0.53, 0.85) | 0.68 (0.48, 0.81) | 0.79 (NR) |
| Pla2G2A, CK-MB, HEPC | Essone, P; 2022 (66) | Gabon | 26 | ORD | 36 | Serum, Frozen | ELISA |  | 0.74 (0.52, 0.88) | 0.88 (0.74, 0.97) | 0.84 (NR) |
| SYWC, kallistatin, C9, gelsolin, testican-2, aldolase C | De Groote, M; 2017 (33) | Peru, South Africa, Vietnam, Bangladesh, Zimbabwe, Columbia | 177 | ORD | 226 | Serum, Frozen | SomaScan | NR | 0.88 (0.81, 0.92) | 0.89 (0.86, 0.90) | 0.94 (0.92, 0.96) |
| HIV-positive |  |  |  |  |  |  |  |  |  |  |  |
| IFN-γ, IL-6 | Farr, K; 2018 (68) | Uganda | 155 | ORD | 107 | Plasma, Frozen | Luminex | NR | 0.90 (0.85, 0.95) | 0.50 (0.47, 0.53) | 0.84 (NR) |
| SYWC, kallistatin, C9, gelsolin, testican-2, aldolase C | De Groote, M; 2017 (33) | Peru, South Africa, Vietnam, Bangladesh, Zimbabwe, Columbia | 85 | ORD | 81 | Serum, Frozen | SomaScan | NR | 0.89 (0.82, 0.92) | 0.83 (0.73, 0.88) | 0.93 (0.87, 0.96) |
| **Mixed HIV-positive and negative** |  |  |  |  |  |  |  |  |  |  |  |
| Apo-AI, CFH, TTR, CRP, INF-γ, SAA, IP-10 | Morris, T; 2021 (34) | South Africa, Malawi | 122 | ORD | 127 | Serum, Frozen | Luminex | NR | 0.98 (0.94, 1.0) | 0.12 (0.07, 0.19) | NR |
| CFH, IP-10, CRP, SAA, TTR | Morris, T; 2021 (34) | South Africa, Malawi | 37 | ORD | 38 | Serum, Frozen | Luminex | NR | 0.79 (0.63, 0.90) | 0.41 (0.25, 0.58) | NR |
| Fibrinogen, α-2-M, CRP, MMP-9, TTR, CFH, IFN-γ, IP-10, TNF | Morris, T; 2021 (34) | South Africa, Malawi | 37 | ORD | 38 | Serum, Frozen | Luminex | NR | **0.92 (0.80, 0.98)** | **0.71 (0.56, 0.84)** | 0.84 (0.73, 0.94) |
| IL-6, IL-8, IL-18 | Ahmad, R; 2019 (61) | Peru, South Africa, Vietnam | 160 | ORD and LTBI | 157 | Serum, Frozen | Simoa assay | NR | 0.79 (0.72, 0.84) | 0.68 (0.60, 0.74) | 0.80 (0.76, 0.85) |
| IL-6, IL-8, IL-18, VEGF | Ahmad, R; 2019 (61) | Peru, South Africa, Vietnam | 160 | ORD and LTBI | 157 | Serum, Frozen | Simoa assay | NR | 0.80 (0.73, 0.85) | 0.65 (0.57, 0.71) | 0.80 (0.75, 0.85) |
| SYWC, kallistatin, C9, gelsolin, testican-2, aldolase C | De Groote, M; 2017 (33) | Peru, South Africa, Vietnam, Bangladesh, Zimbabwe, Columbia | 80 | ORD | 97 | Serum, Frozen | SomaScan | NR | 0.85 (0.75, 0.92) | 0.89 (0.81, 0.94) | 0.92 (0.87, 0.96) |

**Table S9:**
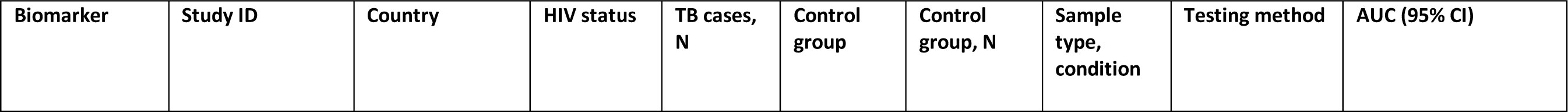
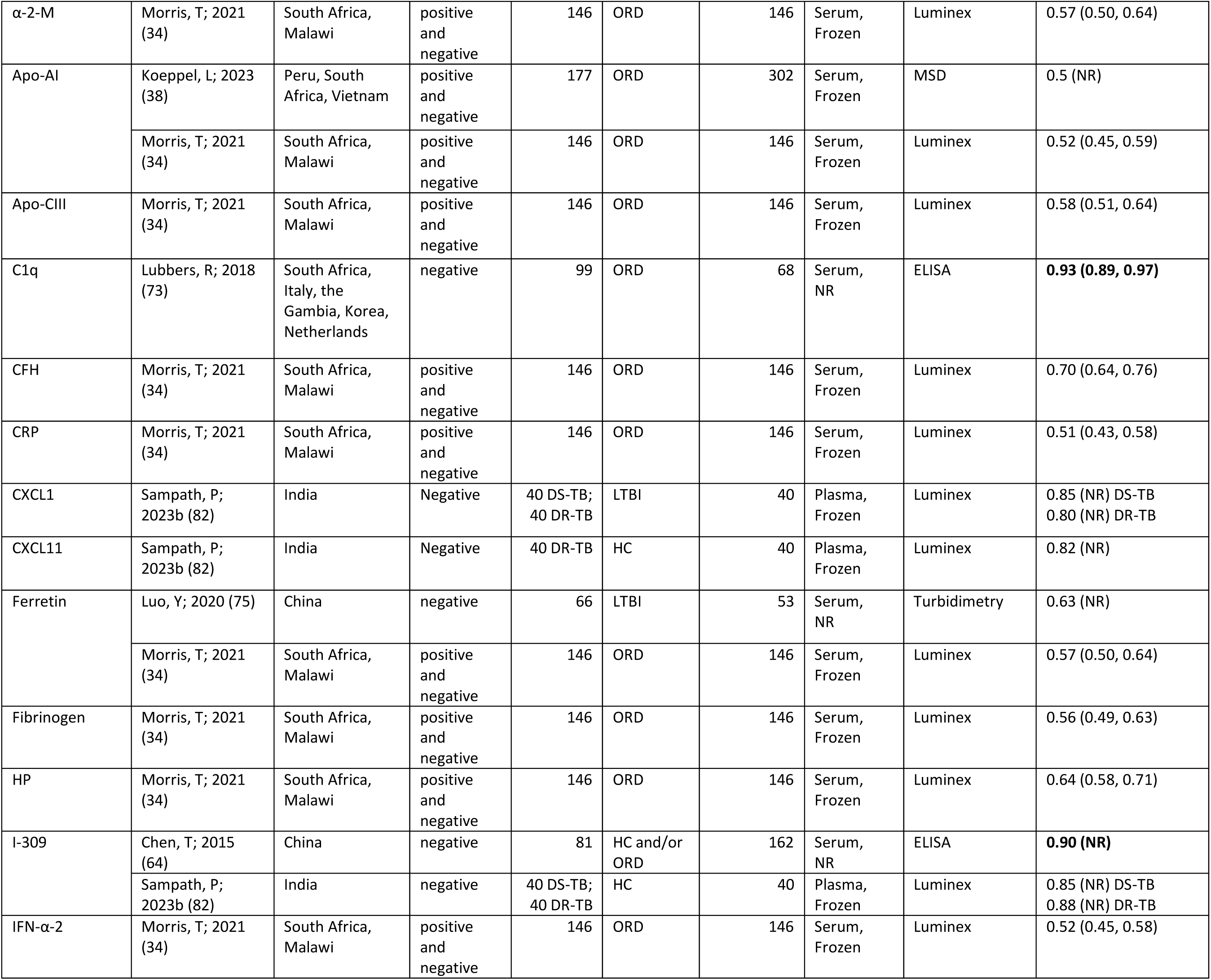
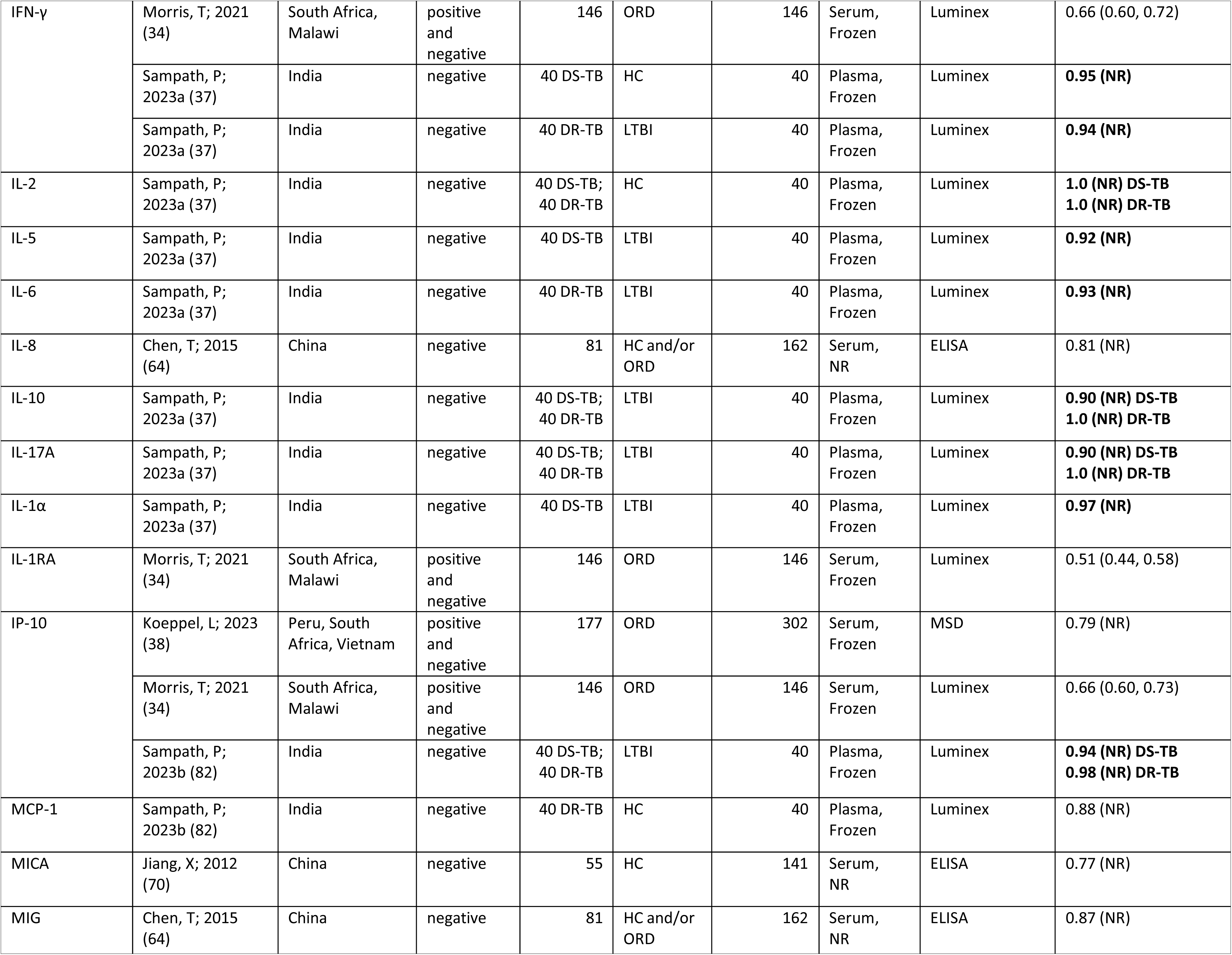
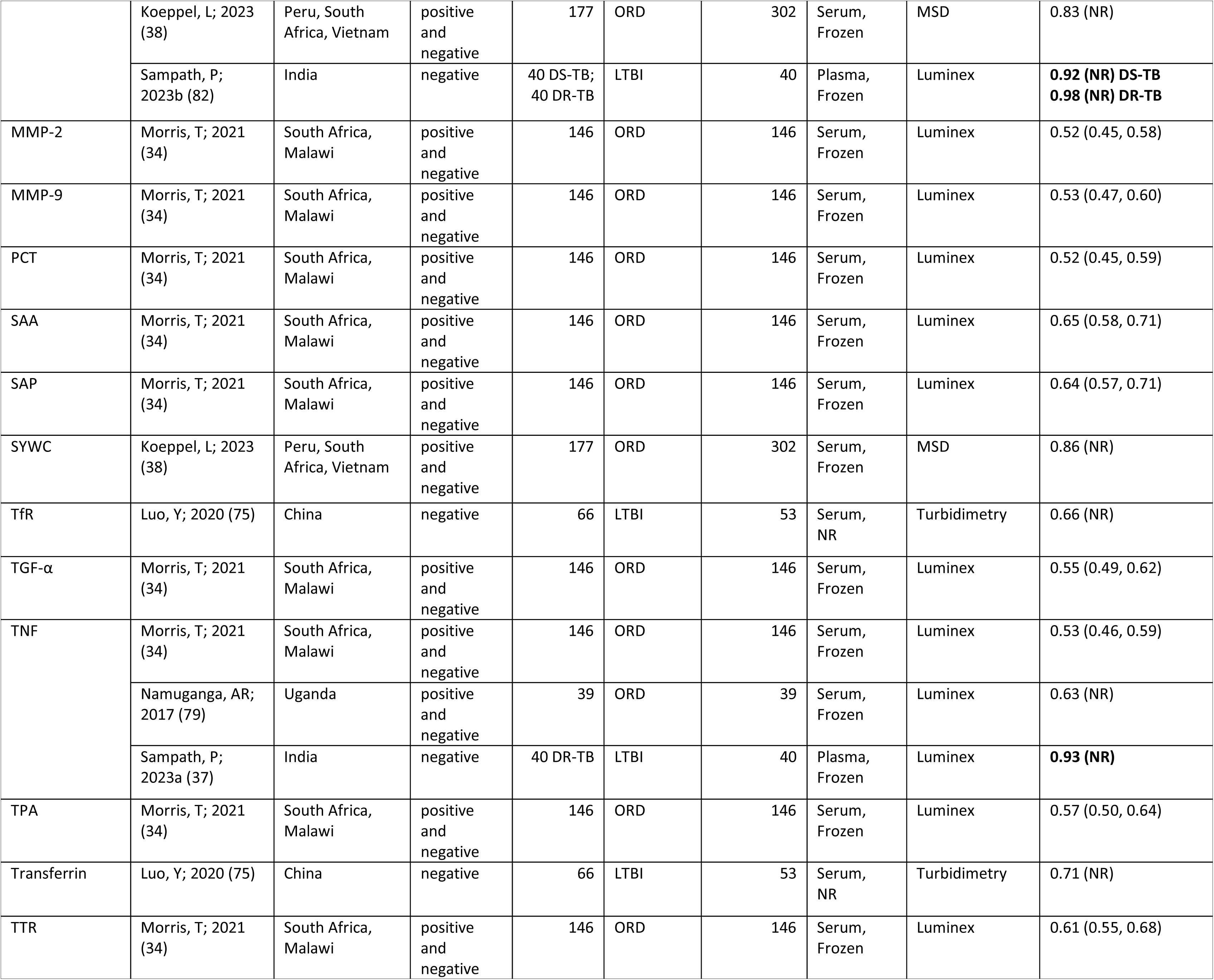
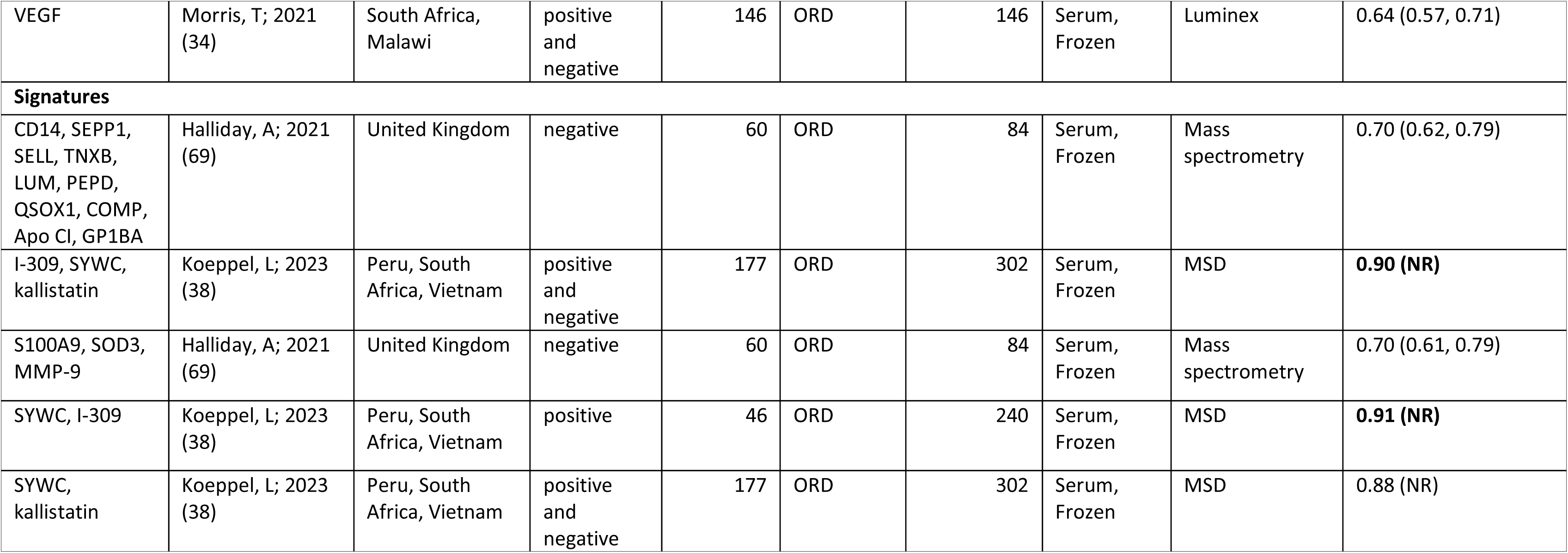
Adult pulmonary TB biomarkers and signatures, reporting only AUC. Biomarkers likely to meet TPP criteria are indicated in bold AUC=area under the curve, DM=diabetes mellitus, ELISA=enzyme-linked immunosorbent assay, HC= healthy controls, LTBI=latent TB infection, MSD=Meso Scale Discovery, N=sample size, ORD=other respiratory diseases, NR=not reported

**Table S10:**
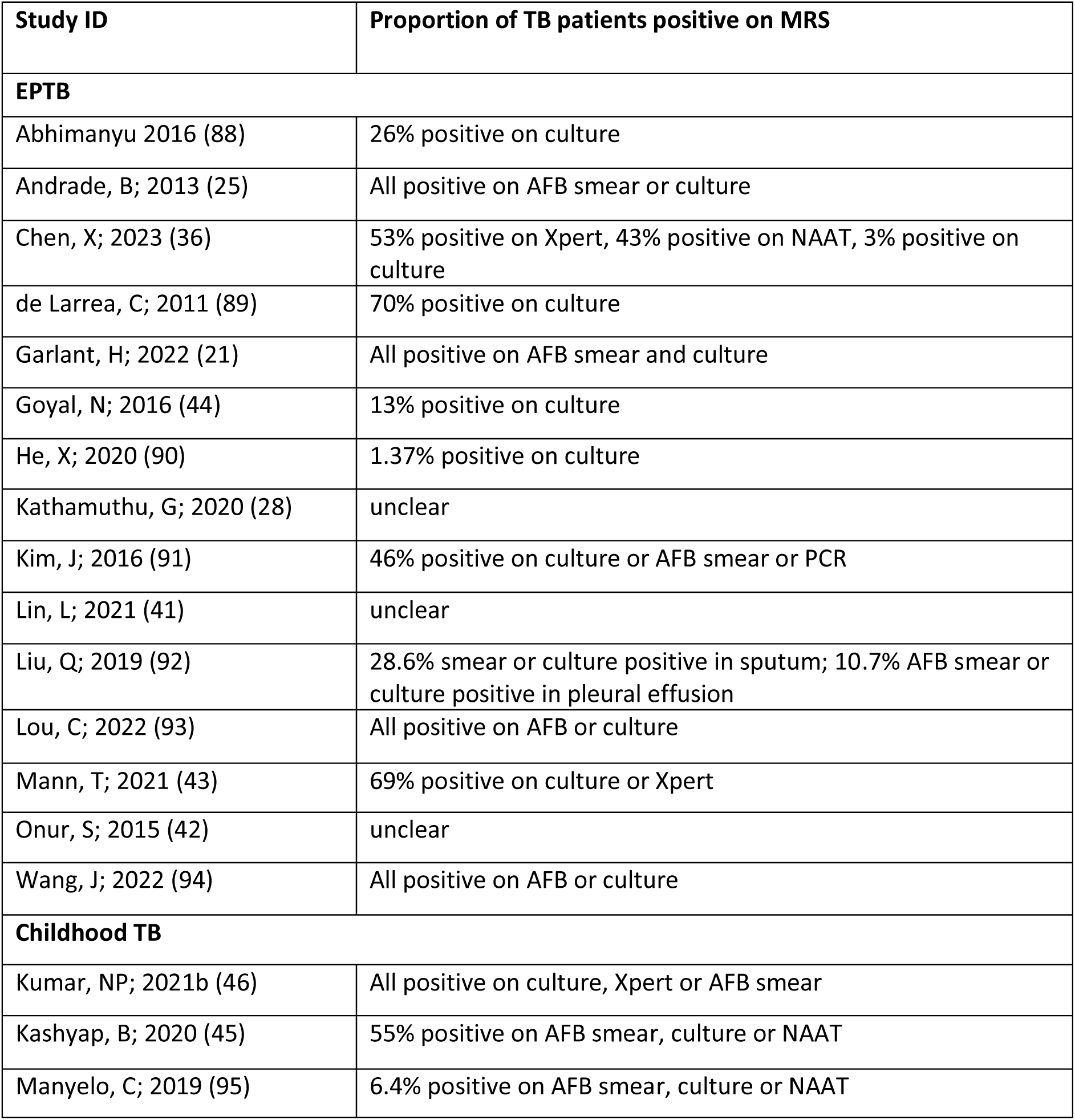
Composite reference standards for adult EPTB and childhood TB.

| Study ID | Proportion of TB patients positive on MRS |
| --- | --- |
| **EPTB** |  |
| Abhimanyu 2016 (88) | 26% positive on culture |
| Andrade, B; 2013 (25) | All positive on AFB smear or culture |
| Chen, X; 2023 (36) | 53% positive on Xpert, 43% positive on NAAT, 3% positive on culture |
| de Larrea, C; 2011 (89) | 70% positive on culture |
| Garlant, H; 2022 (21) | All positive on AFB smear and culture |
| Goyal, N; 2016 (44) | 13% positive on culture |
| He, X; 2020 (90) | 1.37% positive on culture |
| Kathamuthu, G; 2020 (28) | unclear |
| Kim, J; 2016 (91) | 46% positive on culture or AFB smear or PCR |
| Lin, L; 2021 (41) | unclear |
| Liu, Q; 2019 (92) | 28.6% smear or culture positive in sputum; 10.7% AFB smear or culture positive in pleural effusion |
| Lou, C; 2022 (93) | All positive on AFB or culture |
| Mann, T; 2021 (43) | 69% positive on culture or Xpert |
| Onur, S; 2015 (42) | unclear |
| Wang, J; 2022 (94) | All positive on AFB or culture |
| **Childhood TB** |  |
| Kumar, NP; 2021b (46) | All positive on culture, Xpert or AFB smear |
| Kashyap, B; 2020 (45) | 55% positive on AFB smear, culture or NAAT |
| Manyelo, C; 2019 (95) | 6.4% positive on AFB smear, culture or NAAT |

**Table S11:** Adult Extra-pulmonary TB biomarkers and signatures. Biomarkers meeting TPP criteria are indicated in bold AUC=area under the curve, ELISA=enzyme-linked immunosorbent assay, HC= healthy controls, LTBI=latent TB infection, N=sample size, ORD=other respiratory diseases, NR=not reported

| Biomarker | Study ID | Country | Site of disease | HIV-status | TB cases, N | Control group | Control group, N | Sample type | Testing method | Biomarker cut-off value (unit) | Sensitivity (95% CI) | Specificity (95% CI) | AUC (95% CI) |
| --- | --- | --- | --- | --- | --- | --- | --- | --- | --- | --- | --- | --- | --- |
| ADA | He, X; 2020 (90) | China | Abdominal | negative | 73 | OD + HC | 135 | Serum, NR | Peroxidase method | 15.31 IU/mL | 0.74 (0.62, 0.84) | 0.75 (0.67, 0.82) | 0.73 (0.65, 0.80) |
| Apo-AI | Mann, T; 2021 (43) | South Africa | Bone | positive and negative | 26 | OD | 17 | Serum, Frozen | Luminex | NR | 0.71 (0.44, 0.90) | 0.54 (0.33, 0.73) | 0.61 (0.43, 0.80) |
| Apo-CII | Chen, X; 2023 (36) | China | Bone | negative | 30 | OD | 30 | Serum, Frozen | ELISA | 7.788 µg/mL | **0.97 (0.83, 0.99)** | **0.93 (0.78, 0.99)** | NR |
| Apo-E | Chen, X; 2023 (36) | China | Bone | negative | 30 | OD | 30 | Serum, Frozen | ELISA | 49.46 µg/mL | 0.60 (0.42, 0.75) | 0.57 (0.39, 0.73) | NR |
| CA-125 | He, X; 2020 (90) | China | Abdominal | negative | 73 | OD + HC | 135 | Serum, NR | Electrochemiluminescent Immunoassay | 30.34 IU/mL | 0.85 (0.75, 0.92) | 0.60 (0.51, 0.68) | 0.72 (0.65, 0.79) |
| CALCOCO2 | Garlant, H; 2022 (21) | India, United Kingdom | NR | negative | 82 | LTBI | 111 | Whole blood, Frozen | ELISA | 990.7 ng/mL | 0.90 (0.83, 0.95) | 0.40 (0.32, 0.49) | 0.72 (0.66, 0.78) |
| CD163 | Liu, Q; 2019 (92) | China | Pleural | negative | 28 | OD | 21 | Plasma, Frozen | ELISA | 934.7 ng/mL | 0.77 (0.59, 0.92) | 0.80 (0.58, 0.95) | 0.81 (NR) |
| CD52 | Garlant, H; 2022 (21) | India, United Kingdom | NR | negative | 82 | LTBI | 111 | Whole blood, Frozen | ELISA | 964.8 ng/mL | 0.90 (0.83, 0.95) | 0.14 (0.09, 0.21) | 0.59 (0.52, 0.66) |
| CFD | Mann, T; 2021 (43) | South Africa | Bone | positive and negative | 26 | OD | 17 | Serum, Frozen | Luminex | NR | 0.59 (0.33, 0.82) | 0.65 (0.44, 0.83) | 0.62 (0.44, 0.79) |
| CFH | Mann, T; 2021 (43) | South Africa | Bone | positive and negative | 26 | OD | 17 | Serum, Frozen | Luminex | NR | 0.53 (0.23, 0.72) | 0.62 (0.20, 0.59) | 0.51 (0.33, 0.70) |
| CFHR2 | Chen, X; 2023 (36) | China | Bone | negative | 30 | OD | 30 | Serum, Frozen | ELISA | 86.90 µg/ml | 0.80 (0.63, 0.90) | 0.73 (0.56, 0.86) | 0.86 (0.77, 0.95) |
| CFHR3 | Chen, X; 2023 (36) | China | Bone | negative | 30 | OD | 30 | Serum, Frozen | ELISA | 138.25 µg/ml | 0.77 (0.59, 0.88) | 0.77 (0.59, 0.88) | 0.82 (0.70, 0.93) |
| CFHR5 | Chen, X; 2023 (36) | China | Bone | negative | 30 | OD | 30 | Serum, Frozen | ELISA | 2.67 µg/ml | **0.97 (0.83, 0.99)** | **0.97 (0.83, 0.99)** | 0.99 (0.99, 1.0) |
| CRP | Andrade, B; 2013 (25) | India | Pleural, Lymph node | negative | 35 | PTB | 97 | Plasma, NR | Luminex | NR | 0.75 (0.65, 0.83) | 0.39 (0.23, 0.58) | 0.62 (0.52, 0.72) |
|  | He, X; 2020 (90) | China | Abdominal | negative | 73 | OD + HC | 135 | Serum, NR | Turbidimetry | 43.54 IU/mL | 0.68 (0.57, 0.79) | 0.85 (0.78, 0.91) | 0.80 (0.74, 0.87) |
|  | Lin, L; 2021 (41) | China | Pleural | negative | 28 | OD | 16 | Serum, NR | Turbidimetry | ≤ 77.1 mg/L | 0.75 (0.55, 0.89) | 0.88 (0.62, 0.98) | 0.84 (0.70, 0.93) |
|  | Lin, L; 2021 (41) | China | Pleural | negative | 28 | OD | 7 | Serum, NR | Turbidimetry | > 19.3 mg/L | 0.75 (0.55, 0.89) | 1 (0.59, 1.0) | 0.91 (0.76, 0.98) |
|  | Mann, T; 2021 (43) | South Africa | Bone | positive and negative | 26 | OD | 17 | Serum, Frozen | Luminex | 79091 ng/mL | 0.89 (0.70, 0.98) | 0.94 (0.71, 0.99) | 0.95 (0.87, 1.0) |
|  | Kim, J; 2016 (91) | South Korea | Meningeal | negative | 26 | OD | 70 | Serum, NR | NR | NR | NR | NR | 0.78 (0.69, 0.86) |
|  | Wang, J; 2022 (94) | China | Bone | NR | 570 | OD | 147 | Serum, NR | NR | NR | NR | NR | 0.73 (0.67, 0.79) |
| Ferritin | Mann, T; 2021 (43) | South Africa | Bone | positive and negative | 26 | OD | 17 | Serum, Frozen | Luminex | NR | 0.69 (0.48, 0.86) | 0.82 (0.82, 0.96) | 0.78 (0.64, 0.92) |
| Fibrinogen | Mann, T; 2021 (43) | South Africa | Bone | positive and negative | 26 | OD | 17 | Serum, Frozen | Luminex | 3696 ng/mL | **1 (0.89, 1.0)** | **0.94 (0.71, 0.99)** | 0.99 (0.96, 1.0) |
| GBP1 | Garlant, H; 2022 (21) | India, United Kingdom | NR | negative | 82 | LTBI | 111 | Whole blood, Frozen | ELISA | 1394 ng/mL | 0.90 (0.83, 0.95) | 0.36 (0.28, 0.45) | 0.78 (0.72, 0.84) |
| GDF-15 | Mann, T; 2021 (43) | South Africa | Bone | positive and negative | 26 | OD | 17 | Serum, Frozen | Luminex | NR | 0.62 (0.41, 0.80) | 0.71 (0.44, 0.90) | 0.64 (0.48, 0.81) |
| HO-1 | Andrade, B; 2013 (25) | India | Pleural, Lymph nodes | negative | 35 | PTB | 97 | Plasma, NR | ELISA | NR | 0.74 (0.64, 0.83) | 0.53 (0.34, 0.72) | 0.73 (0.65, 0.82) |
| I-309 | Mann, T; 2021 (43) | South Africa | Bone | positive and negative | 26 | OD | 17 | Serum, Frozen | Luminex | NR | 0.73 (0.52, 0.88) | 0.65 (0.38, 0.86) | 0.74 (0.59, 0.90) |
| ICAM-1 | Mann, T; 2021 (43) | South Africa | Bone | positive and negative | 26 | OD | 17 | Serum, Frozen | Luminex | NR | 0.58 (0.37, 0.77) | 0.71 (0.44, 0.90) | 0.60 (0.41, 0.78) |
| IFIT3 | Garlant, H; 2022 (21) | India, United Kingdom | NR | negative | 82 | LTBI | 111 | Whole blood, Frozen | ELISA | 14.43 ng/mL | **0.90 (0.83, 0.95)** | **0.89 (0.83, 0.94)** | 0.94 (0.90, 0.97) |
| IFITM3 | Garlant, H; 2022 (21) | India, United Kingdom | NR | negative | 82 | LTBI | 111 | Whole blood, Frozen | ELISA | 1303 ng/mL | **0.90 (0.83, 0.95)** | **0.71 (0.62, 0.78)** | 0.91 (0.88, 0.95) |
| IFN-γ | Abhimanyu, BM; 2016 (88) | India | Lymph node | negative | 50 | PTB | 84 | Serum, Frozen | ELISA | NR | NR | NR | 0.49 (0.39, 0.59) |
|  | de Larrea, C; 2011 (89) | Venezuela | Pleural | negative | 20 | OD | 40 | Serum, NR | ELISA | 0.162 pg/mL | 0.65 (0.44, 0.90) | 0.55 (0.40, 0.70) | NR |
|  | Goyal, N; 2016 (44) | India | Pleural, lymph node, CSF, urine, ascitic fluid | negative | 69 | HC | 69 | Serum, Frozen | ELISA | 11.4 pg/mL | **0.99 (0.92, 0.99)** | **0.97 (0.90, 0.99)** | 0.99 (0.97, 1.0) |
|  | Mann, T; 2021 (43) | South Africa | Bone | positive and negative | 26 | OD | 17 | Serum, Frozen | Luminex | 8.56 pg/mL | **0.92 (0.75, 0.99)** | **0.94 (0.71, 0.99)** | 0.92 (0.83, 1.0) |
| IL-1β | Abhimanyu, BM; 2016 (88) | India | Lymph node | negative | 50 | PTB | 84 | Serum, Frozen | ELISA | NR | NR | NR | 0.55 (0.44, 0.65) |
| IL-1RA | Abhimanyu, BM; 2016 (88) | India | Lymph node | negative | 50 | PTB | 84 | Serum, Frozen | ELISA | NR | NR | NR | 0.43 (0.34, 0.52) |
| IL-2 | Abhimanyu, BM; 2016 (88) | India | Lymph node | negative | 50 | PTB | 84 | Serum, Frozen | ELISA | NR | NR | NR | 0.58 (0.48, 0.68) |
|  | Goyal, N; 2016 (44) | India | Pleural, lymph node, CSF, urine, ascitic fluid | negative | 69 | HC | 89 | Serum, Frozen | ELISA | NR | NR | NR | 0.50 (0.42, 0.59) |
| IL-2R | Onur, S; 2015 (42) | Turkey | Pleural | negative | 52 | OD | 68 | Serum, Frozen | ELISA | > 0.6 ng/mL | 0.83 (0.69, 0.92) | 0.53 (0.40, 0.65) | NR |
| IL-4 | Abhimanyu, BM; 2016 (88) | India | Lymph node | negative | 50 | PTB | 84 | Serum, Frozen | ELISA | NR | NR | NR | 0.38 (0.28, 0.47) |
| IL-6 | Abhimanyu, BM; 2016 (88) | India | Lymph node | negative | 50 | PTB | 84 | Serum, Frozen | ELISA | NR | NR | NR | 0.58 (0.49, 0.67) |
| IL-8 | Abhimanyu, BM; 2016 (88) | India | Lymph node | negative | 50 | PTB | 84 | Serum, Frozen | ELISA | NR | NR | NR | 0.79 (0.72, 0.86) |
|  | Mann, T;<br>2021 (43) | South<br>Africa | Bone | positive<br>and<br>negative | 26 | OD | 17 | Serum,<br>Frozen | Luminex | NR | 0.65 (0.38, 0.86) | 0.62 (0.41, 0.80) | 0.59 (0.41, 0.78) |
| IL-10 | Abhimanyu,<br>BM; 2016<br>(88) | India | Lymph node | negative | 50 | PTB | 84 | Serum,<br>Frozen | ELISA | NR | NR | NR | 0.74 (0.64, 0.85) |
|  | Mann, T;<br>2021 (43) | South<br>Africa | Bone | positive<br>and<br>negative | 26 | OD | 17 | Serum,<br>Frozen | Luminex | NR | 0.58 (0.37, 0.77) | 0.77 (0.50, 0.93) | 0.68 (0.53, 0.82) |
| IL-12 | Abhimanyu,<br>BM; 2016<br>(88) | India | Lymph node | negative | 50 | PTB | 84 | Serum,<br>Frozen | ELISA | NR | NR | NR | 0.76 (0.69, 0.83) |
| IL-12p40 | de Larrea, C;<br>2011 (89) | Venezuela | Pleural | negative | 20 | OD | 40 | Serum,<br>NR | ELISA | 0.497<br>pg/mL | 0.95 (0.86, 1.05) | 0.50 (0.35, 0.66) | NR |
|  | Onur, S;<br>2015 (42) | Turkey | Pleural | negative | 52 | OD | 68 | Serum,<br>Frozen | ELISA | > 42 pg/mL | 0.81 (0.68, 0.90) | 0.40 (0.28, 0.52) | NR |
| IL-18 | Abhimanyu,<br>BM; 2016<br>(88) | India | Lymph node | negative | 50 | PTB | 84 | Serum,<br>Frozen | ELISA | NR | NR | NR | 0.58 (0.47, 0.69) |
| IP-10 | Mann, T;<br>2021 (43) | South<br>Africa | Bone | positive<br>and<br>negative | 26 | OD | 17 | Serum,<br>Frozen | Luminex | NR | 0.62 (0.41, 0.79) | 0.71 (0.44, 0.90) | 0.63 (0.46, 0.80) |
| ITIH2 | Chen, X;<br>2023 (36) | China | Bone | negative | 30 | OD | 30 | Serum,<br>Frozen | ELISA | 99.85<br>ng/mL | 0.73 (0.56, 0.86) | 0.67 (0.49, 0.81) | 0.72 (0.58, 0.85) |
| KNG1 | Chen, X;<br>2023 (36) | China | Bone | negative | 30 | OD | 30 | Serum,<br>Frozen | ELISA | 157.32<br>µg/ml | 0.63 (0.46, 0.78) | 0.60 (0.42, 0.75) | NR |
| LBP | Lou, C; 2022<br>(93) | China | Bone | negative | 50 | HC | 30 | Serum,<br>Frozen | ELISA | NR | NR | NR | 0.88 (NR) |
| LTA | Abhimanyu,<br>BM; 2016<br>(88) | India | Lymph node | negative | 50 | PTB | 84 | Serum,<br>Frozen | ELISA | NR | NR | NR | 0.61 (0.52, 0.69) |
| MIG | Mann, T;<br>2021 (43) | South<br>Africa | Bone | positive<br>and<br>negative | 26 | OD | 17 | Serum,<br>Frozen | Luminex | NR | 0.58 (0.37, 0.77) | 0.88 (0.64, 0.99) | 0.62 (0.44, 0.79) |
| MMP-1 | Kathamuthu,<br>G; 2020 (28) | India | Cervical<br>lymphadenopathy | negative | 44 | LTBI | 44 | Plasma,<br>NR | Luminex | NR | 0.59 (0.43, 0.74) | 0.70 (0.55, 0.83) | 0.68 (NR) |
| MMP-12 | Kathamuthu, G; 2020 (28) | India | Cervical lymphadenopathy | negative | 44 | LTBI | 44 | Plasma, NR | Luminex | NR | 0.52 (0.37, 0.68) | 0.55 (0.39, 0.69) | 0.50 (NR) |
| MMP-13 | Kathamuthu, G; 2020 (28) | India | Cervical lymphadenopathy | negative | 44 | LTBI | 44 | Plasma, NR | Luminex | NR | **1.0 (0.920, 1.0)** | **1.0 (0.920, 1.0)** | 1.0 (NR) |
| MMP-2 | Kathamuthu, G; 2020 (28) | India | Cervical lymphadenopathy | negative | 44 | LTBI | 44 | Plasma, NR | Luminex | NR | 0.61 (0.46, 0.76) | 0.65 (0.50, 0.79) | 0.62 (NR) |
| MMP-3 | Kathamuthu, G; 2020 (28) | India | Cervical lymphadenopathy | negative | 44 | LTBI | 44 | Plasma, NR | Luminex | NR | 0.57 (0.41, 0.72) | 0.55 (0.39, 0.69) | 0.56 (NR) |
| MMP-7 | Kathamuthu, G; 2020 (28) | India | Cervical lymphadenopathy | negative | 44 | LTBI | 44 | Plasma, NR | Luminex | NR | 0.75 (0.60, 0.87) | 0.70 (0.55, 0.83) | 0.80 (NR) |
| MMP-8 | Kathamuthu, G; 2020 (28) | India | Cervical lymphadenopathy | negative | 44 | LTBI | 44 | Plasma, NR | Luminex | NR | 0.66 (0.50, 0.79) | 0.55 (0.39, 0.69) | 0.66 (NR) |
| MMP-9 | Kathamuthu, G; 2020 (28) | India | Cervical lymphadenopathy | negative | 44 | LTBI | 44 | Plasma, NR | Luminex | NR | 0.50 (0.35, 0.65) | 0.55 (0.39, 0.69) | 0.59 (NR) |
| MPO | Mann, T; 2021 (43) | South Africa | Bone | positive and negative | 26 | OD | 17 | Serum, Frozen | Luminex | NR | 0.58 (0.37, 0.77) | 0.76 (0.44, 0.90) | 0.60 (0.43, 0.80) |
| NCAM-1 | Mann, T; 2021 (43) | South Africa | Bone | positive and negative | 26 | OD | 17 | Serum, Frozen | Luminex | 299421 pg/mL | 0.77 (0.56, 0.91) | 0.88 (0.64, 0.98) | 0.88 (0.78, 0.98) |
| OPG | Mann, T; 2021 (43) | South Africa | Bone | positive and negative | 26 | OD | 17 | Serum, Frozen | Luminex | NR | 0.54 (0.27, 0.67) | 0.65 (0.14, 0.62) | 0.56 (0.38, 0.74) |
| PCT | Kim, J; 2016 (91) | South Korea | Meningeal | negative | 26 | OD | 70 | Serum, NR | Electrochemiluminescent Immunoassay | ≤ 1.27 ng/mL | 0.96 (0.80, 0.99) | 0.63 (0.51, 0.74) | 0.84 (0.75, 0.90) |
| PD-L1 | Garlant, H; 2022 (21) | India, United Kingdom | NR | negative | 49 | LTBI | 111 | Whole blood, Frozen | ELISA | 0.5756 ng/mL | **1.0 (0.96, 1.0)** | **1.0 (0.97, 1.0)** | 1.0 (1.0, 1.0) |
| SAA | Andrade, B; 2013 (25) | India | Pleural, Lymph node | negative | 35 | PTB | 97 | Plasma, NR | Luminex | NR | 0.79 (0.69, 0.87) | 0.39 (0.23, 0.58) | 0.66 (0.56, 0.77) |
| SAA-4 | Chen, X;<br>2023 (36) | China | Bone | negative | 30 | OD | 30 | Serum,<br>Frozen | ELISA | 2.197<br>µg/ml | 0.73 (0.56, 0.86) | 0.70 (0.52, 0.83) | NR |
| SAMD9L | Garlant, H;<br>2022 (21) | India,<br>United<br>Kingdom | NR | negative | 82 | LTBI | 111 | Whole<br>blood,<br>Frozen | ELISA | 1439 ng/mL | **0.90 (0.83,**<br>**0.95)** | **0.88 (0.82,**<br>**0.93)** | 0.96 (0.94, 0.98) |
| SNX10 | Garlant, H;<br>2022 (21) | India,<br>United<br>Kingdom | NR | negative | 82 | LTBI | 111 | Whole<br>blood,<br>Frozen | ELISA | 1739 ng/mL | 0.90 (0.83, 0.95) | 0.37 (0.30, 0.46) | 0.81 (0.76, 0.87) |
| TIMP-1 | Kathamuthu,<br>G; 2020 (28) | India | Cervical<br>lymphadenopathy | negative | 44 | LTBI | 44 | Plasma,<br>NR | Luminex | NR | 0.57 (0.41, 0.72) | 0.64 (0.48, 0.78) | 0.61 (NR) |
| TIMP-2 | Kathamuthu,<br>G; 2020 (28) | India | Cervical<br>lymphadenopathy | negative | 44 | LTBI | 44 | Plasma,<br>NR | Luminex | NR | 0.82 (0.67, 0.92) | 0.91 (0.78, 0.97) | 0.86 (NR) |
| TIMP-3 | Kathamuthu,<br>G; 2020 (28) | India | Cervical<br>lymphadenopathy | negative | 44 | LTBI | 44 | Plasma,<br>NR | Luminex | NR | 0.65 (0.48, 0.77) | 0.82 (0.67, 0.92) | 0.62 (NR) |
| TIMP-4 | Kathamuthu,<br>G; 2020 (28) | India | Cervical<br>lymphadenopathy | negative | 44 | LTBI | 44 | Plasma,<br>NR | Luminex | NR | 0.62 (0.46, 0.76) | 0.73 (0.57, 0.85) | 0.69 (NR) |
| TMEM49 | Garlant, H;<br>2022 (21) | India,<br>United<br>Kingdom | NR | negative | 82 | LTBI | 111 | Whole<br>blood,<br>Frozen | ELISA | 266.3<br>ng/mL | 0.90 (0.83, 0.95) | 0.0 (0.0, 0.3) | 0.42 (0.35, 0.50) |
| TNF | Abhimanyu,<br>BM; 2016<br>(88) | India | Lymph node | negative | 50 | PTB | 84 | Serum,<br>Frozen | ELISA | NR | NR | NR | 0.38 (0.28, 0.48) |
| VCAM-1 | Mann, T;<br>2021 (43) | South<br>Africa | Bone | positive<br>and<br>negative | 26 | OD | 17 | Serum,<br>Frozen | Luminex | NR | 0.54 (0.33, 0.73) | 0.71 (0.44, 0.90) | 0.61 (0.44, 0.79) |
| VEGF | Mann, T;<br>2021 (43) | South<br>Africa | Bone | positive<br>and<br>negative | 26 | OD | 17 | Serum,<br>Frozen | Luminex | NR | 0.50 (0.30, 0.70) | 0.88 (0.64, 0.99) | 0.63 (0.46, 0.80) |
| **Signatures** |  |  |  |  |  |  |  |  |  |  |  |  |  |
| CFHR2,<br>CFHR3 | Chen, X;<br>2023 (36) | China | Bone | negative | 30 | OD | 30 | Serum,<br>Frozen | ELISA | NR | **0.90 (0.74,**<br>**0.97)** | **0.88 (0.70,**<br>**0.95)** | 0.94 (0.87, 0.98) |
| CRP, NCAM,<br>Ferritin, IL-8,<br>GDF-15 | Mann, T;<br>2021 (43) | South<br>Africa | Bone | positive<br>and<br>negative | 26 | OD | 17 | Serum,<br>Frozen | Luminex | NR | **1 (0.89, 1.0)** | **1 (0.84, 1.0)** | 1 (1.0, 1.0) |

**Table S12:**
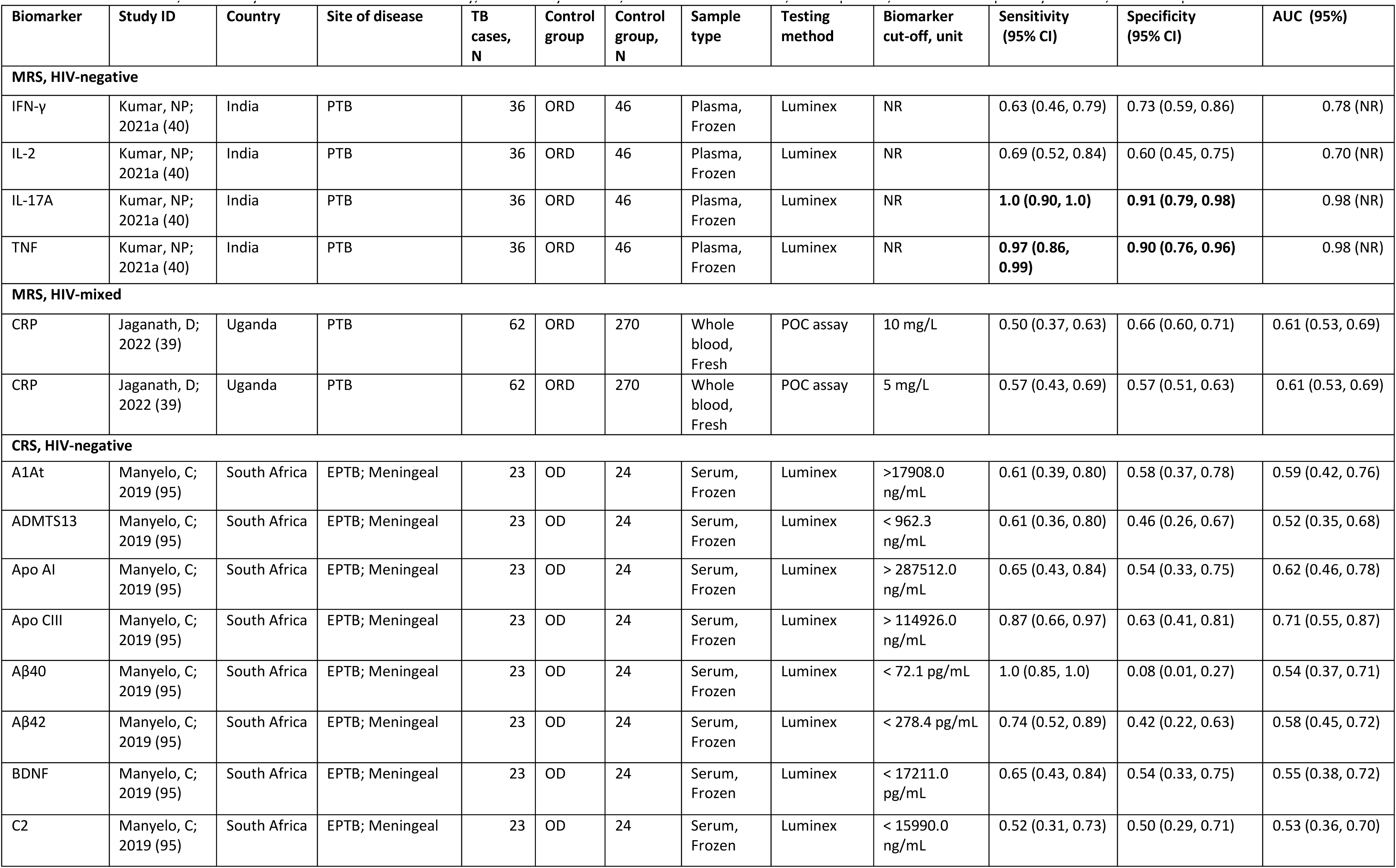
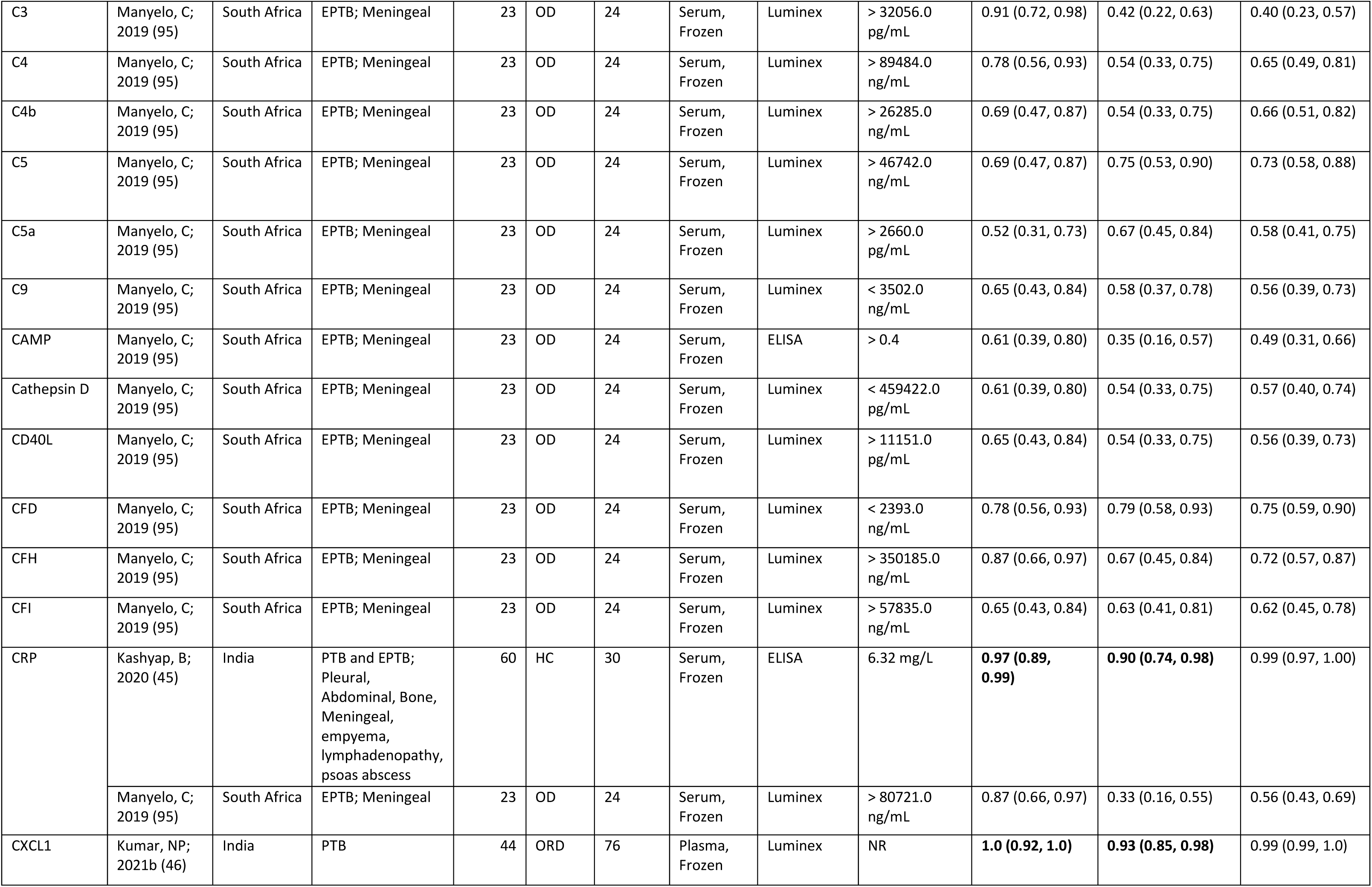
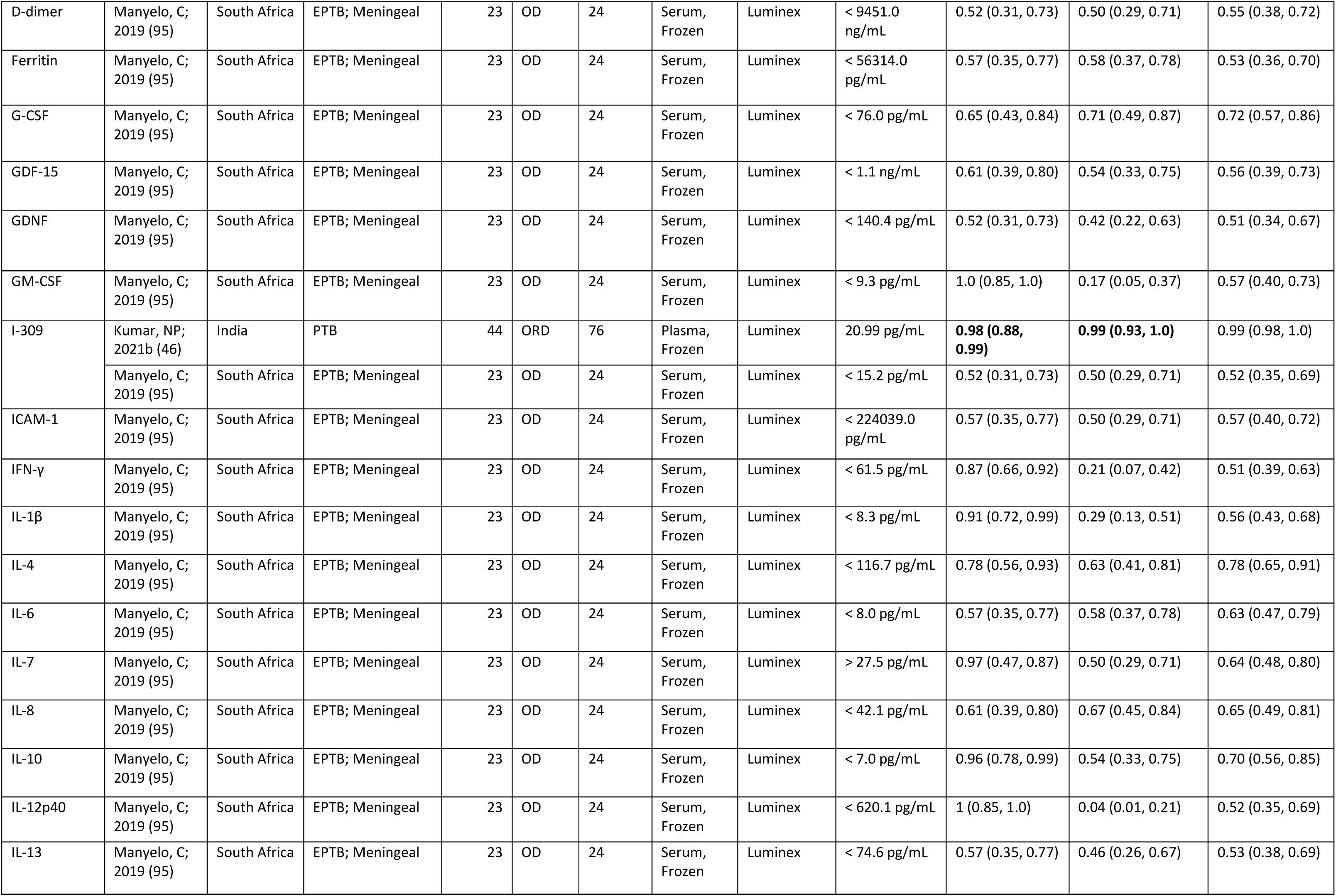
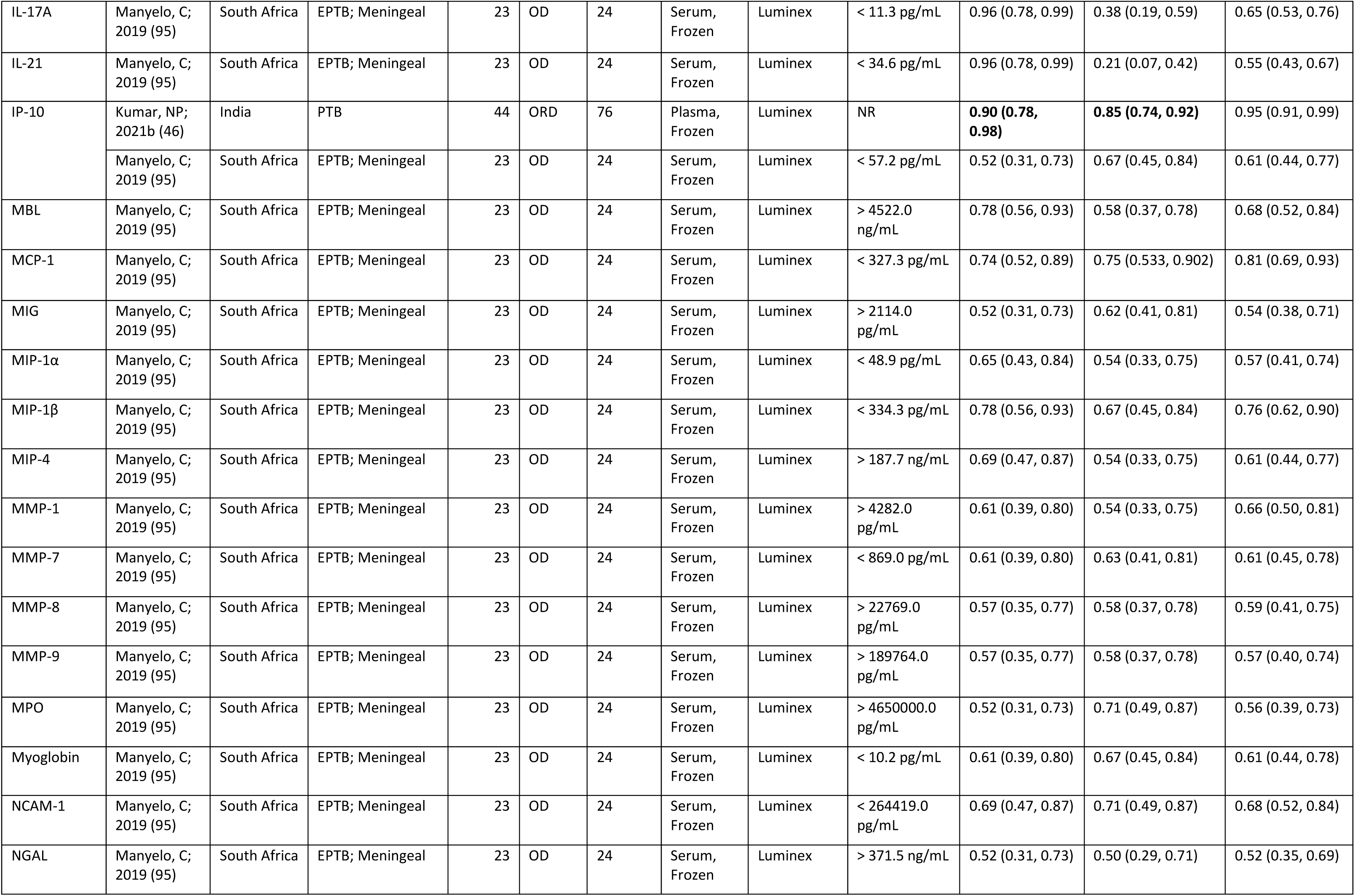
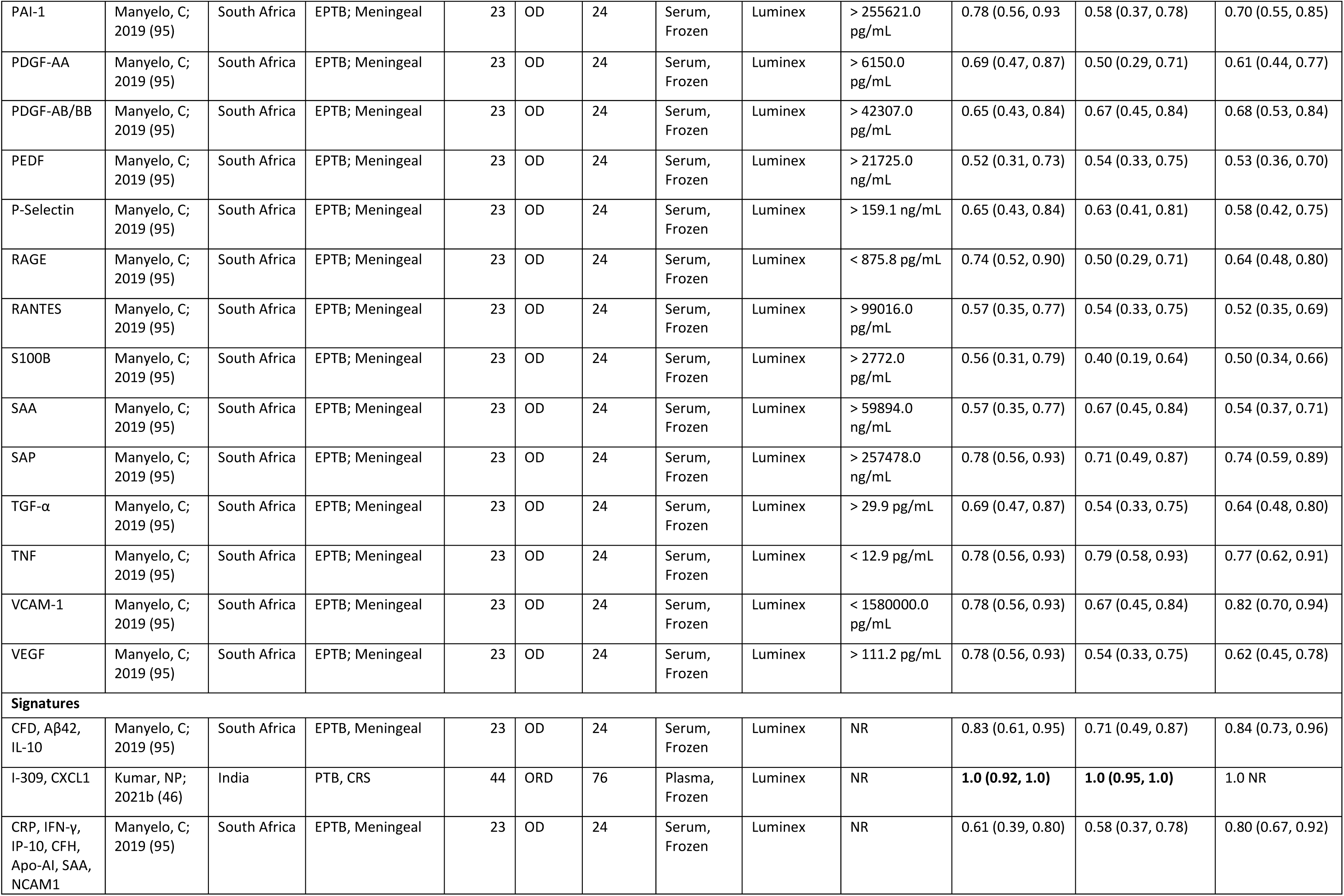
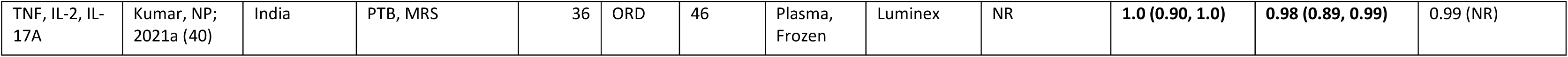
Childhood TB biomarkers. Biomarkers meeting TPP criteria are indicated in bold AUC=area under the curve, ELISA=enzyme-linked immunosorbent assay, HC= healthy controls, LTBI=latent TB infection, N=sample size, ORD=other respiratory diseases, NR=not reported

| Biomarker | Study ID | Country | Site of disease | TB cases, N | Control group | Control group, N | Sample type | Testing method | Biomarker cut-off, unit | Sensitivity (95% CI) | Specificity (95% CI) | AUC (95%) |
| --- | --- | --- | --- | --- | --- | --- | --- | --- | --- | --- | --- | --- |
| **MRS, HIV-negative** |  |  |  |  |  |  |  |  |  |  |  |  |
| IFN-γ | Kumar, NP; 2021a (40) | India | PTB | 36 | ORD | 46 | Plasma, Frozen | Luminex | NR | 0.63 (0.46, 0.79) | 0.73 (0.59, 0.86) | 0.78 (NR) |
| IL-2 | Kumar, NP; 2021a (40) | India | PTB | 36 | ORD | 46 | Plasma, Frozen | Luminex | NR | 0.69 (0.52, 0.84) | 0.60 (0.45, 0.75) | 0.70 (NR) |
| IL-17A | Kumar, NP; 2021a (40) | India | PTB | 36 | ORD | 46 | Plasma, Frozen | Luminex | NR | **1.0 (0.90, 1.0)** | **0.91 (0.79, 0.98)** | 0.98 (NR) |
| TNF | Kumar, NP; 2021a (40) | India | PTB | 36 | ORD | 46 | Plasma, Frozen | Luminex | NR | **0.97 (0.86, 0.99)** | **0.90 (0.76, 0.96)** | 0.98 (NR) |
| **MRS, HIV-mixed** |  |  |  |  |  |  |  |  |  |  |  |  |
| CRP | Jaganath, D; 2022 (39) | Uganda | PTB | 62 | ORD | 270 | Whole blood, Fresh | POC assay | 10 mg/L | 0.50 (0.37, 0.63) | 0.66 (0.60, 0.71) | 0.61 (0.53, 0.69) |
| CRP | Jaganath, D; 2022 (39) | Uganda | PTB | 62 | ORD | 270 | Whole blood, Fresh | POC assay | 5 mg/L | 0.57 (0.43, 0.69) | 0.57 (0.51, 0.63) | 0.61 (0.53, 0.69) |
| **CRS, HIV-negative** |  |  |  |  |  |  |  |  |  |  |  |  |
| A1At | Manyelo, C; 2019 (95) | South Africa | EPTB; Meningeal | 23 | OD | 24 | Serum, Frozen | Luminex | >17908.0 ng/mL | 0.61 (0.39, 0.80) | 0.58 (0.37, 0.78) | 0.59 (0.42, 0.76) |
| ADMTS13 | Manyelo, C; 2019 (95) | South Africa | EPTB; Meningeal | 23 | OD | 24 | Serum, Frozen | Luminex | < 962.3 ng/mL | 0.61 (0.36, 0.80) | 0.46 (0.26, 0.67) | 0.52 (0.35, 0.68) |
| Apo AI | Manyelo, C; 2019 (95) | South Africa | EPTB; Meningeal | 23 | OD | 24 | Serum, Frozen | Luminex | > 287512.0 ng/mL | 0.65 (0.43, 0.84) | 0.54 (0.33, 0.75) | 0.62 (0.46, 0.78) |
| Apo CIII | Manyelo, C; 2019 (95) | South Africa | EPTB; Meningeal | 23 | OD | 24 | Serum, Frozen | Luminex | > 114926.0 ng/mL | 0.87 (0.66, 0.97) | 0.63 (0.41, 0.81) | 0.71 (0.55, 0.87) |
| Aβ40 | Manyelo, C; 2019 (95) | South Africa | EPTB; Meningeal | 23 | OD | 24 | Serum, Frozen | Luminex | < 72.1 pg/mL | 1.0 (0.85, 1.0) | 0.08 (0.01, 0.27) | 0.54 (0.37, 0.71) |
| Aβ42 | Manyelo, C; 2019 (95) | South Africa | EPTB; Meningeal | 23 | OD | 24 | Serum, Frozen | Luminex | < 278.4 pg/mL | 0.74 (0.52, 0.89) | 0.42 (0.22, 0.63) | 0.58 (0.45, 0.72) |
| BDNF | Manyelo, C; 2019 (95) | South Africa | EPTB; Meningeal | 23 | OD | 24 | Serum, Frozen | Luminex | < 17211.0 pg/mL | 0.65 (0.43, 0.84) | 0.54 (0.33, 0.75) | 0.55 (0.38, 0.72) |
| C2 | Manyelo, C; 2019 (95) | South Africa | EPTB; Meningeal | 23 | OD | 24 | Serum, Frozen | Luminex | < 15990.0 ng/mL | 0.52 (0.31, 0.73) | 0.50 (0.29, 0.71) | 0.53 (0.36, 0.70) |
| C3 | Manyelo, C; 2019 (95) | South Africa | EPTB; Meningeal | 23 | OD | 24 | Serum, Frozen | Luminex | > 32056.0 pg/mL | 0.91 (0.72, 0.98) | 0.42 (0.22, 0.63) | 0.40 (0.23, 0.57) |
| C4 | Manyelo, C; 2019 (95) | South Africa | EPTB; Meningeal | 23 | OD | 24 | Serum, Frozen | Luminex | > 89484.0 ng/mL | 0.78 (0.56, 0.93) | 0.54 (0.33, 0.75) | 0.65 (0.49, 0.81) |
| C4b | Manyelo, C; 2019 (95) | South Africa | EPTB; Meningeal | 23 | OD | 24 | Serum, Frozen | Luminex | > 26285.0 ng/mL | 0.69 (0.47, 0.87) | 0.54 (0.33, 0.75) | 0.66 (0.51, 0.82) |
| C5 | Manyelo, C; 2019 (95) | South Africa | EPTB; Meningeal | 23 | OD | 24 | Serum, Frozen | Luminex | > 46742.0 ng/mL | 0.69 (0.47, 0.87) | 0.75 (0.53, 0.90) | 0.73 (0.58, 0.88) |
| C5a | Manyelo, C; 2019 (95) | South Africa | EPTB; Meningeal | 23 | OD | 24 | Serum, Frozen | Luminex | > 2660.0 pg/mL | 0.52 (0.31, 0.73) | 0.67 (0.45, 0.84) | 0.58 (0.41, 0.75) |
| C9 | Manyelo, C; 2019 (95) | South Africa | EPTB; Meningeal | 23 | OD | 24 | Serum, Frozen | Luminex | < 3502.0 ng/mL | 0.65 (0.43, 0.84) | 0.58 (0.37, 0.78) | 0.56 (0.39, 0.73) |
| CAMP | Manyelo, C; 2019 (95) | South Africa | EPTB; Meningeal | 23 | OD | 24 | Serum, Frozen | ELISA | > 0.4 | 0.61 (0.39, 0.80) | 0.35 (0.16, 0.57) | 0.49 (0.31, 0.66) |
| Cathepsin D | Manyelo, C; 2019 (95) | South Africa | EPTB; Meningeal | 23 | OD | 24 | Serum, Frozen | Luminex | < 459422.0 pg/mL | 0.61 (0.39, 0.80) | 0.54 (0.33, 0.75) | 0.57 (0.40, 0.74) |
| CD40L | Manyelo, C; 2019 (95) | South Africa | EPTB; Meningeal | 23 | OD | 24 | Serum, Frozen | Luminex | > 11151.0 pg/mL | 0.65 (0.43, 0.84) | 0.54 (0.33, 0.75) | 0.56 (0.39, 0.73) |
| CFD | Manyelo, C; 2019 (95) | South Africa | EPTB; Meningeal | 23 | OD | 24 | Serum, Frozen | Luminex | < 2393.0 ng/mL | 0.78 (0.56, 0.93) | 0.79 (0.58, 0.93) | 0.75 (0.59, 0.90) |
| CFH | Manyelo, C; 2019 (95) | South Africa | EPTB; Meningeal | 23 | OD | 24 | Serum, Frozen | Luminex | > 350185.0 ng/mL | 0.87 (0.66, 0.97) | 0.67 (0.45, 0.84) | 0.72 (0.57, 0.87) |
| CFI | Manyelo, C; 2019 (95) | South Africa | EPTB; Meningeal | 23 | OD | 24 | Serum, Frozen | Luminex | > 57835.0 ng/mL | 0.65 (0.43, 0.84) | 0.63 (0.41, 0.81) | 0.62 (0.45, 0.78) |
| CRP | Kashyap, B; 2020 (45) | India | PTB and EPTB; Pleural, Abdominal, Bone, Meningeal, empyema, lymphadenopathy, psoas abscess | 60 | HC | 30 | Serum, Frozen | ELISA | 6.32 mg/L | **0.97 (0.89, 0.99)** | **0.90 (0.74, 0.98)** | 0.99 (0.97, 1.00) |
|  | Manyelo, C; 2019 (95) | South Africa | EPTB; Meningeal | 23 | OD | 24 | Serum, Frozen | Luminex | > 80721.0 ng/mL | 0.87 (0.66, 0.97) | 0.33 (0.16, 0.55) | 0.56 (0.43, 0.69) |
| CXCL1 | Kumar, NP; 2021b (46) | India | PTB | 44 | ORD | 76 | Plasma, Frozen | Luminex | NR | **1.0 (0.92, 1.0)** | **0.93 (0.85, 0.98)** | 0.99 (0.99, 1.0) |
| D-dimer | Manyelo, C; 2019 (95) | South Africa | EPTB; Meningeal | 23 | OD | 24 | Serum, Frozen | Luminex | < 9451.0 ng/mL | 0.52 (0.31, 0.73) | 0.50 (0.29, 0.71) | 0.55 (0.38, 0.72) |
| Ferritin | Manyelo, C; 2019 (95) | South Africa | EPTB; Meningeal | 23 | OD | 24 | Serum, Frozen | Luminex | < 56314.0 pg/mL | 0.57 (0.35, 0.77) | 0.58 (0.37, 0.78) | 0.53 (0.36, 0.70) |
| G-CSF | Manyelo, C; 2019 (95) | South Africa | EPTB; Meningeal | 23 | OD | 24 | Serum, Frozen | Luminex | < 76.0 pg/mL | 0.65 (0.43, 0.84) | 0.71 (0.49, 0.87) | 0.72 (0.57, 0.86) |
| GDF-15 | Manyelo, C; 2019 (95) | South Africa | EPTB; Meningeal | 23 | OD | 24 | Serum, Frozen | Luminex | < 1.1 ng/mL | 0.61 (0.39, 0.80) | 0.54 (0.33, 0.75) | 0.56 (0.39, 0.73) |
| GDNF | Manyelo, C; 2019 (95) | South Africa | EPTB; Meningeal | 23 | OD | 24 | Serum, Frozen | Luminex | < 140.4 pg/mL | 0.52 (0.31, 0.73) | 0.42 (0.22, 0.63) | 0.51 (0.34, 0.67) |
| GM-CSF | Manyelo, C; 2019 (95) | South Africa | EPTB; Meningeal | 23 | OD | 24 | Serum, Frozen | Luminex | < 9.3 pg/mL | 1.0 (0.85, 1.0) | 0.17 (0.05, 0.37) | 0.57 (0.40, 0.73) |
| I-309 | Kumar, NP; 2021b (46) | India | PTB | 44 | ORD | 76 | Plasma, Frozen | Luminex | 20.99 pg/mL | **0.98 (0.88, 0.99)** | **0.99 (0.93, 1.0)** | 0.99 (0.98, 1.0) |
|  | Manyelo, C; 2019 (95) | South Africa | EPTB; Meningeal | 23 | OD | 24 | Serum, Frozen | Luminex | < 15.2 pg/mL | 0.52 (0.31, 0.73) | 0.50 (0.29, 0.71) | 0.52 (0.35, 0.69) |
| ICAM-1 | Manyelo, C; 2019 (95) | South Africa | EPTB; Meningeal | 23 | OD | 24 | Serum, Frozen | Luminex | < 224039.0 pg/mL | 0.57 (0.35, 0.77) | 0.50 (0.29, 0.71) | 0.57 (0.40, 0.72) |
| IFN-γ | Manyelo, C; 2019 (95) | South Africa | EPTB; Meningeal | 23 | OD | 24 | Serum, Frozen | Luminex | < 61.5 pg/mL | 0.87 (0.66, 0.92) | 0.21 (0.07, 0.42) | 0.51 (0.39, 0.63) |
| IL-1β | Manyelo, C; 2019 (95) | South Africa | EPTB; Meningeal | 23 | OD | 24 | Serum, Frozen | Luminex | < 8.3 pg/mL | 0.91 (0.72, 0.99) | 0.29 (0.13, 0.51) | 0.56 (0.43, 0.68) |
| IL-4 | Manyelo, C; 2019 (95) | South Africa | EPTB; Meningeal | 23 | OD | 24 | Serum, Frozen | Luminex | < 116.7 pg/mL | 0.78 (0.56, 0.93) | 0.63 (0.41, 0.81) | 0.78 (0.65, 0.91) |
| IL-6 | Manyelo, C; 2019 (95) | South Africa | EPTB; Meningeal | 23 | OD | 24 | Serum, Frozen | Luminex | < 8.0 pg/mL | 0.57 (0.35, 0.77) | 0.58 (0.37, 0.78) | 0.63 (0.47, 0.79) |
| IL-7 | Manyelo, C; 2019 (95) | South Africa | EPTB; Meningeal | 23 | OD | 24 | Serum, Frozen | Luminex | > 27.5 pg/mL | 0.97 (0.47, 0.87) | 0.50 (0.29, 0.71) | 0.64 (0.48, 0.80) |
| IL-8 | Manyelo, C; 2019 (95) | South Africa | EPTB; Meningeal | 23 | OD | 24 | Serum, Frozen | Luminex | < 42.1 pg/mL | 0.61 (0.39, 0.80) | 0.67 (0.45, 0.84) | 0.65 (0.49, 0.81) |
| IL-10 | Manyelo, C; 2019 (95) | South Africa | EPTB; Meningeal | 23 | OD | 24 | Serum, Frozen | Luminex | < 7.0 pg/mL | 0.96 (0.78, 0.99) | 0.54 (0.33, 0.75) | 0.70 (0.56, 0.85) |
| IL-12p40 | Manyelo, C; 2019 (95) | South Africa | EPTB; Meningeal | 23 | OD | 24 | Serum, Frozen | Luminex | < 620.1 pg/mL | 1 (0.85, 1.0) | 0.04 (0.01, 0.21) | 0.52 (0.35, 0.69) |
| IL-13 | Manyelo, C; 2019 (95) | South Africa | EPTB; Meningeal | 23 | OD | 24 | Serum, Frozen | Luminex | < 74.6 pg/mL | 0.57 (0.35, 0.77) | 0.46 (0.26, 0.67) | 0.53 (0.38, 0.69) |
| IL-17A | Manyelo, C; 2019 (95) | South Africa | EPTB; Meningeal | 23 | OD | 24 | Serum, Frozen | Luminex | < 11.3 pg/mL | 0.96 (0.78, 0.99) | 0.38 (0.19, 0.59) | 0.65 (0.53, 0.76) |
| IL-21 | Manyelo, C; 2019 (95) | South Africa | EPTB; Meningeal | 23 | OD | 24 | Serum, Frozen | Luminex | < 34.6 pg/mL | 0.96 (0.78, 0.99) | 0.21 (0.07, 0.42) | 0.55 (0.43, 0.67) |
| IP-10 | Kumar, NP; 2021b (46) | India | PTB | 44 | ORD | 76 | Plasma, Frozen | Luminex | NR | **0.90 (0.78, 0.98)** | **0.85 (0.74, 0.92)** | 0.95 (0.91, 0.99) |
|  | Manyelo, C; 2019 (95) | South Africa | EPTB; Meningeal | 23 | OD | 24 | Serum, Frozen | Luminex | < 57.2 pg/mL | 0.52 (0.31, 0.73) | 0.67 (0.45, 0.84) | 0.61 (0.44, 0.77) |
| MBL | Manyelo, C; 2019 (95) | South Africa | EPTB; Meningeal | 23 | OD | 24 | Serum, Frozen | Luminex | > 4522.0 ng/mL | 0.78 (0.56, 0.93) | 0.58 (0.37, 0.78) | 0.68 (0.52, 0.84) |
| MCP-1 | Manyelo, C; 2019 (95) | South Africa | EPTB; Meningeal | 23 | OD | 24 | Serum, Frozen | Luminex | < 327.3 pg/mL | 0.74 (0.52, 0.89) | 0.75 (0.533, 0.902) | 0.81 (0.69, 0.93) |
| MIG | Manyelo, C; 2019 (95) | South Africa | EPTB; Meningeal | 23 | OD | 24 | Serum, Frozen | Luminex | > 2114.0 pg/mL | 0.52 (0.31, 0.73) | 0.62 (0.41, 0.81) | 0.54 (0.38, 0.71) |
| MIP-1α | Manyelo, C; 2019 (95) | South Africa | EPTB; Meningeal | 23 | OD | 24 | Serum, Frozen | Luminex | < 48.9 pg/mL | 0.65 (0.43, 0.84) | 0.54 (0.33, 0.75) | 0.57 (0.41, 0.74) |
| MIP-1β | Manyelo, C; 2019 (95) | South Africa | EPTB; Meningeal | 23 | OD | 24 | Serum, Frozen | Luminex | < 334.3 pg/mL | 0.78 (0.56, 0.93) | 0.67 (0.45, 0.84) | 0.76 (0.62, 0.90) |
| MIP-4 | Manyelo, C; 2019 (95) | South Africa | EPTB; Meningeal | 23 | OD | 24 | Serum, Frozen | Luminex | > 187.7 ng/mL | 0.69 (0.47, 0.87) | 0.54 (0.33, 0.75) | 0.61 (0.44, 0.77) |
| MMP-1 | Manyelo, C; 2019 (95) | South Africa | EPTB; Meningeal | 23 | OD | 24 | Serum, Frozen | Luminex | > 4282.0 pg/mL | 0.61 (0.39, 0.80) | 0.54 (0.33, 0.75) | 0.66 (0.50, 0.81) |
| MMP-7 | Manyelo, C; 2019 (95) | South Africa | EPTB; Meningeal | 23 | OD | 24 | Serum, Frozen | Luminex | < 869.0 pg/mL | 0.61 (0.39, 0.80) | 0.63 (0.41, 0.81) | 0.61 (0.45, 0.78) |
| MMP-8 | Manyelo, C; 2019 (95) | South Africa | EPTB; Meningeal | 23 | OD | 24 | Serum, Frozen | Luminex | > 22769.0 pg/mL | 0.57 (0.35, 0.77) | 0.58 (0.37, 0.78) | 0.59 (0.41, 0.75) |
| MMP-9 | Manyelo, C; 2019 (95) | South Africa | EPTB; Meningeal | 23 | OD | 24 | Serum, Frozen | Luminex | > 189764.0 pg/mL | 0.57 (0.35, 0.77) | 0.58 (0.37, 0.78) | 0.57 (0.40, 0.74) |
| MPO | Manyelo, C; 2019 (95) | South Africa | EPTB; Meningeal | 23 | OD | 24 | Serum, Frozen | Luminex | > 4650000.0 pg/mL | 0.52 (0.31, 0.73) | 0.71 (0.49, 0.87) | 0.56 (0.39, 0.73) |
| Myoglobin | Manyelo, C; 2019 (95) | South Africa | EPTB; Meningeal | 23 | OD | 24 | Serum, Frozen | Luminex | < 10.2 pg/mL | 0.61 (0.39, 0.80) | 0.67 (0.45, 0.84) | 0.61 (0.44, 0.78) |
| NCAM-1 | Manyelo, C; 2019 (95) | South Africa | EPTB; Meningeal | 23 | OD | 24 | Serum, Frozen | Luminex | < 264419.0 pg/mL | 0.69 (0.47, 0.87) | 0.71 (0.49, 0.87) | 0.68 (0.52, 0.84) |
| NGAL | Manyelo, C; 2019 (95) | South Africa | EPTB; Meningeal | 23 | OD | 24 | Serum, Frozen | Luminex | > 371.5 ng/mL | 0.52 (0.31, 0.73) | 0.50 (0.29, 0.71) | 0.52 (0.35, 0.69) |
| PAI-1 | Manyelo, C; 2019 (95) | South Africa | EPTB; Meningeal | 23 | OD | 24 | Serum, Frozen | Luminex | > 255621.0 pg/mL | 0.78 (0.56, 0.93) | 0.58 (0.37, 0.78) | 0.70 (0.55, 0.85) |
| PDGF-AA | Manyelo, C; 2019 (95) | South Africa | EPTB; Meningeal | 23 | OD | 24 | Serum, Frozen | Luminex | > 6150.0 pg/mL | 0.69 (0.47, 0.87) | 0.50 (0.29, 0.71) | 0.61 (0.44, 0.77) |
| PDGF-AB/BB | Manyelo, C; 2019 (95) | South Africa | EPTB; Meningeal | 23 | OD | 24 | Serum, Frozen | Luminex | > 42307.0 pg/mL | 0.65 (0.43, 0.84) | 0.67 (0.45, 0.84) | 0.68 (0.53, 0.84) |
| PEDF | Manyelo, C; 2019 (95) | South Africa | EPTB; Meningeal | 23 | OD | 24 | Serum, Frozen | Luminex | > 21725.0 ng/mL | 0.52 (0.31, 0.73) | 0.54 (0.33, 0.75) | 0.53 (0.36, 0.70) |
| P-Selectin | Manyelo, C; 2019 (95) | South Africa | EPTB; Meningeal | 23 | OD | 24 | Serum, Frozen | Luminex | > 159.1 ng/mL | 0.65 (0.43, 0.84) | 0.63 (0.41, 0.81) | 0.58 (0.42, 0.75) |
| RAGE | Manyelo, C; 2019 (95) | South Africa | EPTB; Meningeal | 23 | OD | 24 | Serum, Frozen | Luminex | < 875.8 pg/mL | 0.74 (0.52, 0.90) | 0.50 (0.29, 0.71) | 0.64 (0.48, 0.80) |
| RANTES | Manyelo, C; 2019 (95) | South Africa | EPTB; Meningeal | 23 | OD | 24 | Serum, Frozen | Luminex | > 99016.0 pg/mL | 0.57 (0.35, 0.77) | 0.54 (0.33, 0.75) | 0.52 (0.35, 0.69) |
| S100B | Manyelo, C; 2019 (95) | South Africa | EPTB; Meningeal | 23 | OD | 24 | Serum, Frozen | Luminex | > 2772.0 pg/mL | 0.56 (0.31, 0.79) | 0.40 (0.19, 0.64) | 0.50 (0.34, 0.66) |
| SAA | Manyelo, C; 2019 (95) | South Africa | EPTB; Meningeal | 23 | OD | 24 | Serum, Frozen | Luminex | > 59894.0 ng/mL | 0.57 (0.35, 0.77) | 0.67 (0.45, 0.84) | 0.54 (0.37, 0.71) |
| SAP | Manyelo, C; 2019 (95) | South Africa | EPTB; Meningeal | 23 | OD | 24 | Serum, Frozen | Luminex | > 257478.0 ng/mL | 0.78 (0.56, 0.93) | 0.71 (0.49, 0.87) | 0.74 (0.59, 0.89) |
| TGF-α | Manyelo, C; 2019 (95) | South Africa | EPTB; Meningeal | 23 | OD | 24 | Serum, Frozen | Luminex | > 29.9 pg/mL | 0.69 (0.47, 0.87) | 0.54 (0.33, 0.75) | 0.64 (0.48, 0.80) |
| TNF | Manyelo, C; 2019 (95) | South Africa | EPTB; Meningeal | 23 | OD | 24 | Serum, Frozen | Luminex | < 12.9 pg/mL | 0.78 (0.56, 0.93) | 0.79 (0.58, 0.93) | 0.77 (0.62, 0.91) |
| VCAM-1 | Manyelo, C; 2019 (95) | South Africa | EPTB; Meningeal | 23 | OD | 24 | Serum, Frozen | Luminex | < 1580000.0 pg/mL | 0.78 (0.56, 0.93) | 0.67 (0.45, 0.84) | 0.82 (0.70, 0.94) |
| VEGF | Manyelo, C; 2019 (95) | South Africa | EPTB; Meningeal | 23 | OD | 24 | Serum, Frozen | Luminex | > 111.2 pg/mL | 0.78 (0.56, 0.93) | 0.54 (0.33, 0.75) | 0.62 (0.45, 0.78) |
| **Signatures** |  |  |  |  |  |  |  |  |  |  |  |  |
| CFD, Aβ42, IL-10 | Manyelo, C; 2019 (95) | South Africa | EPTB, Meningeal | 23 | OD | 24 | Serum, Frozen | Luminex | NR | 0.83 (0.61, 0.95) | 0.71 (0.49, 0.87) | 0.84 (0.73, 0.96) |
| I-309, CXCL1 | Kumar, NP; 2021b (46) | India | PTB, CRS | 44 | ORD | 76 | Plasma, Frozen | Luminex | NR | **1.0 (0.92, 1.0)** | **1.0 (0.95, 1.0)** | 1.0 NR |
| CRP, IFN-γ, IP-10, CFH, Apo-AI, SAA, NCAM1 | Manyelo, C; 2019 (95) | South Africa | EPTB, Meningeal | 23 | OD | 24 | Serum, Frozen | Luminex | NR | 0.61 (0.39, 0.80) | 0.58 (0.37, 0.78) | 0.80 (0.67, 0.92) |
| TNF, IL-2, IL-17A | Kumar, NP; 2021a (40) | India | PTB, MRS | 36 | ORD | 46 | Plasma, Frozen | Luminex | NR | **1.0 (0.90, 1.0)** | **0.98 (0.89, 0.99)** | 0.99 (NR) |

## Data Availability

All data produced in the present work are contained in the manuscript

